# Defining influenza epidemic zones through temporal clustering of global surveillance data

**DOI:** 10.64898/2026.04.17.26351048

**Authors:** Norman Hassell, Perrine Marcenac, Cédric S. Bationo, Siddhivinayak Hirve, Stefano Tempia, Melissa A. Rolfes, Lindsey M. Duca, Aspen Hammond, Pushpa R. Wijesinghe, Jean Michel Heraud, Dmitriy Pereyaslov, Wenqing Zhang, Rebecca J. Kondor, Eduardo Azziz-Baumgartner

## Abstract

**Introduction:** Modeling when influenza epidemics typically occur can help countries optimize surveillance, time clinical and public health interventions, and reduce the burden of influenza.

**Methods:** We used influenza virus detections reported during 2011–2024 by 180 countries to the Global Influenza Surveillance and Response System, excluding years impacted by the COVID-19 pandemic (2020–2023). We analyzed data by calendar year (week 1–52) or shifted year (week 30–29) time windows, based on when most influenza detections occurred in each country. For countries with sufficient data, we computed generalized additive models (GAMs) of each country’s weekly influenza-positive tests to smooth and impute time series distributions. From these GAMs, we calculated each country’s weekly influenza burden. Country-specific time series were grouped using ensemble consensus k-means clustering, reducing the Euclidean distance between time series within clusters. We calculated cluster-specific GAMs to estimate average seasonal timing. Countries without sufficient data were assigned to a cluster based on population-weighted latitudinal distance to a cluster’s mean latitude.

**Results:** We identified five clusters, or epidemic zones, from 111 countries with sufficient data. The influenza burden in epidemic zones A and B was consistent with a northern hemisphere pattern, with most influenza detections occurring during November–May, while epidemic zones D and E were characterized by southern hemisphere-like seasonal timing, with most influenza burden occurring during June–November (D) and May–November (E). Epidemic zone C had most influenza burden occurring during September–March; most countries assigned to this cluster were in the tropics.

**Conclusion:** Epidemic zones may serve as a useful tool to strengthen and optimize influenza surveillance for global health decision-making (e.g., during vaccine strain composition discussions) and to guide country preparedness efforts for seasonal influenza epidemics, including timing enhanced surveillance and informing the availability of prevention and control efforts.

**What is already known on this topic:** Previous initiatives to provide a framework for describing global influenza patterns and to support national and regional prevention and control strategies have classified countries into influenza transmission zones, based primarily on geographic proximity. Many countries have improved their surveillance systems for respiratory viruses, providing an opportunity to re-assess patterns of influenza dynamics using analytic approaches focused on influenza detection data and maximizing global coverage.

**What this study adds:** Optimal clustering of influenza surveillance data from 111 countries and proximal assignment by latitude of countries lacking sufficient data grouped countries into five epidemic zones.

**How this study might affect research, practice or policy:** By providing improved resolution on the temporal and geographical dynamics of influenza activity, this study offers an evidence base to aid national and global decision-making on enhanced surveillance strategies, seasonal epidemic preparedness, and resource allocation, thereby strengthening efforts to prevent and control influenza worldwide. This data-driven framework for characterizing global patterns of influenza virus circulation can be leveraged to reexamine patterns as new data become available.

## INTRODUCTION

Influenza virus is characterized by seasonal epidemics every year and causes significant morbidity and mortality globally. When analyzing global data, geographic regions of countries, areas, and territories are often the finest unit of analysis available. We will refer to these geographic units as “countries” for simplified terminology. In temperate climates, most influenza illnesses occur during the colder, winter months of the year; countries in the northern hemisphere (NH) have most influenza illnesses October–April, while southern hemisphere (SH) countries have most influenza illnesses May–September.^1–3^ The epidemiology of influenza in countries with tropical and subtropical climates is less clear. Countries may have multiple peaks of influenza detections throughout the year, year-round activity with no clear epidemic peaks, or have peaks during periods that are not aligned with typical NH or SH epidemic periods.^2–4^

Since 2011, several efforts have grouped countries together into, so called, influenza transmission zones based on geographic proximity and similarities in the timing of influenza detections.^2–8^ Influenza transmission zones were intended to concisely describe influenza activity globally and inform prevention and control efforts in countries, including what influenza vaccine formulation to select and when to start influenza vaccine campaigns. Different approaches were used to summarize typical influenza activity—aggregating weekly case proportion data from multiple seasons into summary curves or calculating a monthly case proportion of positive cases—to find periods, or seasons, when influenza activity typically increased.^2,4,5^

The zones currently in use^9^ were last reviewed in 2015 during a World Health Organization (WHO) expert consultation and relied on geographical regions with adjustments based on influenza trends.^6^ Since then, many countries have improved their surveillance systems for respiratory viruses, providing an opportunity to re-assess patterns of influenza dynamics using more recent data and possibly incorporate additional countries in the analysis.^5,10–12^ We wanted to expand upon the prior efforts using analytic approaches that rely primarily on influenza detection data to redefine and improve confidence in clustering countries into updated influenza epidemic zones.

## METHODS

### Data sources

The Global Influenza Surveillance and Response System (GISRS) has publicly-available data on virologic surveillance of influenza viruses through FluNet,^13^ including the weekly number of laboratory-confirmed specimens positive for influenza virus by type, subtype, and lineage.

Countries may collect these data through sentinel and other systematically conducted virologic surveillance. We included consolidated influenza surveillance data from January 2011– December 2019 and July 2023–December 2024 for all countries, where these data were available. We excluded data reported during January 2020–June 2023 because the COVID-19 pandemic altered influenza virus circulation and reporting patterns.

### Data analysis

#### Analysis time windows

We categorized countries into two time units of analysis to center each country’s distributions of influenza detections: a calendar-year or shifted-year time window. Calendar-year time windows organize influenza data from week 1–52, as typically done for countries with SH-like influenza activity, while shifted-year time windows organize data from week 30–week 29 of the next year, characteristic of countries with NH-like activity. We assessed distributions of influenza detections over a 52-week calendar year for each year of available data and determined the calendar quarter that most frequently had the maximum density (mode). We defined calendar quarters (Q) from 21 December–19 March (Q1), 20 March–20 June (Q2), 21 June–21 September (Q3), and 22 September–20 December (Q4). For countries where the peak density occurred most frequently during Q1 or Q4, we analyzed data using a shifted-year window. For those with peaks occurring most frequently during Q2 or Q3, we used a calendar-year window. Countries with identical peaks in ≥1 quarter or with no influenza detections during our analytic years were excluded. For the main analysis, we included time windows (individual calendar-year or shifted-year units) where ≥100 influenza-positive specimens were detected and ≥40% of the time-window weeks had data reported, including partial time windows that met these criteria among countries in the shifted-year unit analytic group (i.e., 2011 weeks 1–30 or 2024 weeks 31–52). To ensure sufficient data for the time series clustering analysis, we further restricted to countries with ≥5 complete time windows (i.e., ≥260 weeks).

#### Data imputation and smoothing

For weeks without reported data, we imputed missing influenza detections and smoothed each country’s time series using generalized additive models (GAMs) on each period of continuous data for each country (**Supplementary Figure 1**). Models used a negative binomial distribution, thin plate regression spline, and restricted maximum likelihood (REML) smoothing. We set the basis dimension (knots) parameter to 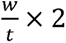, where represents the number of weeks in a period of continuous data, and represents the total number of available time-window years. When the boundary weeks of a country’s time series had missing data, we set that week’s value to zero influenza-positive specimens (zero padded) to anchor the smooth at the time window edges instead of leaving it to extrapolate unconstrained from the nearest interior observation.

We selected the penalty term m=1 to penalize the first derivative slope and allow the fitted curve to flatten smoothly toward a constant level in instances when boundary weeks were zero-padded. For countries where ≥20% of data was missing from a particular period, m was auto-initialized.

We used the fitted values from the GAMs to define each country’s weekly influenza burden, which we defined as the proportion of influenza detections from each unique time-window year (e.g., 2011–2012, 2012–2013, […] for countries in the shifted year partition; 2011, 2012, […] for countries in the calendar year partition) that occurred in a given week.

#### Clustering into influenza epidemic zones

Separately for the countries grouped by shifted-year or calendar-year windows, we generated matrices for each unique time-window year with each country’s weekly burden; each matrix was weighted by the number of weeks in that particular time-window year to account for partial years. Countries had to have ≥1 complete country-to-country time-window year available for comparison and for later determination of ideal cluster size (k), so we included only countries that had data reported during the time-window year with maximum country representation (anchor year: 2019 for calendar-year and 2015 for shifted-year partitions). We then implemented an ensemble consensus clustering approach:^14^ for each time-window year, we ran a resampling scheme drawing 90% of that year’s countries without replacement and clustered these sampled countries together based on Euclidean distance. We ran this resampling scheme for each time-window year and ensembled these data into a consensus matrix with each country pair’s co-clustering frequency. We used an ensemble consensus matrix generated from 100 resamples across all time-window years to calculate Calinski-Harabasz indices, Dunn indices, Silhouette scores, and proportion of ambiguous clustering (PAC) scores for each algorithm (k-means, hierarchical k-means, hierarchical, and partitioning around medoids [PAM]) and k-value. We then calculated a composite score (range: 0–1) that merged these four per-algorithm quality metrics into one score for the comparison of algorithms at each k, with values closer to 1 indicating better algorithm performance. The composite score was the mean of the four metrics normalized to their range for each k value; we converted the PAC score to 1-PAC, so that for all four metrics, values closer to 1 indicated better performance. Scores were only comparable between algorithms at the same k, and all four metrics were weighted equally by default. We used an ensemble consensus matrix generated from 100 resamples in the calendar and shifted anchor years to calculate the within-cluster sum of squares (WSS), mean Silhouette width, and Calinski-Harabasz index for determining the optimal k values of the calendar and shifted-year partitions. Optimal k was identified as the WSS “elbow,” maximum mean Silhouette width, and Calinski-Harabasz index peak for each time-window partition. For our final ensemble clustering, we resampled data 1,000 times and used spectral clustering to assign countries to clusters from the ensemble consensus matrix. To assess the stability of country assignments to clusters, we calculated a consensus score for each country which averaged the pairwise consensus value between each country and its co-clustered countries to determine how consistently a given country was grouped with co-clustered countries across resamples; higher scores demonstrated greater stability. We also calculated a cluster consensus score by taking the mean of constituent country consensus scores. We named the resulting clusters “influenza epidemic zones.”

#### Estimating seasonal timing of influenza

For each epidemic zone, we generated summary seasonal curves to estimate the typical start and end of each zone’s influenza epidemics. We ran hierarchical GAMs for each cluster on country-year influenza burden data using a Tweedie log-linked family, REML smoothing, and two additive smooth terms: a shared cyclic smooth term to wrap week 52 to week 1 to permit seasonal continuity, and a country-specific factor-smooth term to account for between-country variability with the cyclic smooth term. The fitted curves and 95% confidence intervals were normalized to probability curves to better demonstrate relative influenza burden over the year. We implemented the same approach to calculate country GAMs, removing the country-specific factor-smooth term, and estimate country-specific seasonal timing. We identified the main influenza epidemic for clusters and countries, which we defined as the influenza epidemic with the longest duration. We defined the start of influenza epidemics as the first week in which the weekly relative influenza burden exceeded the median of the weekly relative influenza burden values for ≥3 weeks and the end of influenza epidemics as the first week in which the burden remained below the median for ≥3 weeks.^3,15^ We used area under the curve calculations to estimate what proportion of influenza activity occurred during the main epidemic. We converted calendar weeks to months using the average number of weeks per month (4.345).

#### Assigning countries with limited influenza data to epidemic zones

For countries ^16–18^ with limited or no influenza detection data in FluNet, we proposed to use latitude as a proxy to assign these countries to epidemic zones. We calculated a population-weighted mean latitude for each country using the latitude of cities^19^ and city population size^20^ and a mean latitude for each zone from the population-weighted mean latitudes of constituent countries. To test the robustness of this proxying approach, we first used one-way ANOVA to test whether the mean population-weighted latitude of countries within each cluster differed across epidemic zones. Secondly, we implemented a leave-one-out approach with the countries that had sufficient data to be included in the primary clustering analysis, whereby we sequentially left countries out of each clustering analysis partition, recomputed the ensemble consensus clustering approach as previously described (using the optimal algorithm, optimal k, and 1,000 resamples), and then proxied the country that was left out to the cluster with the minimum difference between the country’s population-weighted mean latitude and the mean cluster latitude. We then calculated the proportion of proxying success per cluster as the number of countries that were assigned to the same cluster via the original clustering and proxying approaches over the total number of countries. Following these robustness checks, we assigned countries with insufficient or no data to the closest epidemic zone based on population-weighted mean latitude.

## RESULTS

Of the 180 countries that submitted influenza virus testing data to FluNet during 2011 2019 and 2023–2024, 126 (70%) had most of their influenza detections during Q1 and Q4 and were classified into a shifted-year window of analysis, 49 (27%) had most of their detections during Q2 and Q3 and were classified into a calendar-year window of analysis, and 5 (3%) had insufficient data to determine the most appropriate unit of analysis (**Supplementary Table 1**). Of the 175 (97%) countries for whom the window of analysis could be determined, 111 (63%) were included in the clustering analysis (**Figure 1**). We excluded 63 (36% of 175) countries because they had <5 complete time windows with sufficient data for analysis after including only analytic time windows that had ≥100 influenza detections and ≥40% of weeks with data reported. We excluded one (<1%) country as it had insufficient data during the anchor year. All continents had countries that had insufficient or no data in FluNet and thus were not clustered in the primary analysis; most of these countries were in Africa and the Americas.

**Figure 1:**
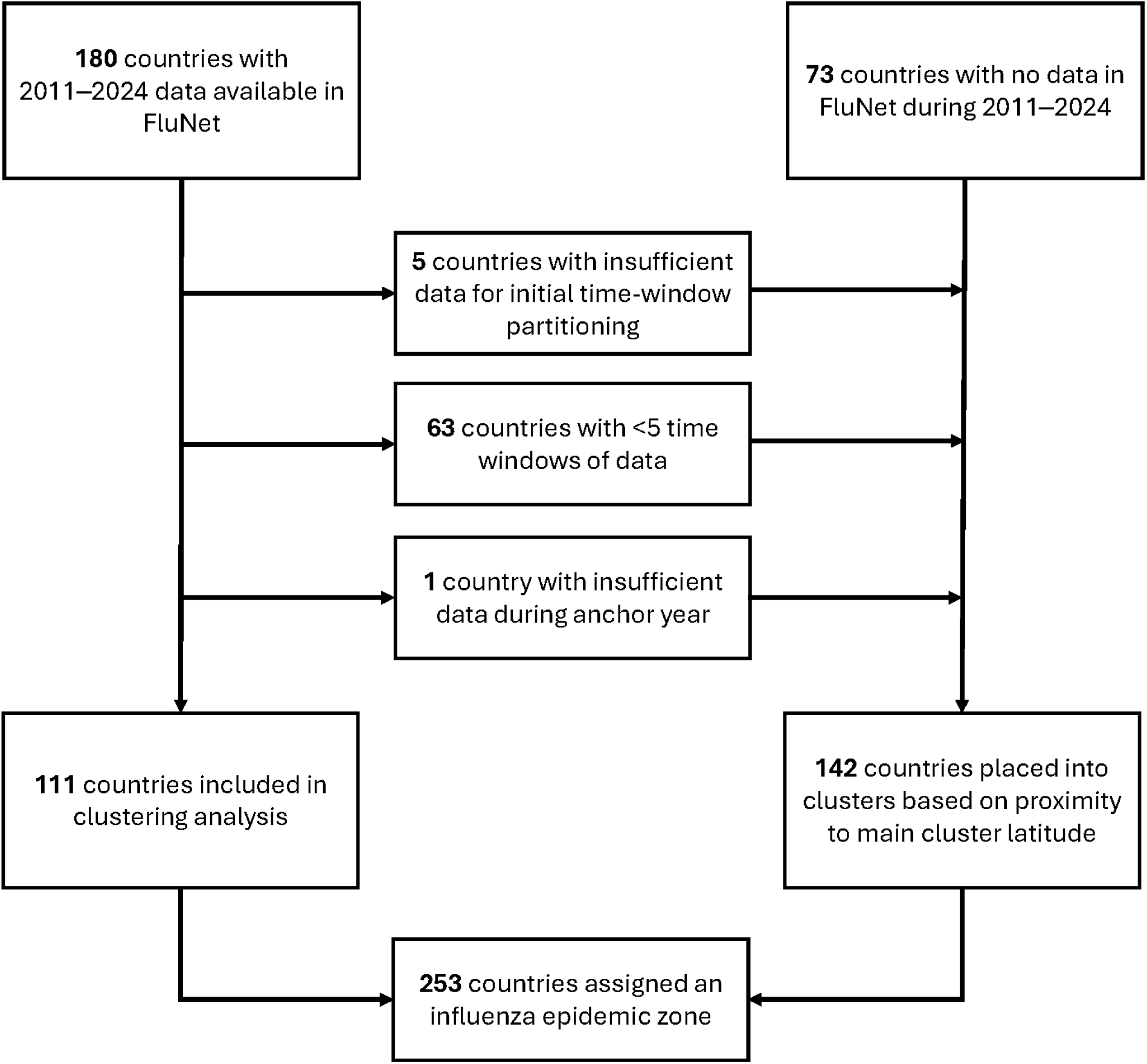
Schematic of inclusion and exclusion of countries into the analysis to cluster countries into influenza epidemic zones.

K-means and hierarchical k-means outperformed hierarchical and PAM clustering for both calendar and shifted year partitions (**Supplementary Figure 2**). K-means modestly outperformed hierarchical k-means in the shifted year partition, and so we selected it as our optimal clustering algorithm. Optimal cluster size was three for countries analyzed with a shifted-year time window, labeled as epidemic zones A, B, and C, and two for countries analyzed with a calendar-year time window, labeled as zones D and E, for a total of five globally distributed epidemic zones (**Supplementary Figure 2–3**). Consensus scores to evaluate cluster stability were ≥60% for all epidemic zones except for epidemic zone B, which had a score of 47% (**Supplementary Table 2**). Country stability within clusters had a wide range, with consensus scores from 30% (Kyrgyzstan in zone B) to 91% (Lao PDR, Cameroon, Côte d’Ivoire, and Nicaragua in zone C) (**Supplementary Table 3**).

We sought to assign the 69 countries with insufficient influenza detection data and 73 countries with no data in FluNet to these influenza epidemic zones. We observed a strong association with country population-weighted mean latitude in each cluster (**Figure 2**). Moreover, for countries included in the original clustering analysis, leaving individual countries out and assigning them to a zone based on their population-weighted mean latitude showed that, on average, 77% of left-out countries were correctly assigned to their original epidemic zone (range: 68–86%) (**Supplementary Table 4**). Therefore, we assigned the 142 countries with insufficient influenza data to be included in the primary clustering analysis to one of the five epidemic zones using their population-weighted mean latitudes (**Figure 3, Supplementary Table 5**).

**Figure 2:**
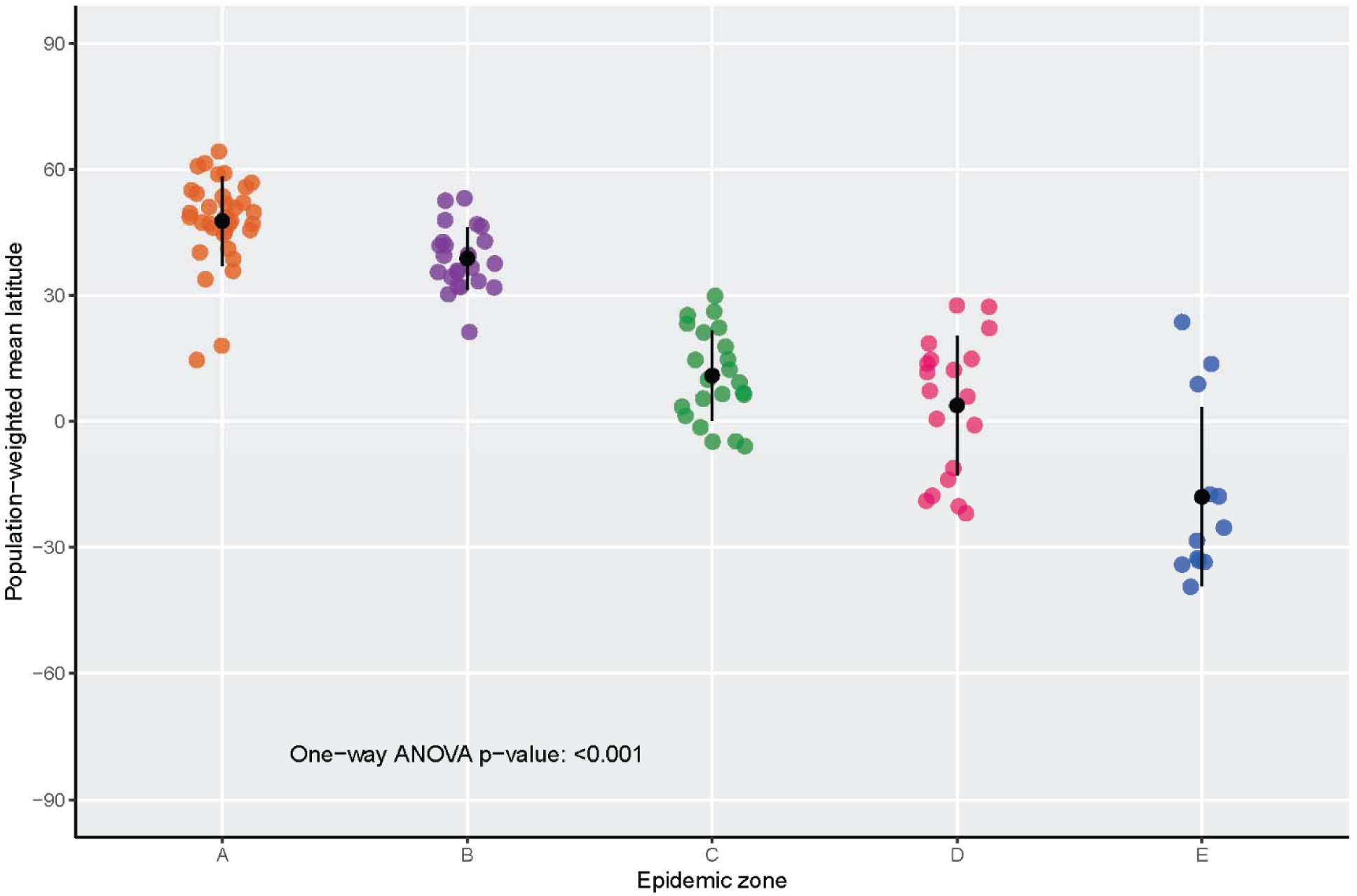
Population-weighted latitude of countries clustered into each epidemic zone, with each dot representing a country assigned to that cluster. Mean and one standard deviation within the cluster are shown in black.

**Figure 3:**
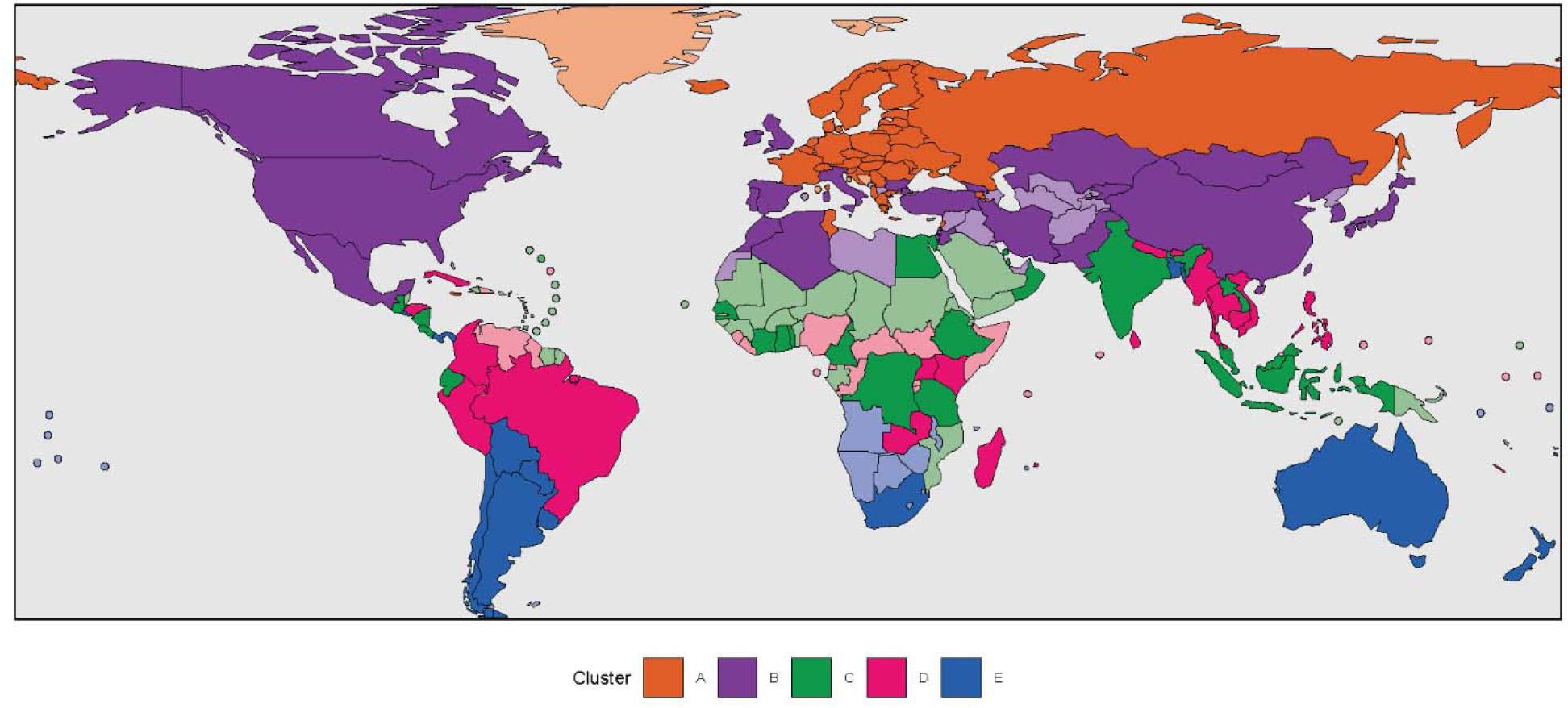
Global map of countries clustering into five influenza epidemic zones based on patterns of influenza virus detection from routine surveillance, 2011–2024. Epidemic zones are differentiated by color. Countries with darker shading were included in the primary clustering analysis, while countries with lighter shading had their epidemic zone inferred based on their population-weighted mean latitude.

Thirty-four countries were assigned to epidemic zone A from the primary clustering analysis, with all countries geographically located in the NH; 29 countries (85%) were in Europe (**Figure 3, Supplementary Table 5**). Modeling of the influenza burden of the countries in this epidemic zone showed that most influenza activity (98%) occurred during weeks 47–21, corresponding to NH-like seasonality during November–May (**Figure 4, Supplementary Table 6**). Individual country models aligned with this timing, with some variation (**Supplementary Figure 4**). Thirteen countries with insufficient or no influenza detection data were also assigned to epidemic zone A based on latitude, with most (85%) in Europe.

**Figure 4:**
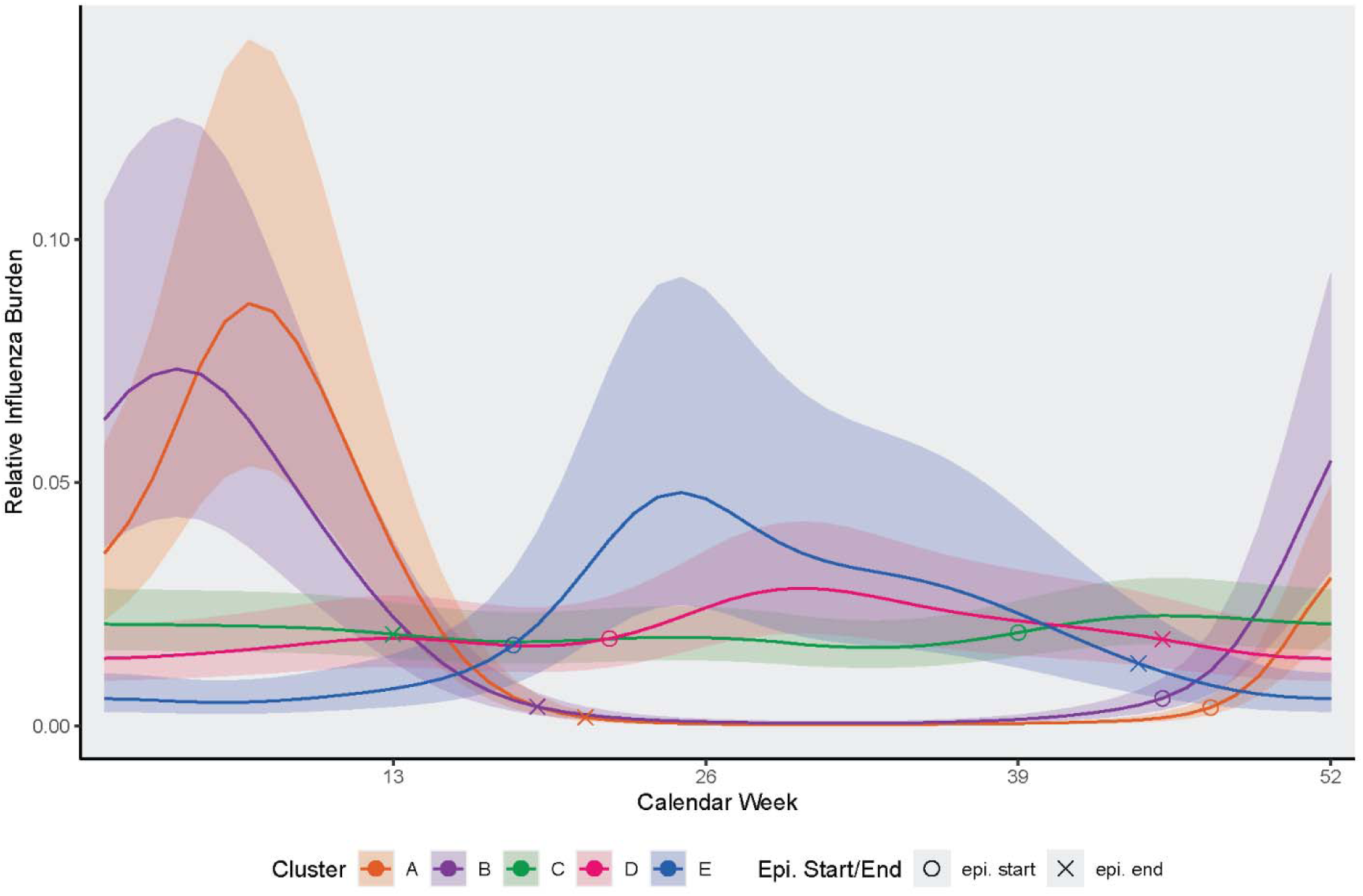
Generalized additive curves modeling the influenza detection pattern typical for each epidemic zone with 95% confidence intervals. Summary seasonal curves were calculated for each zone using hierarchical GAMs from country-year influenza burden. The main influenza epidemic for each cluster is identified through the season start (open circle; first week in which the weekly relative influenza burden exceeded the median of the weekly relative influenza burden values for ≥3 weeks) and the end (x; first week in which the burden remained below the median for ≥3 weeks).

There were 24 countries assigned to epidemic zone B; 12 (50%) were in Asia, seven (29%) were in Europe, three (13%) were in the Americas, and two (8%) were in Africa. All countries were geographically in the NH. Most influenza activity (97%) for this zone occurred during weeks 45–19, corresponding to NH-like seasonality during November–May. Twenty-five additional countries were assigned to epidemic zone B according to their population-weighted mean latitude, with more than half of countries (52%) in Asia.

Twenty-two countries were assigned to epidemic zone C, including nine (41%) in Asia, nine (41%) in Africa, and four (18%) in the Americas. More than half of influenza activity (57%) for this zone occurred during weeks 39–13, corresponding to September–March. Nineteen (86%) countries that clustered into epidemic zone C were in the tropics (with population-weighted mean latitude between 23.5° north and 23.5° south). An additional 49 countries were assigned to zone C by latitudinal proximity, with most (96%) geographically located in the tropics and nearly half (49%) in Central America and the Caribbean.

Nineteen countries were assigned to epidemic zone D, including eight (42%) countries in Asia, five (26%) in the Americas, five (26%) in Africa, and one (5%) in Oceania. Seven (37%) countries were geographically in the SH. This epidemic zone was characterized by mostly SH-like seasonality, with more than half of influenza activity (55%) occurring during weeks 22–45, corresponding to June–November, but with activity detected in other months of the year as well. An additional 22 countries were assigned to zone D based on latitude, with more than half (55%) in Africa.

Twelve countries clustered into epidemic zone E, with seven (58%) countries in the Americas, three (25%) in Oceania, one (8%) in Asia, and one (8%) in Africa. Nine (75%) countries were geographically in the SH. This epidemic zone was characterized by SH-like seasonality, with most influenza activity (82%) occurring during weeks 18–44, corresponding to May–November. Thirty-three countries, all geographically in the SH, were additionally assigned to zone E based on latitude, with most countries in Oceania (46%) and Africa (37%).

Each influenza epidemic zone defined in this analysis included countries across multiple influenza transmission zones^9^ (**Supplementary Figure 5**). While countries in 56% of the 18 transmission zones were divided across ≤2 epidemic zones, there was substantial split of some transmission zones across epidemic zones; countries in the Southern Asia and Central America/Caribbean transmission zones were distributed across four and five epidemic zones, respectively.

## DISCUSSION

Influenza viruses circulate with periodicity in most countries around the world, and these patterns can be used to guide influenza surveillance, prevention, and control decisions. However, the influenza transmission zones currently in use focused on geographic proximity and applied crude measures to summarize when influenza activity occurs in countries. We aimed to improve upon these efforts, using influenza surveillance data from 111 countries to statistically describe country-specific patterns and then parsimoniously consolidate those patterns into influenza epidemic zones. We used data from WHO’s FluNet database, which collates influenza detection data from 180 contributing countries, maximizing the global coverage of our analyses. We found that these epidemic zones were strongly associated with latitude and, therefore, leveraged this association to predict the epidemic zone for countries that did not have sufficient influenza surveillance data. The result is a global clustering of all countries into five epidemic zones that are defined by the similarity of influenza virus detection patterns throughout the year. We propose these epidemic zones as an updated framework for classifying global influenza trends.^6^

Historically, two seasonal patterns, reflecting influenza circulation in the temperate regions of the northern and southern hemispheres, have been described. Unfortunately, the seasonal pattern in a country has often been assumed based on whether the country is geographically in the northern or southern hemisphere without considering local influenza circulation. We observed patterns typical of temperate regions in our epidemic zones A and B, which include mostly countries in the NH that tended to have more influenza detections during colder months from November–May, and in zone E, which includes mostly countries in the SH that tended to have regular and defined seasons during May–November. It has also long been recognized that this crude split does not capture the experience of countries in the tropics who detect influenza viruses year-round or have influenza epidemics at inconsistent times in the calendar year.^3,4^ Countries in the tropics primarily clustered into epidemic zones C and D in our analysis, and, accordingly, had influenza activity characterized by multiple seasonal peaks, lengthy periods of detection, or seasonal timing outside the typical NH and SH months.

After the 2009 influenza A(H1N1) pandemic, surveillance for influenza improved in many countries, which spurred multiple efforts to characterize influenza circulation patterns globally.^2–4,6,21^ The objective of these efforts was to help countries discern when influenza epidemic activity typically starts so that the country could choose the most appropriate influenza vaccine formulation for their population and better time vaccine and antiviral procurement and use. These efforts were jointly summarized by WHO that resulted in 18 influenza “transmission zones,” largely based on administrative zones, geography, when influenza epidemics typically started, and how many epidemic peaks occurred, on average.^4^ Our epidemic zone analysis clustered across some but split up other transmission zones. Countries in the European transmission zones (Northern, Eastern, and Southwest Europe), Central and Eastern Asia, North and Temperate South America, and Southern Africa grouped into one or two epidemic zones. However, the Southern Asia and Central America/Caribbean transmission zones were substantially split up across epidemic zones in our analysis. This dis-array of the transmission zones in our epidemic zones highlights the original emphasis on geographic adjacency to assign countries into influenza transmission zones and the diversity in influenza detection patterns in relatively small geographic areas, particularly in the tropics.

Our analyses optimizing the number of clusters highlight that influenza detection data from countries best resolve into five clusters. We implemented an ensemble consensus clustering approach to statistically group countries based on their influenza detection data, which differs from previous regional and global analyses that used k-means and hierarchical clustering approaches (including complete and average linkage) without ensembling.^7,22^ Greater cluster resolution into groupings with fewer countries at the regional level would likely require additional data sources, such as climate, population mobility, and influenza sequencing data, though current sequencing data are less representative than influenza surveillance data and may introduce bias due to heterogeneous data availability across regions.

The diversity in influenza circulation patterns within geographically similar countries has been described for the African continent.^5^ Igboh, et al. examined a subset of the surveillance data (2010 2019) used in our analysis and described four general patterns of activity in Africa that they aligned with influenza vaccine formulations, either northern or southern hemisphere. In that analysis, the authors found neighboring countries with patterns of influenza detection that were different enough to align with opposing vaccine formulations. For example, data suggested that influenza viruses circulate in Burkina Faso in a pattern aligned with NH activity, while in Togo the pattern was mostly aligned with the northern but occasionally with the southern hemisphere, and detection patterns in Ghana aligned with SH activity. This diversity, seen in Africa but also in other areas like Central America, could reflect true and distinct influenza circulation patterns, informed by a myriad of factors such as population movement or climate factors.^23^ It could also reflect the limitations of surveillance data and the characteristics of influenza detection examined when generating clusters. Thus, as with prior efforts, our analysis and the five epidemic zones we describe may serve as useful but imperfect representations of average influenza activity by country.

In our analysis, we observed diversity in influenza patterns across seasons, with countries shifting to different clusters depending on the influenza season. In particular, epidemic zone B had lower cluster stability, with several countries with robust surveillance data shifting in assignment across epidemic zones A–C depending on the season, resulting in a lower consensus score (48%). These findings may reflect true differences in influenza seasonality and timing, potentially due to climate^24^ and differences in circulating influenza virus subtypes,^25^ and they highlight the importance of regularly revisiting global patterns in influenza seasonality. Our analytic framework and data pipeline is set up to enable ingestion of more recent data to reanalyze and consolidate influenza surveillance data into epidemic clusters.

Given the well-described relationship between latitude and influenza seasonality,^2,3,21,26–29^ we assessed the validity of assigning the 142 countries with limited or no influenza detection data available for the primary clustering analysis to epidemic zones based on the proximity of their latitude to the mean latitude of the epidemic zone. This approach classified, on average, 77% of countries with sufficient data for inclusion in the clustering analysis to their correct cluster when they were synthetically left out of the analysis, suggesting that this method can be leveraged in the context of our analyses to assign countries to epidemic zones even if they do not yet have sufficient data to be formally incorporated. By drawing on seasonal patterns observed in countries within the same cluster, this approach provides a basis for identifying plausible circulation patterns where country-specific data are sparse for regional and national public health decisionmakers to use for epidemic readiness and planning. These latitude-based assignments should be interpreted with caution as estimates of potential seasonality; nonetheless, this approach offers a practical way to extend global health surveillance and its broader public health relevance. Periodic reassessment of epidemic zones from more recent data may also permit inclusion of these countries in the future.

As a means for data aggregation and summary, epidemic zones may be useful for optimizing the use of influenza surveillance data for decision-making. For example, they may serve as a data-driven unit of analysis for vaccine strain composition discussions; consultation experts could evaluate data that have good coverage of influenza surveillance and virus information by epidemic zone when it is challenging to get enough data by country. Public health officials may see benefits from aggregating influenza surveillance and situational awareness by epidemic zone rather than country when country-specific data may be too sparse, for example, when developing seasonal scenario models or short-term forecasts.

In conjunction with more granular evaluation of national and subnational influenza trends, health officials may use these new epidemic zones to anticipate the timing of influenza activity in their countries and plan interventions for epidemic readiness. Countries could use epidemic zones and similar findings to better time communications with healthcare facilities on when to engage in enhanced surveillance; anticipate the use of medical countermeasures (such as influenza antivirals); and plan for surge timing for hospital beds, oxygen, and staffing. While public health officials should use country-specific influenza trends for precise operational decision-making, they may also use these zones to time the development, testing, and launch of risk communication campaigns and inform availability of prevention and control efforts. Finally, public health agencies can also leverage these influenza epidemic zones for global health security, monitoring for when influenza activity is predicted to be heightened in countries and developing communication and response efforts, such as guiding vaccination recommendations, for citizens travelling or stationed abroad.

Our analysis is limited in several ways. First, despite including data through 2024, our findings are mostly based on influenza surveillance data collected prior to the COVID-19 pandemic. We deliberately excluded data from the early pandemic years due to the significant disruptions in influenza detection caused by pandemic mitigation measures and SARS-CoV-2 transmission dynamics.^23,30–35^ Future analyses should incorporate additional data from ongoing surveillance to assess potential changes in seasonal timing and their implications for the epidemic zones we have identified. Second, we used publicly available influenza surveillance data submitted by countries to WHO FluNet. While a standardized approach to influenza surveillance is recommended, the methodologies employed for virologic surveillance differ across countries.

Some countries conduct systematic testing of patients meeting syndromic case definitions at sentinel sites, while other surveillance systems combine systematic and clinical testing data. We tried to reduce this bias by analyzing a standardized estimate of influenza “burden” within each country prior to cross-country comparisons and clustering. Nonetheless, our clustering analysis may still be sensitive to these methodological discrepancies between countries. Third, countries report aggregated, national-level influenza virologic data to WHO; a centralized dataset of subnational-level data with comparable coverage to FluNet is not available. However, certain countries, such as Brazil, Peru, Kenya, India, and China, encompass multiple climate zones that likely exhibit distinct subregional patterns of influenza circulation. Consequently, national summary trends and cluster assignments may not accurately reflect these subregional dynamics. Future efforts to disaggregate influenza surveillance data by subregion would enhance our understanding of these differences and could inform more effective influenza prevention and control strategies. Fourth, our analysis, like prior efforts, struggled with insufficient or limited data from many countries to robustly define when influenza likely circulates.^3,4^ Further improvements to influenza surveillance would strengthen global situational awareness and these types of analyses.

Future research could explore whether the influenza epidemic zones we have delineated would be altered or corroborated by incorporating additional variables, such as population dynamics, climate factors, or geographic features, known to influence influenza circulation patterns.^3,21,36,37^ Furthermore, we have described the detection of any influenza virus, and future work should build upon these methods to explore specific characteristics of influenza viruses, including virus type and subtype, antigenic, and genetic information that could be shaping influenza dynamics globally.

## CONCLUSION

We have applied a data-driven method to advance our understanding of influenza detection patterns around the world. By identifying the temporal and geographical patterns of influenza virus detection, our findings can inform national and global health officials about optimal timing for enhanced surveillance efforts, resource allocation, and medical countermeasures, ultimately contributing to improved influenza prevention and control measures worldwide.

## Acknowledgements

This work is the result of a collaboration during 2023–2024 within GISRS. The authors would like to thank countries and regions for submitting data to GISRS and their continued support for global influenza surveillance.

## EXTRA MATERIALS

### Contributors

PM, EA-B, NH, RJK, CSB, SH, and ST conceived the study. NH, PM, and CSB compiled, managed, cleaned, evaluated, and analyzed the data. RJK, EA-B, CSB, MAR, LMD, SH, ST, and AH provided technical expertise and feedback on data sources and analytic methodologies. PM, NH, and MAR drafted the manuscript, created the tables and figures, compiled feedback, and addressed all comments from coauthors. All authors reviewed, contributed to, and approved results, manuscript drafts, and the publication. NH, PM, and CSB contributed equally to this paper.

### Funding

There was no funding associated with this study.

### Disclaimer

The findings and conclusions in this Original Research paper are those of the authors and do not necessarily represent the official position of the Centers for Disease Control and Prevention or the World Health Organization.

### Ethics approval

This activity was reviewed by CDC, deemed not research, and was conducted consistent with applicable federal law and CDC policy (45 C.F.R. part 46.102(l)(2); 44 U.S.C. §3501 et seq).

### Data availability

The data used in this analysis are publicly available and accessible from GISRS.^13^ The analytic code for this study is available from the following GitHub repository: https://github.com/CDCgov/epidemic_zones.

