## Supplemental Materials for "Defining influenza epidemic zones through temporal clustering of global surveillance data"

### SUPPLEMENTARY MATERIALS

**Supplementary Figure 1:** Generalized additive models (GAMs) of influenza virus detection data from 2011–2024. Countries analyzed using calendar-year time windows have data shown by calendar year and weeks (weeks 1–52), while countries analyzed using shifted-year time windows have data shown by shifted year and weeks (week 30–week 29 of the following year). The raw data for influenza detections are indicated as individual gray points (crosses), while the GAMs are depicted as solid red lines.

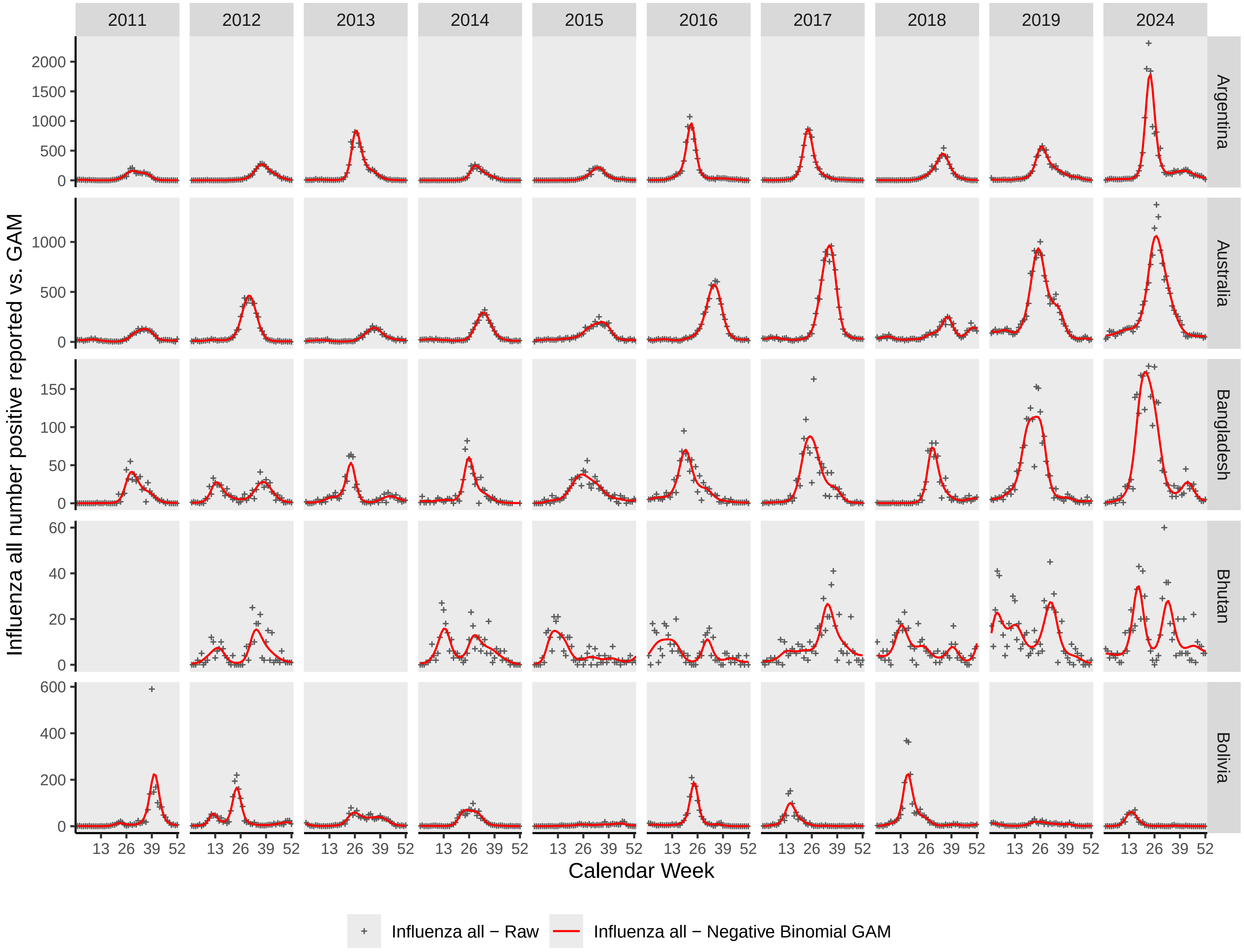

2011
2012
2013
2014
2015
2016
2017
2018
2019
2024
Argentina
Australia
Bangladesh
Bhutan
Bolivia
Influenza all number positive reported vs. GAM
Calendar Week
13 26 39 52
Influenza all − Raw
Influenza all − Negative Binomial GAM

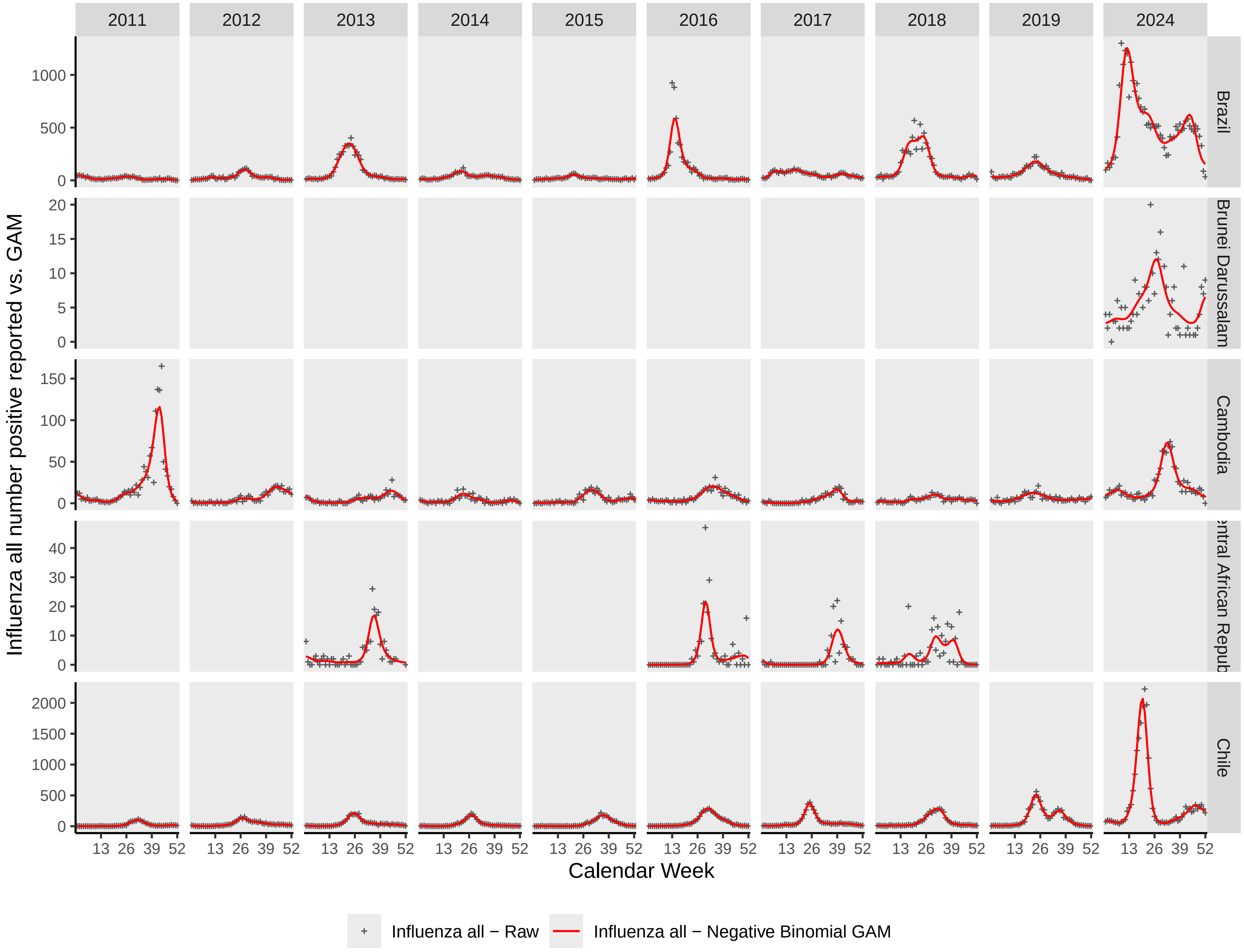
2011
2012
2013
2014
2015
2016
2017
2018
2019
2024
Brazil
Brunei Darussalam
Cambodia
Central African Repub
Chile
Influenza all number positive reported vs. GAM
Calendar Week
13 26 39 52
Influenza all − Raw
Influenza all − Negative Binomial GAM

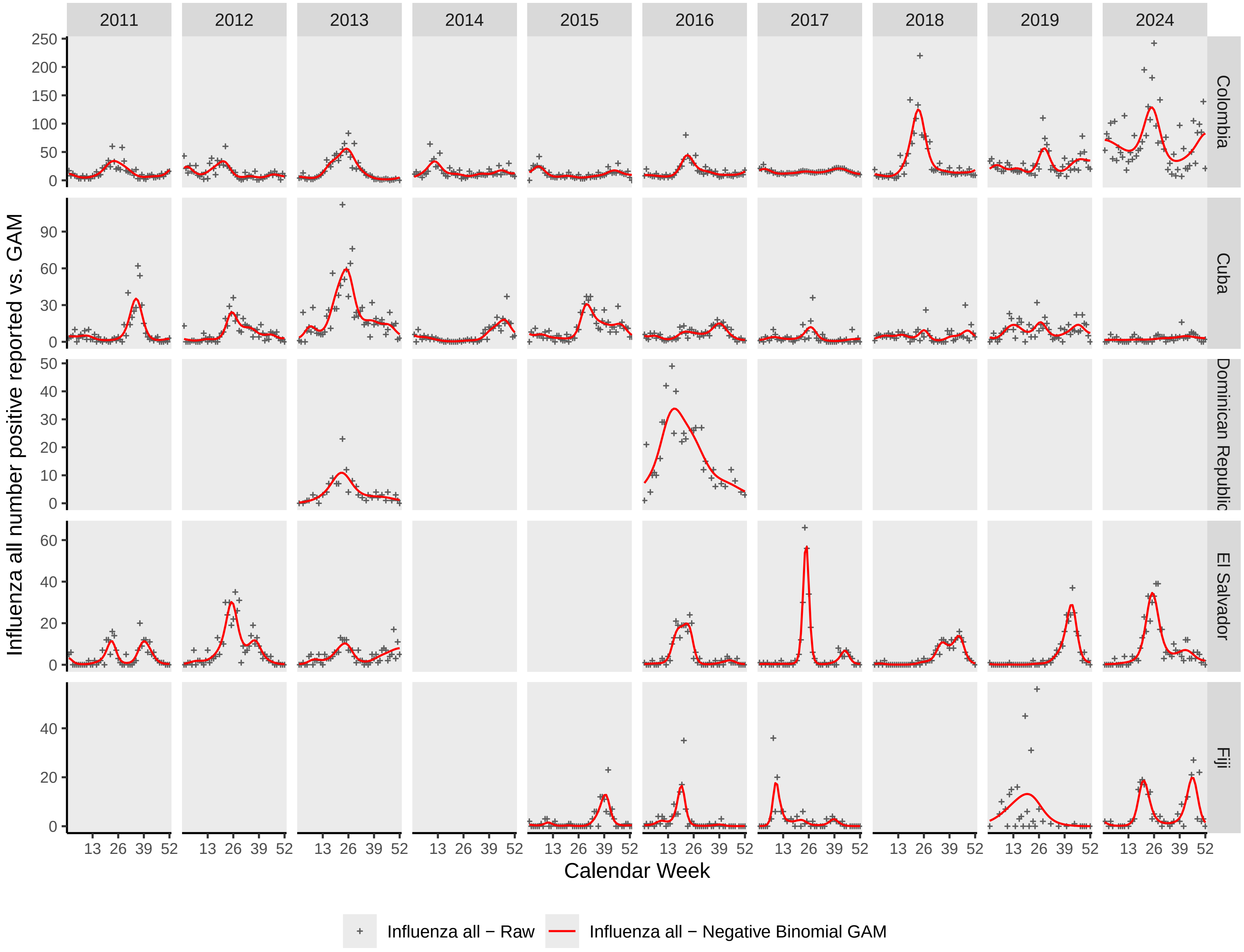

2011
2012
2013
2014
2015
2016
2017
2018
2019
2024
Colombia
Cuba
Dominican Republic
El Salvador
Fiji
Influenza all number positive reported vs. GAM
Calendar Week
13 26 39 52
Influenza all − Raw
Influenza all − Negative Binomial GAM

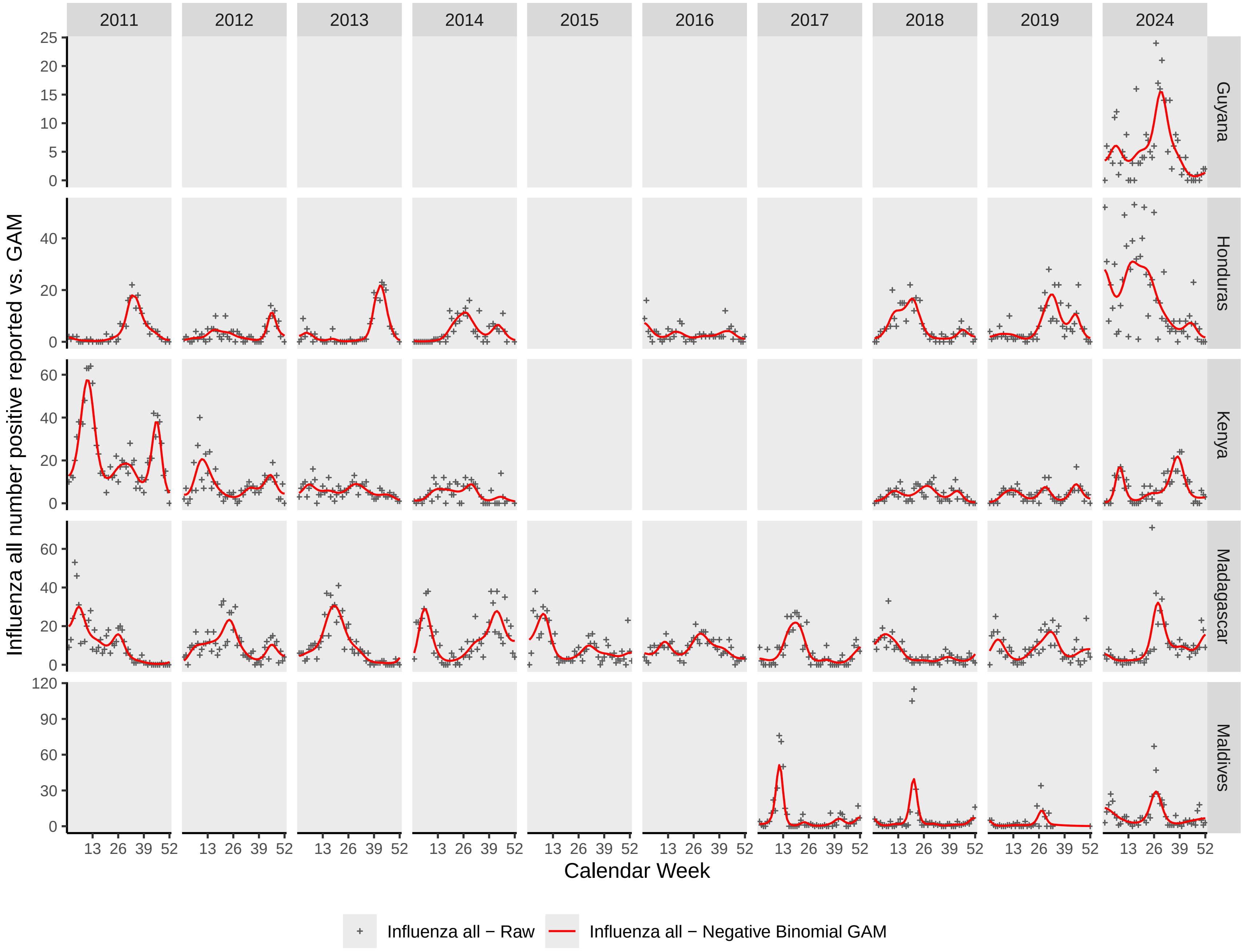

2011
2012
2013
2014
2015
2016
2017
2018
2019
2024
Guyana
Honduras
Kenya
Madagascar
Maldives
Influenza all number positive reported vs. GAM
Calendar Week
13 26 39 52
Influenza all − Raw
Influenza all − Negative Binomial GAM

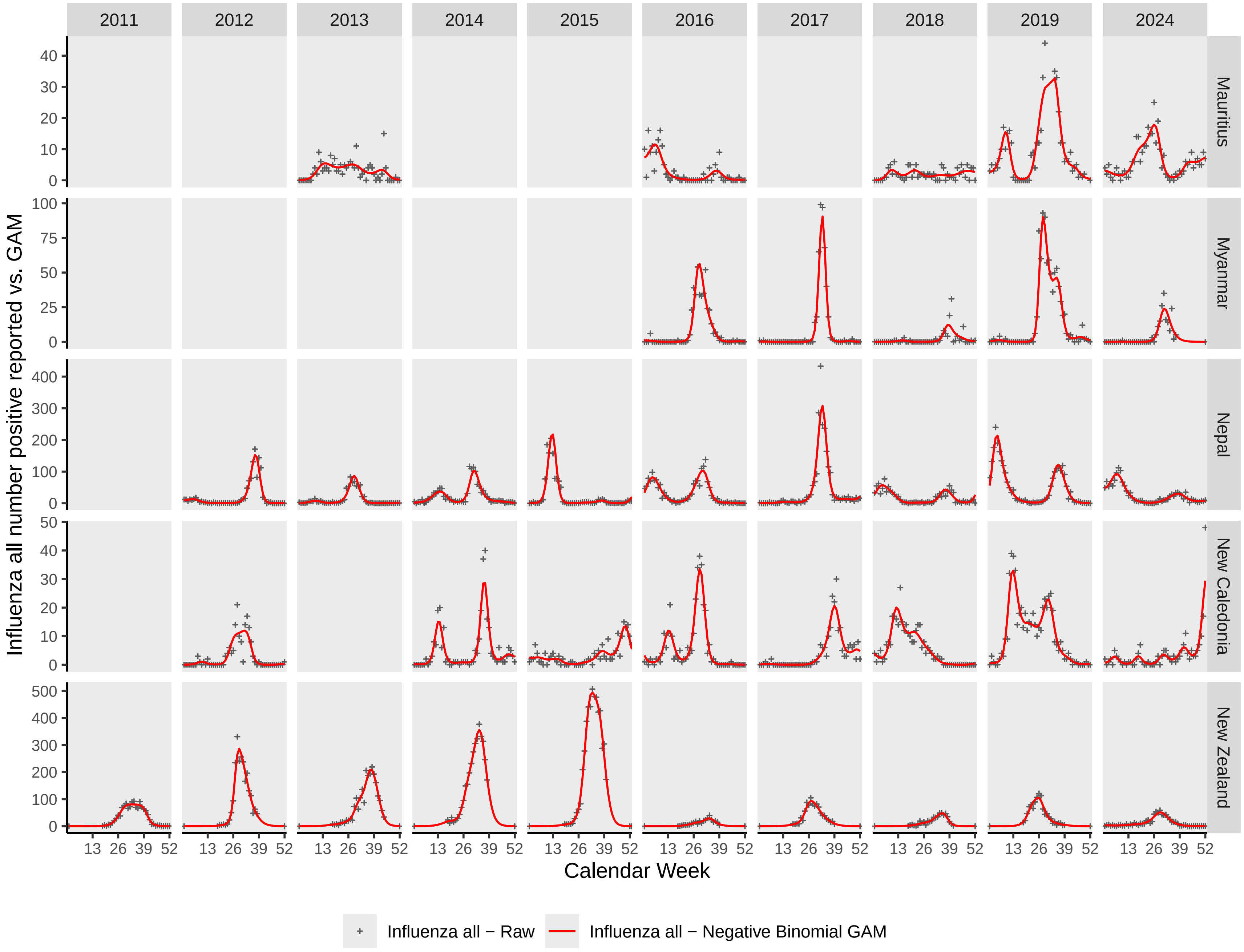

2011
2012
2013
2014
2015
2016
2017
2018
2019
2024
Mauritius
Myanmar
Nepal
New Caledonia
New Zealand
Influenza all number positive reported vs. GAM
Calendar Week
13 26 39 52
Influenza all − Raw
Influenza all − Negative Binomial GAM

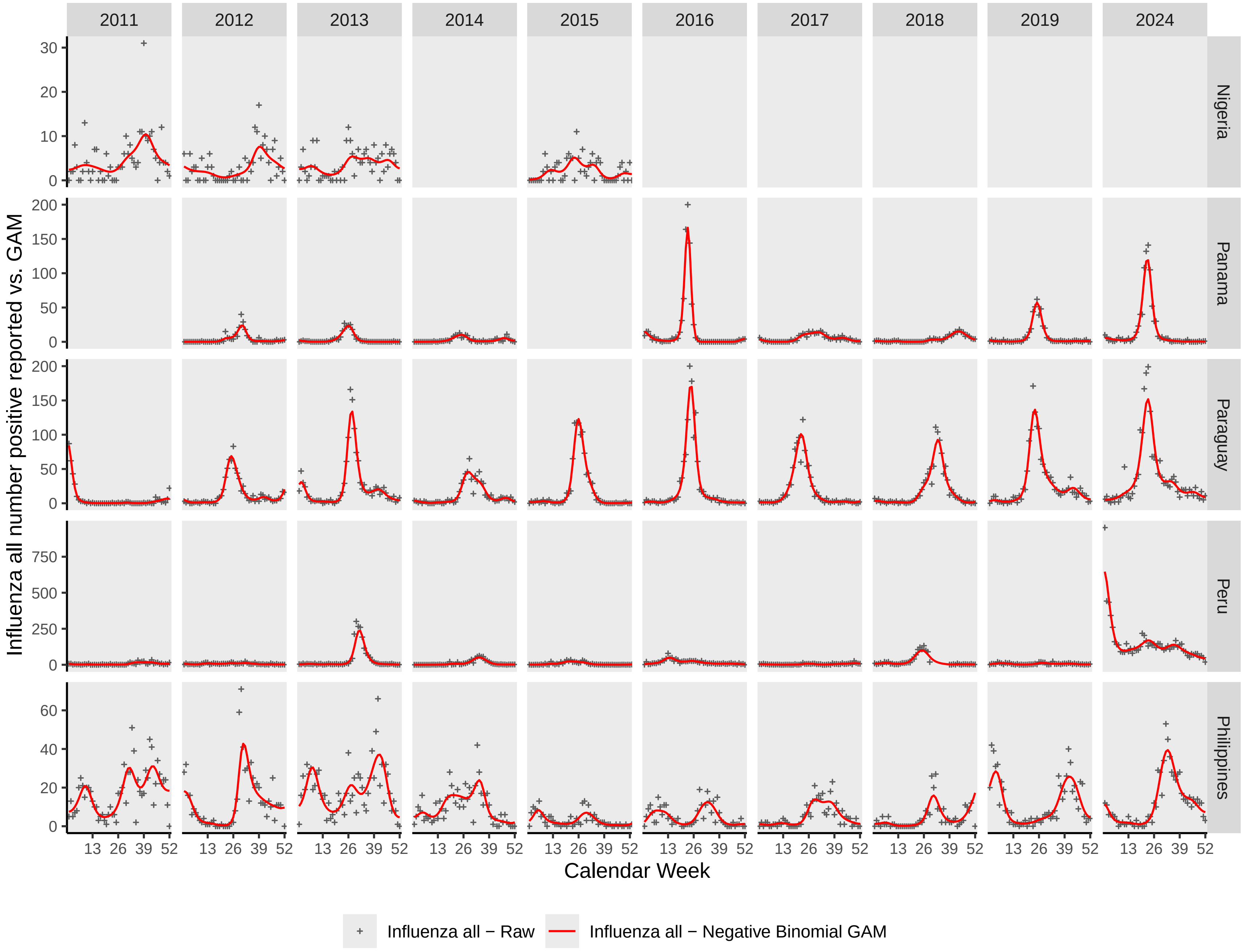

2011
2012
2013
2014
2015
2016
2017
2018
2019
2024
Nigeria
Panama
Paraguay
Peru
Philippines
Influenza all number positive reported vs. GAM
Calendar Week
13 26 39 52
Influenza all − Raw
Influenza all − Negative Binomial GAM

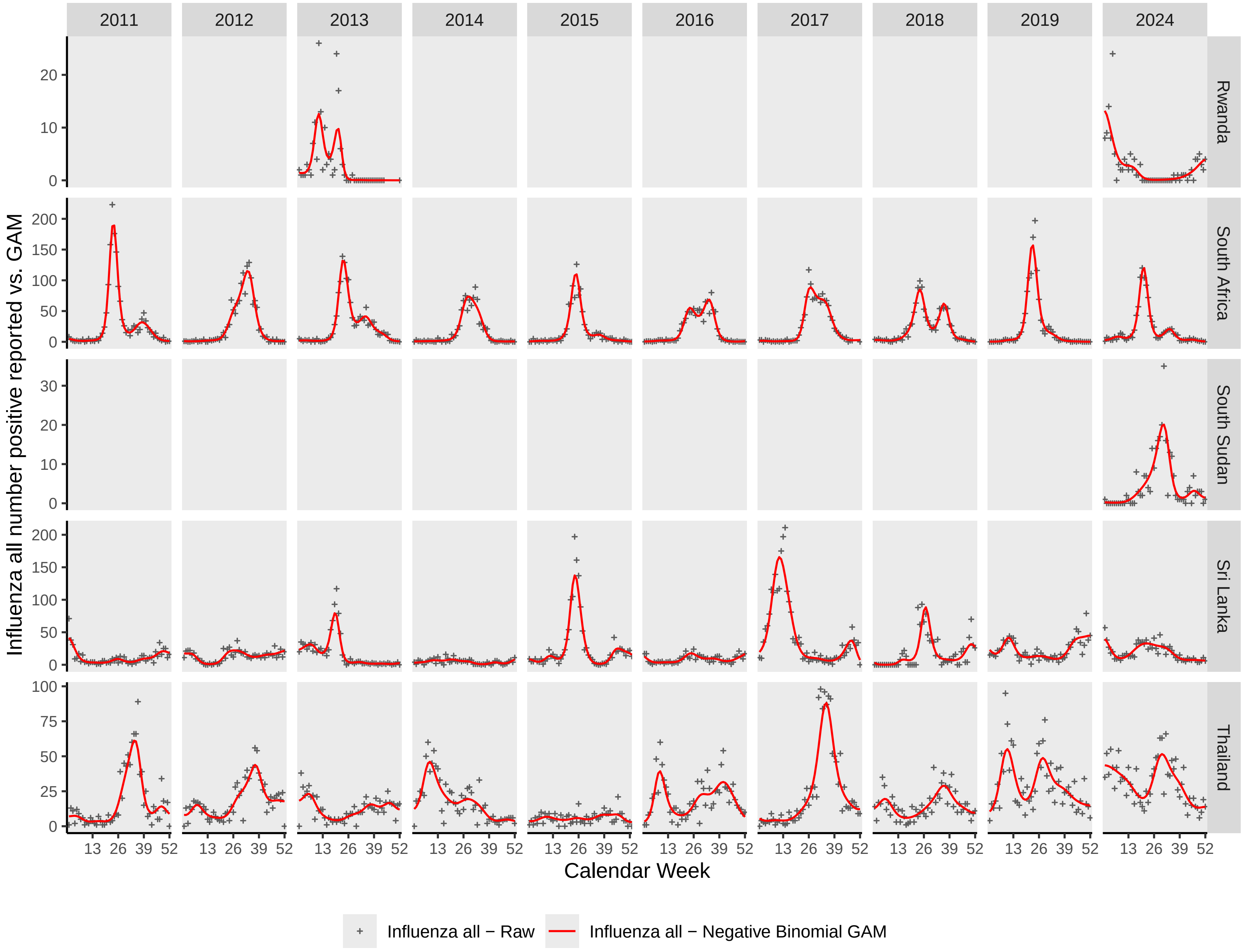

2011
2012
2013
2014
2015
2016
2017
2018
2019
2024
Rwanda
South Africa
South Sudan
Sri Lanka
Thailand
Influenza all number positive reported vs. GAM
Calendar Week
13 26 39 52
Influenza all − Raw
Influenza all − Negative Binomial GAM

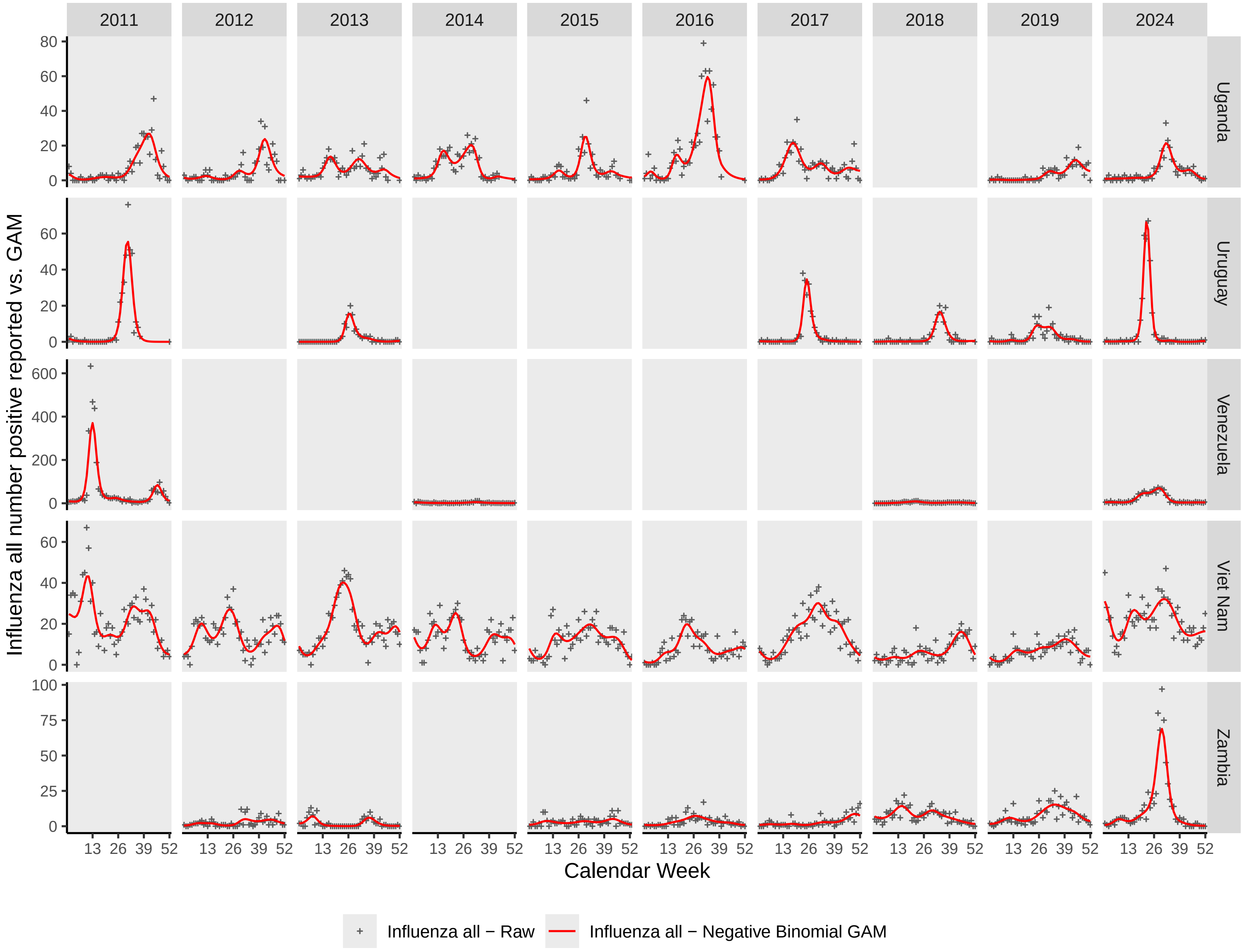
2011
2012
2013
2014
2015
2016
2017
2018
2019
2024
Uganda
Uruguay
Venezuela
Viet Nam
Zambia
Influenza all number positive reported vs. GAM
Calendar Week
13 26 39 52
Influenza all − Raw
Influenza all − Negative Binomial GAM

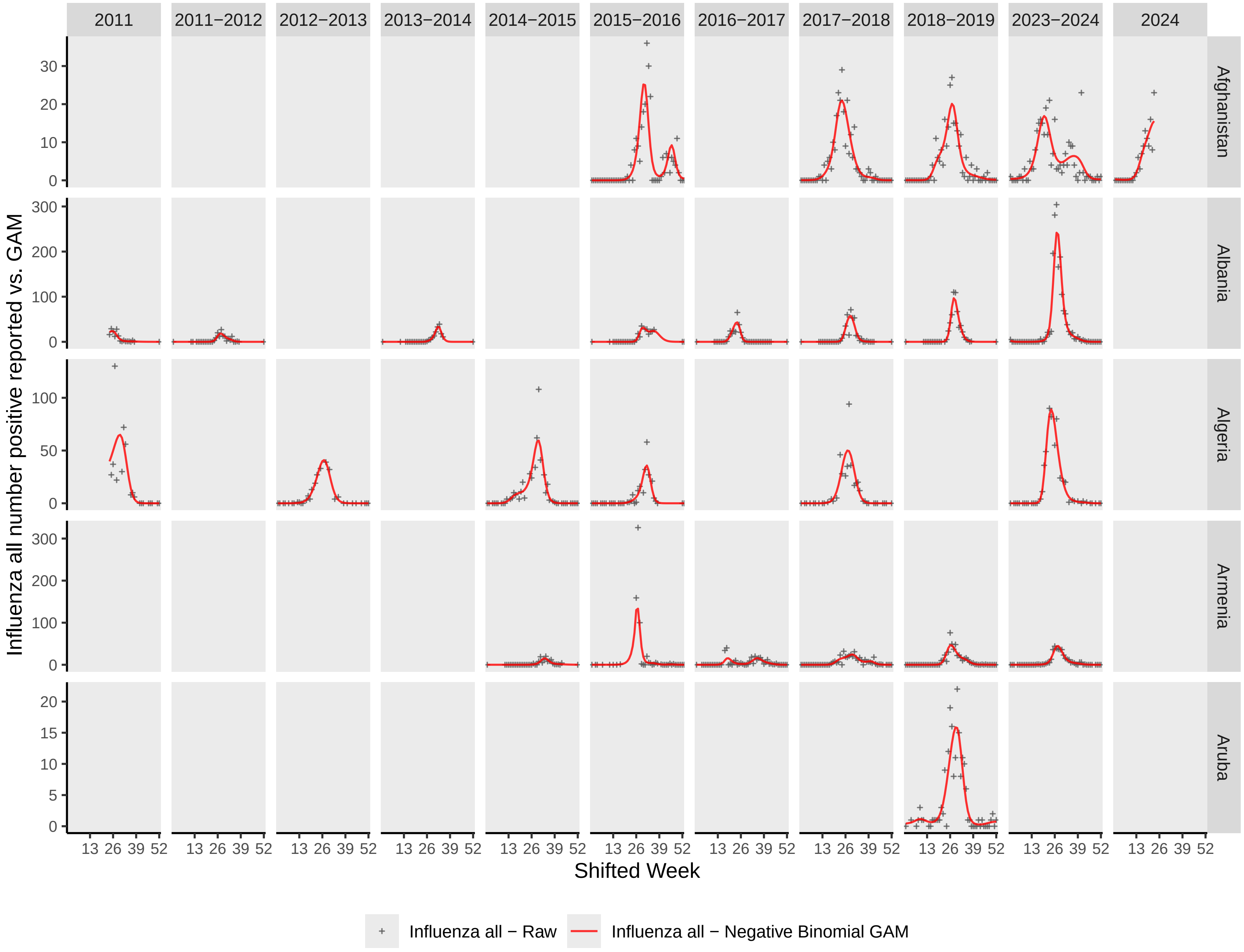

2011
2011−2012
2012−2013
2013−2014
2014−2015
2015−2016
2016−2017
2017−2018
2018−2019
2023−2024
2024
Afghanistan
Albania
Algeria
Armenia
Aruba
Influenza all number positive reported vs. GAM
Shifted Week
13 26 39 52
Influenza all − Raw
Influenza all − Negative Binomial GAM

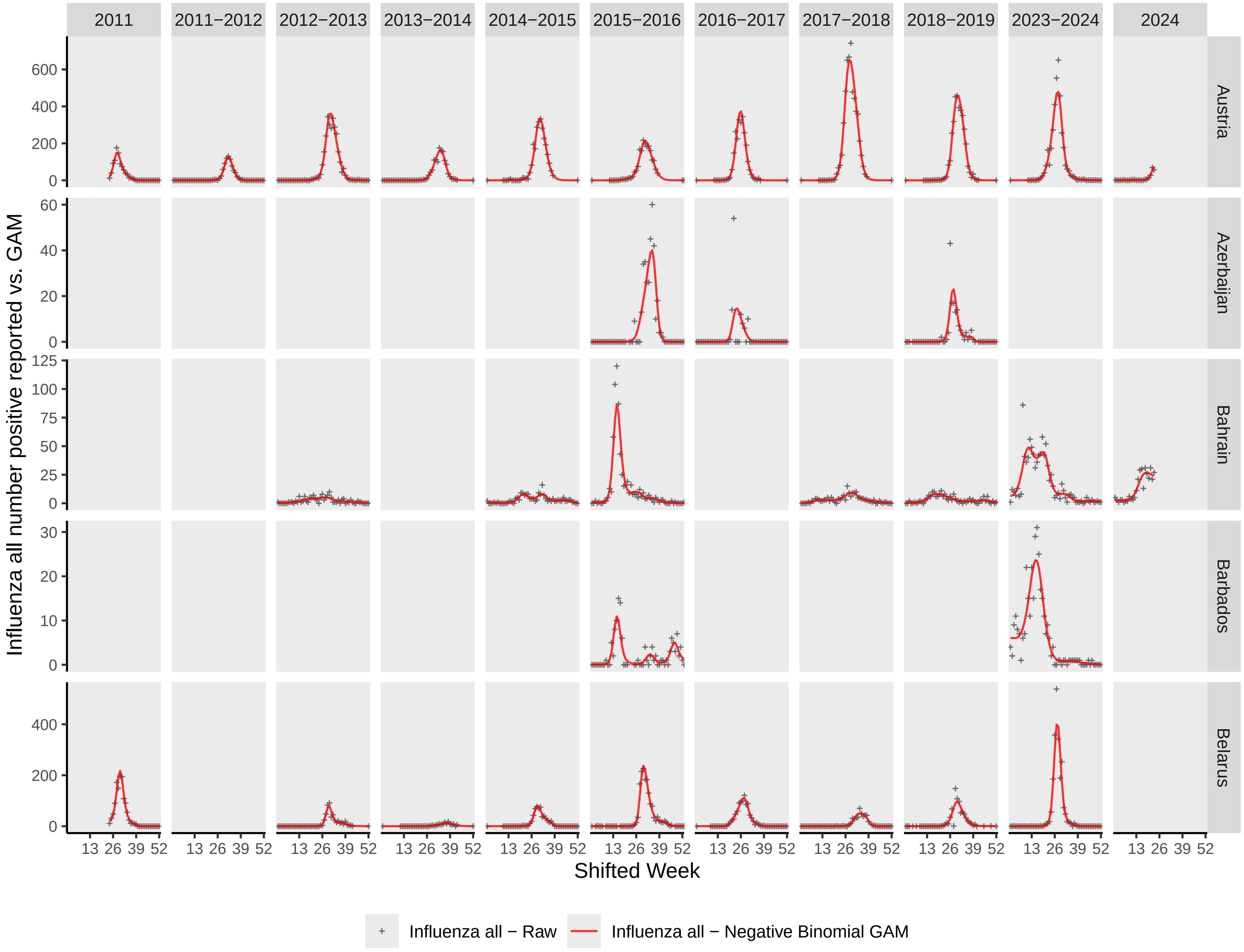

2011
2011−2012
2012−2013
2013−2014
2014−2015
2015−2016
2016−2017
2017−2018
2018−2019
2023−2024
2024
Austria
Azerbaijan
Bahrain
Barbados
Belarus
Influenza all number positive reported vs. GAM
Shifted Week
13 26 39 52
0
200
400
600
20
40
60
25
50
75
100
125
10
30
Influenza all − Raw
Influenza all − Negative Binomial GAM

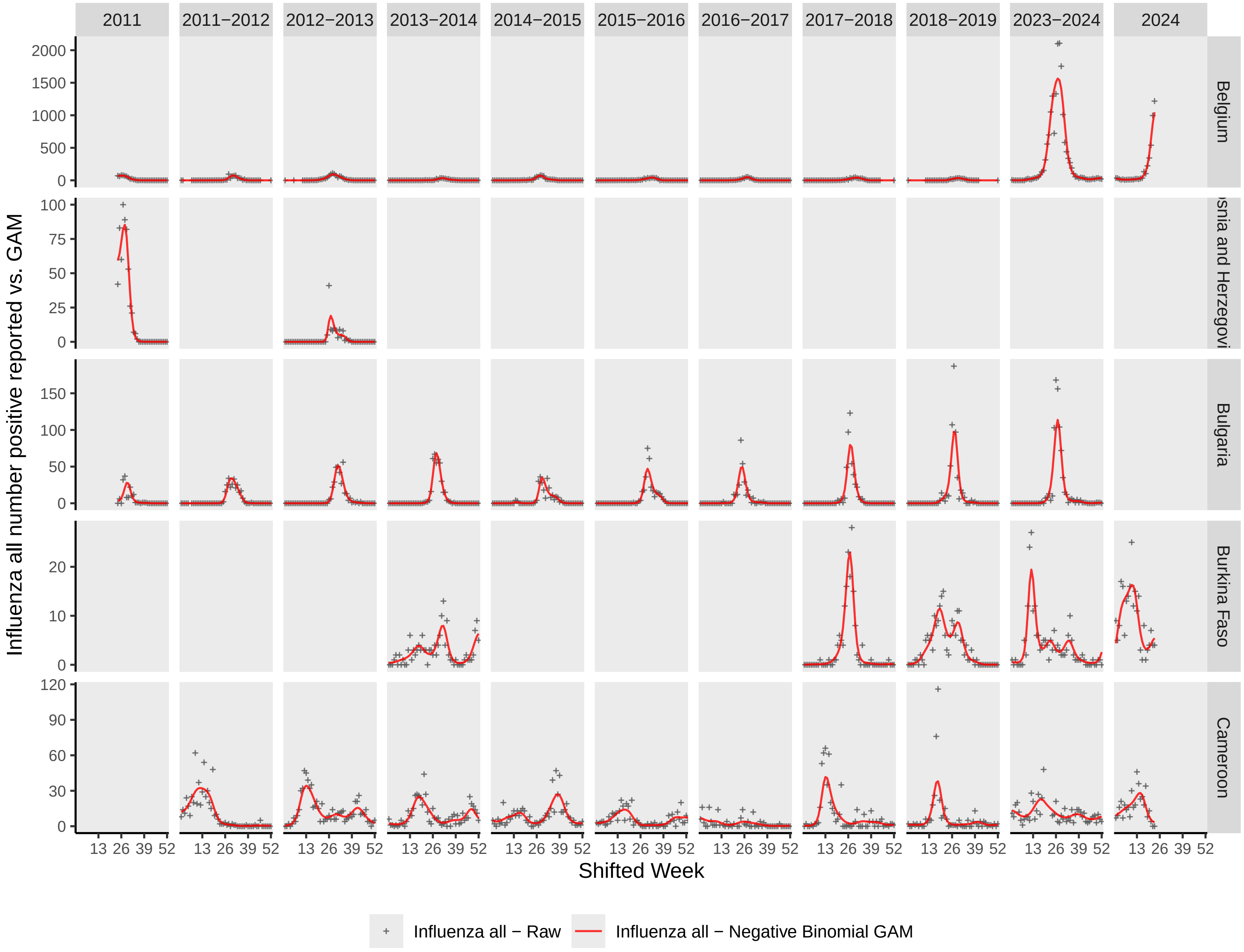

2011
2011−2012
2012−2013
2013−2014
2014−2015
2015−2016
2016−2017
2017−2018
2018−2019
2023−2024
2024
Belgium
snia and Herzegovi
Bulgaria
Burkina Faso
Cameroon
Influenza all number positive reported vs. GAM
Shifted Week
13 26 39 52
Influenza all − Raw
Influenza all − Negative Binomial GAM

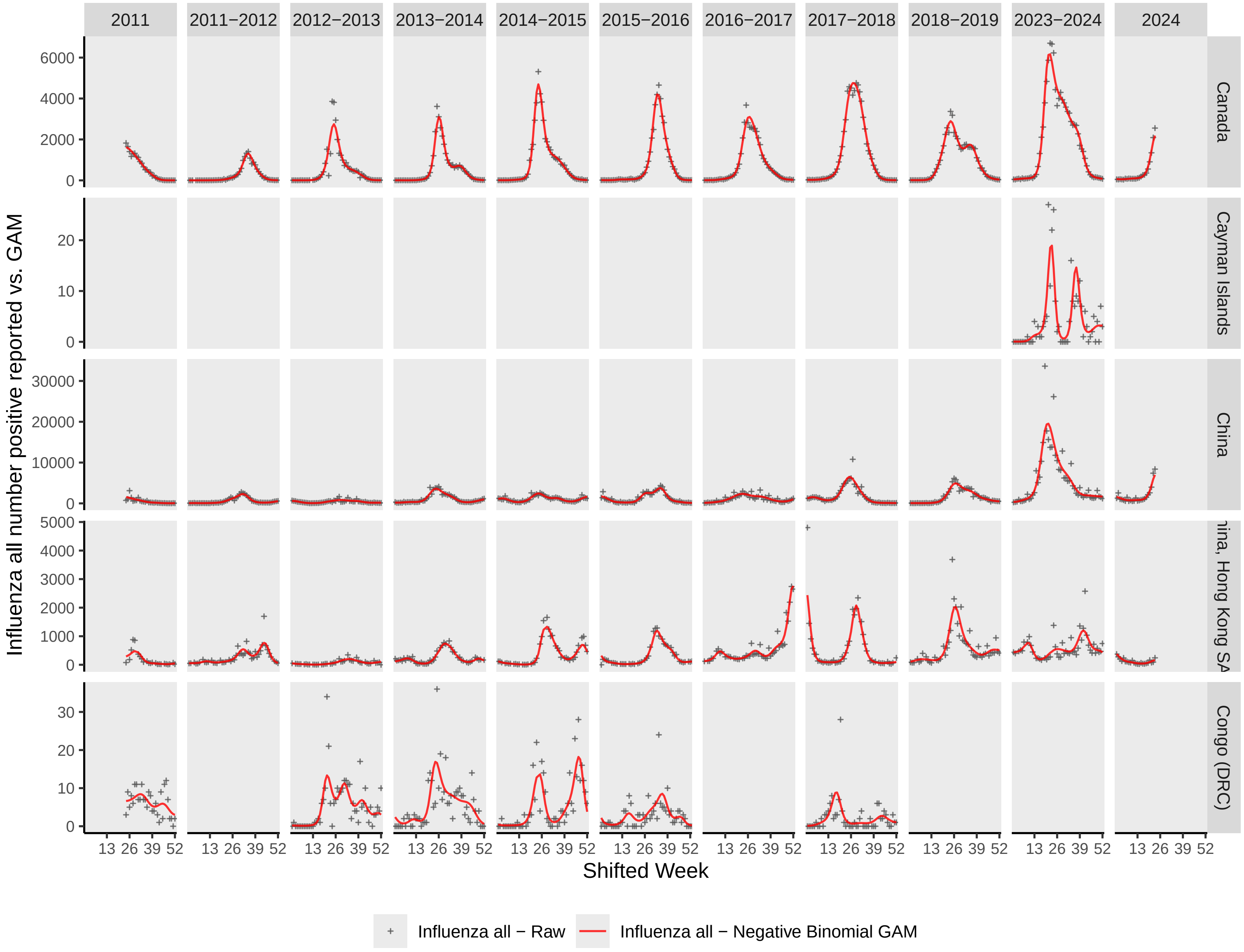
2011
2011−2012
2012−2013
2013−2014
2014−2015
2015−2016
2016−2017
2017−2018
2018−2019
2023−2024
2024
Canada
Cayman Islands
China
ina, Hong Kong SA
Congo (DRC)
Influenza all number positive reported vs. GAM
Shifted Week
Influenza all − Raw
Influenza all − Negative Binomial GAM

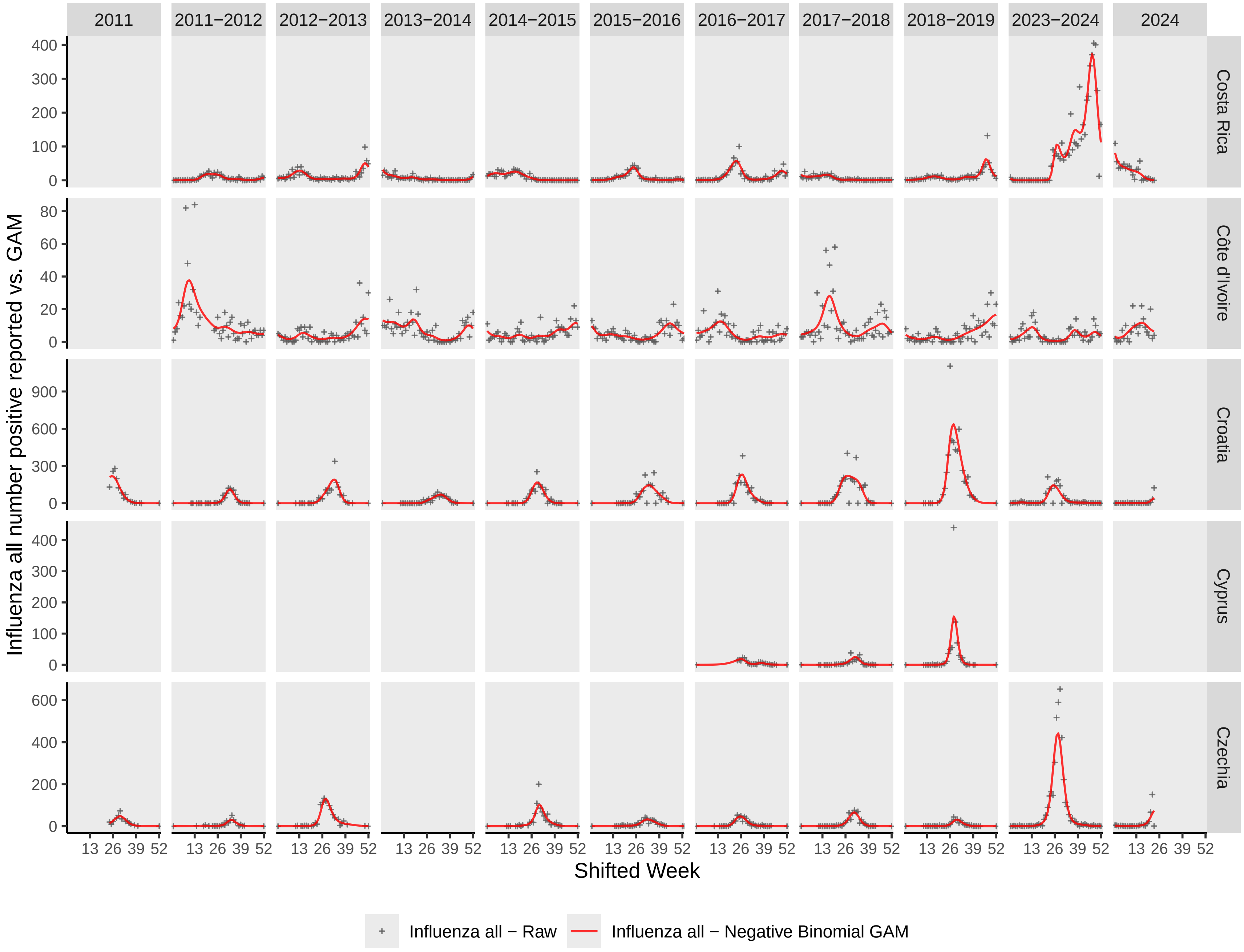

2011
2011−2012
2012−2013
2013−2014
2014−2015
2015−2016
2016−2017
2017−2018
2018−2019
2023−2024
2024
Costa Rica
Côte d'Ivoire
Croatia
Cyprus
Czechia
Influenza all number positive reported vs. GAM
Shifted Week
13 26 39 52
Influenza all − Raw
Influenza all − Negative Binomial GAM

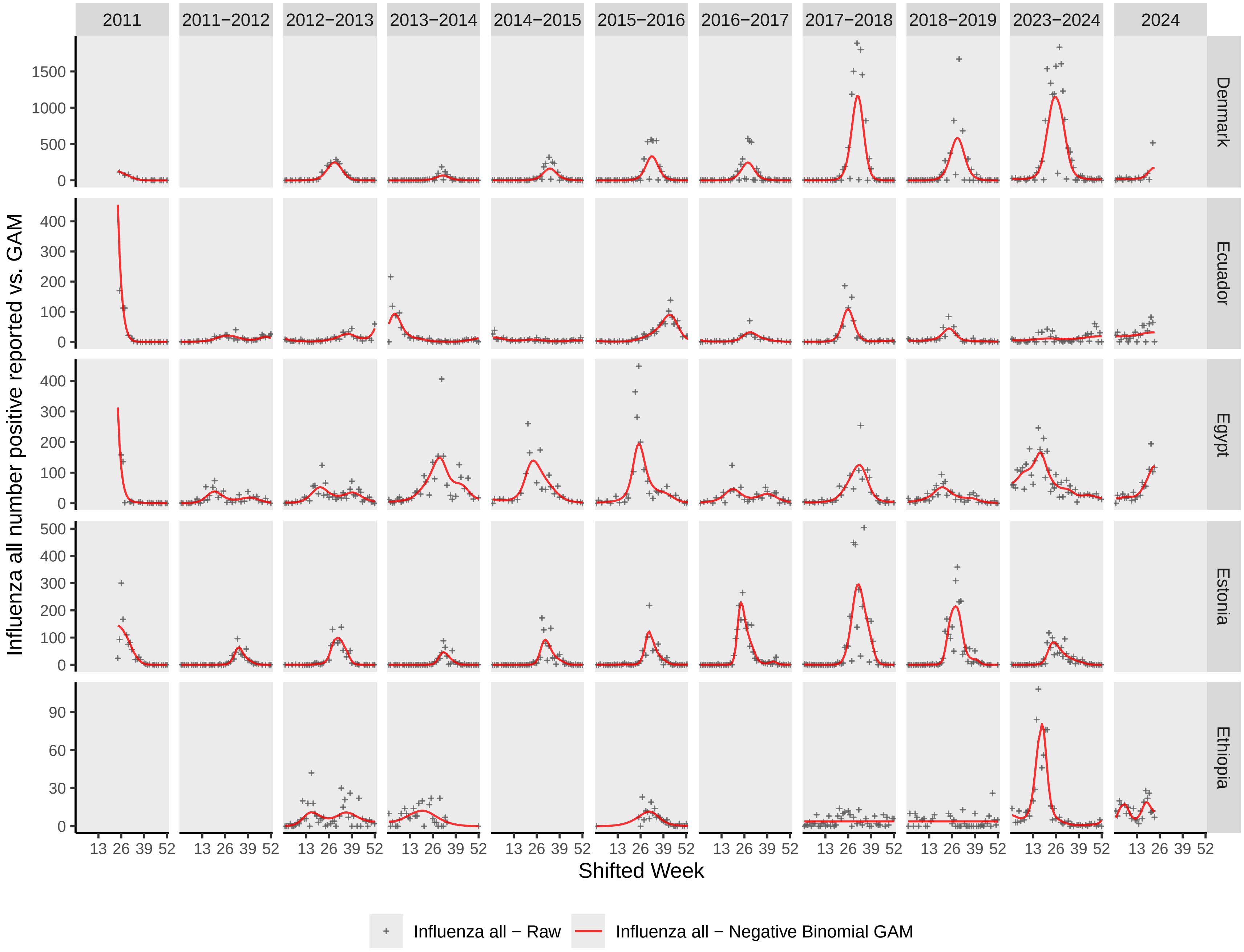
2011
2011−2012
2012−2013
2013−2014
2014−2015
2015−2016
2016−2017
2017−2018
2018−2019
2023−2024
2024
Denmark
Ecuador
Egypt
Estonia
Ethiopia
Influenza all number positive reported vs. GAM
Shifted Week
13 26 39 52
Influenza all − Raw
Influenza all − Negative Binomial GAM

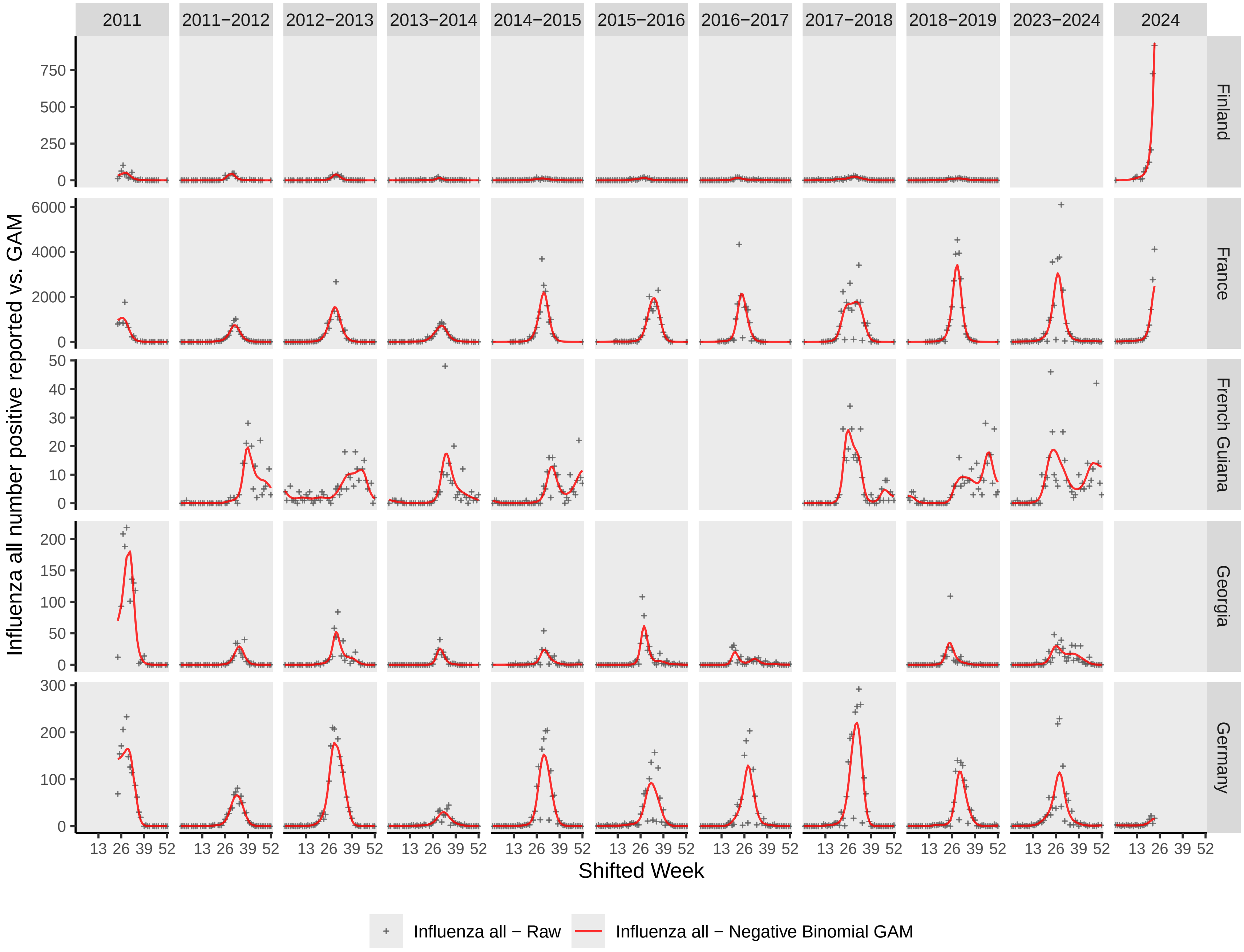
2011
2011−2012
2012−2013
2013−2014
2014−2015
2015−2016
2016−2017
2017−2018
2018−2019
2023−2024
2024
Finland
France
French Guiana
Georgia
Germany
Influenza all number positive reported vs. GAM
Shifted Week
13 26 39 52
Influenza all − Raw
Influenza all − Negative Binomial GAM

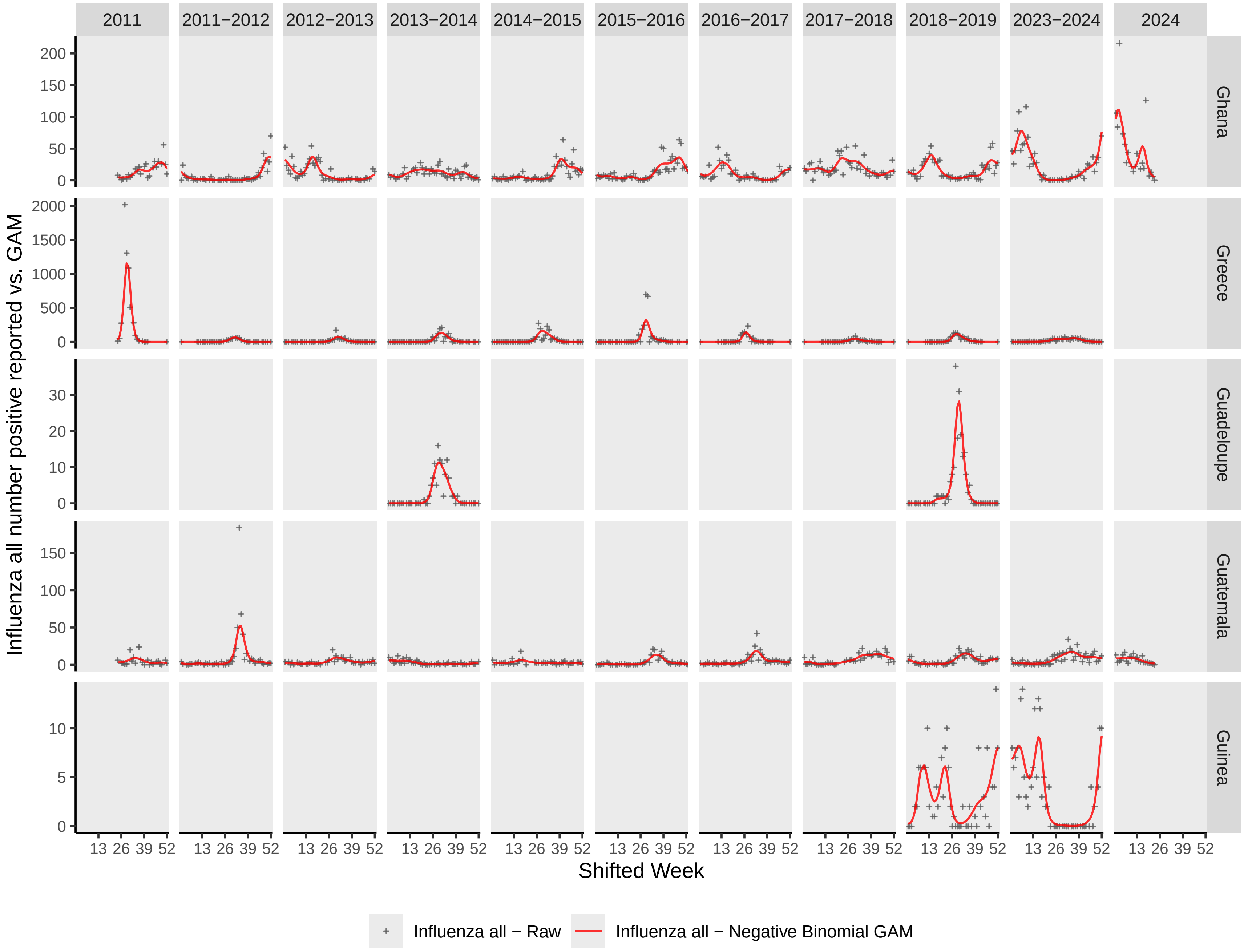

2011
2011−2012
2012−2013
2013−2014
2014−2015
2015−2016
2016−2017
2017−2018
2018−2019
2023−2024
2024
Ghana
Greece
Guadeloupe
Guatemala
Guinea
Influenza all number positive reported vs. GAM
Shifted Week
13 26 39 52
Influenza all − Raw
Influenza all − Negative Binomial GAM

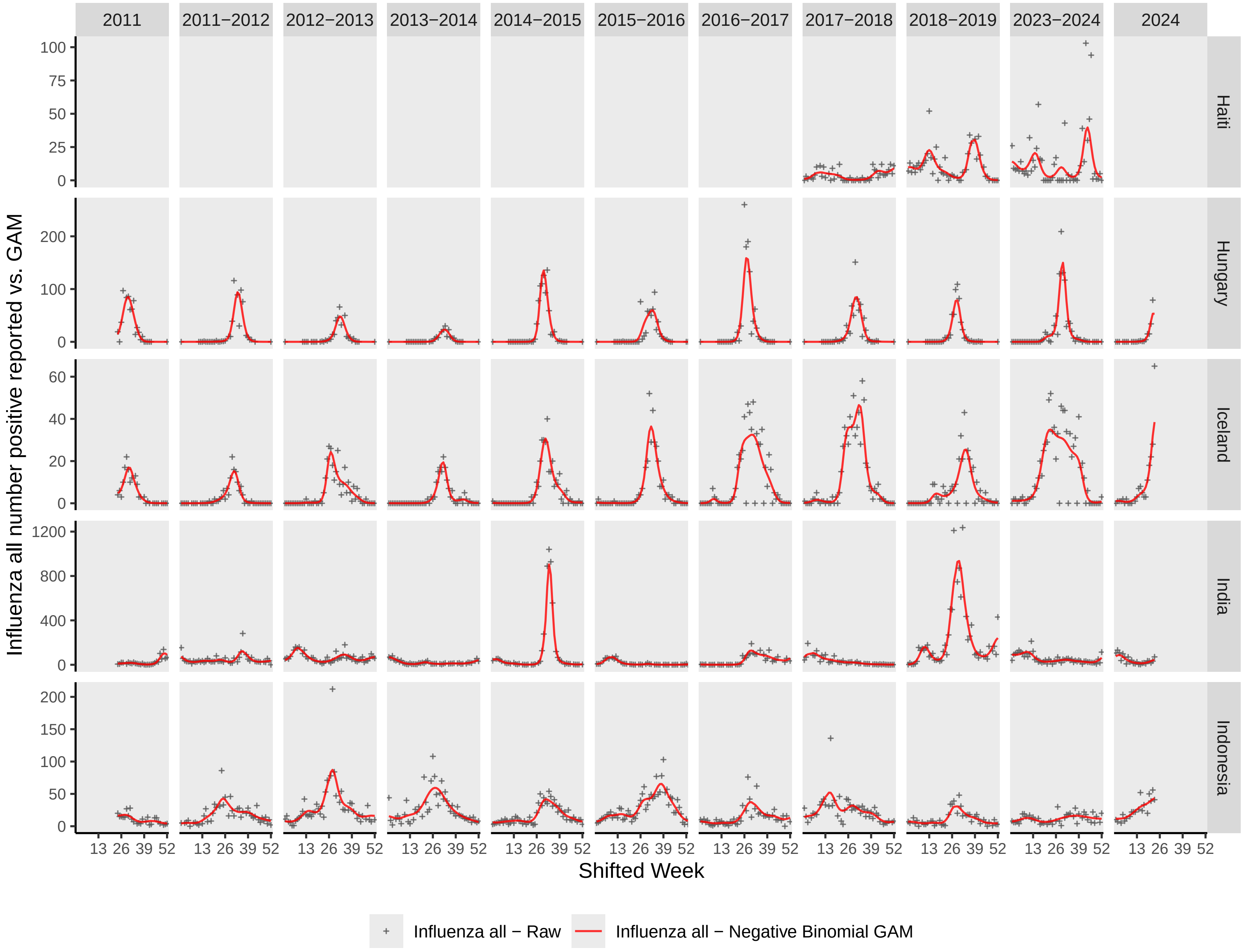

2011
2011−2012
2012−2013
2013−2014
2014−2015
2015−2016
2016−2017
2017−2018
2018−2019
2023−2024
2024
Haiti
Hungary
Iceland
India
Indonesia
Influenza all number positive reported vs. GAM
Shifted Week
13 26 39 52
0 25 50 75 100
0 100 200
0 20 40 60
0 400 800 1200
0 50 100 150 200
Influenza all − Raw
Influenza all − Negative Binomial GAM

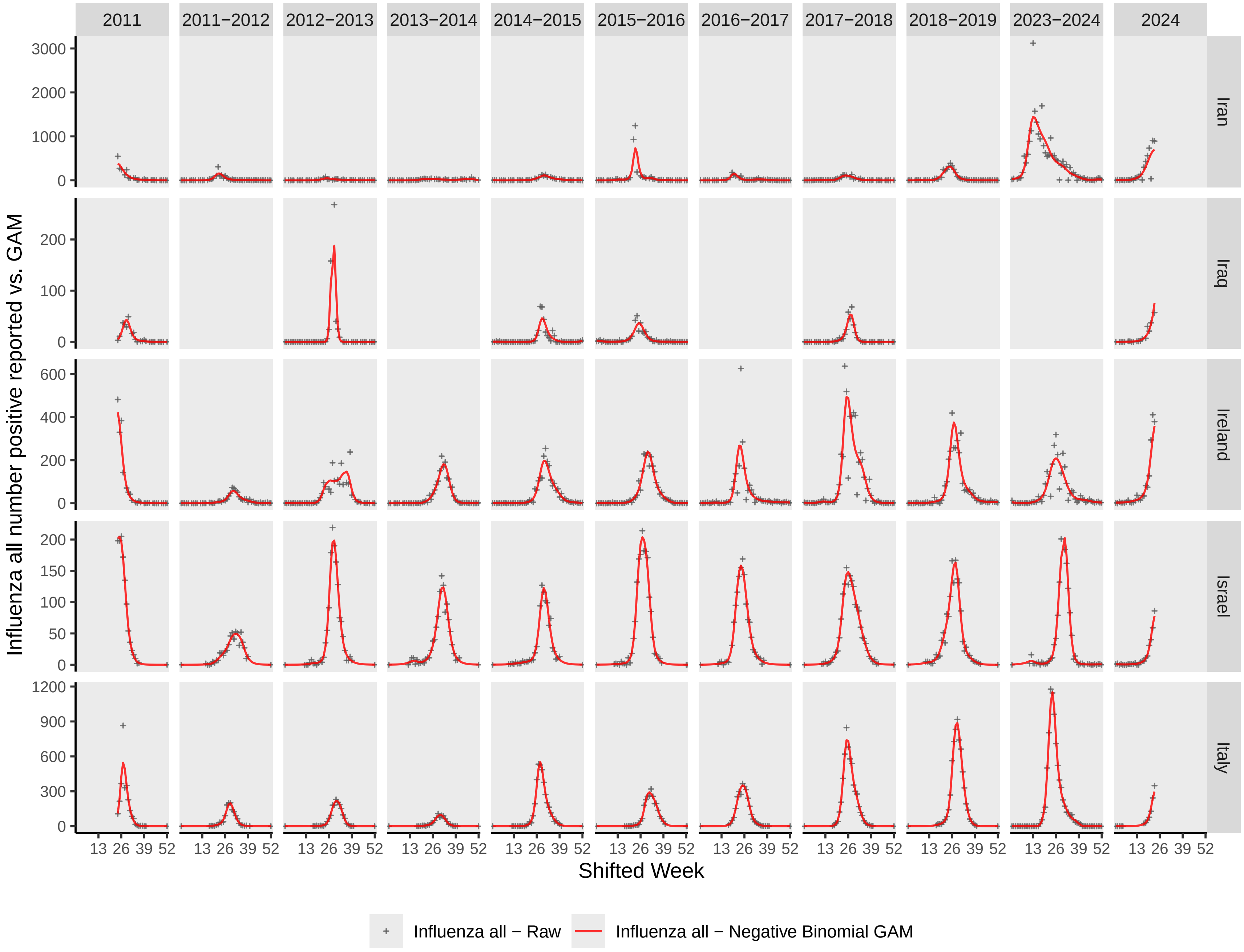

2011
2011−2012
2012−2013
2013−2014
2014−2015
2015−2016
2016−2017
2017−2018
2018−2019
2023−2024
2024
Iran
Iraq
Ireland
Israel
Italy
Influenza all number positive reported vs. GAM
Shifted Week
13 26 39 52
Influenza all − Raw
Influenza all − Negative Binomial GAM

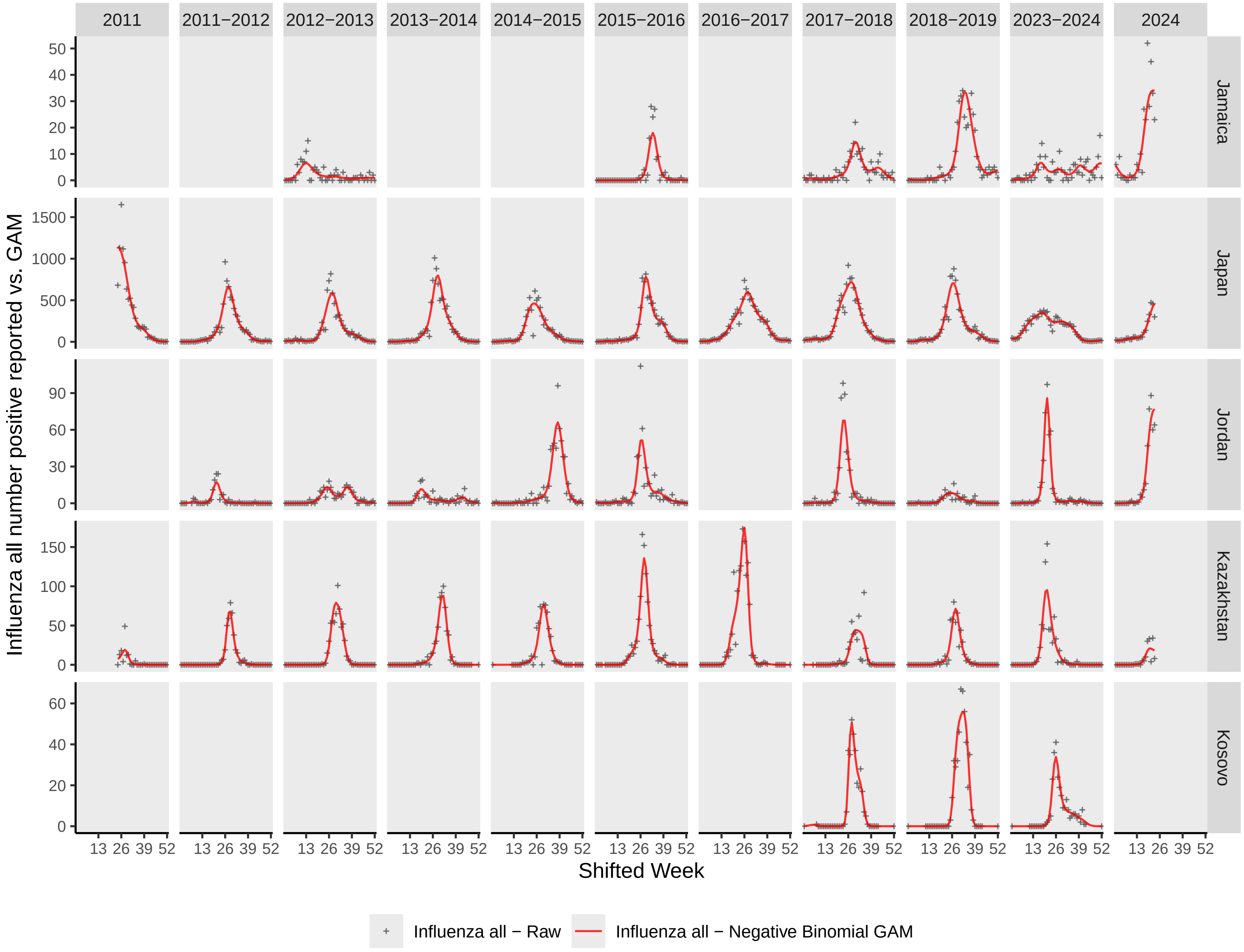

2011
2011−2012
2012−2013
2013−2014
2014−2015
2015−2016
2016−2017
2017−2018
2018−2019
2023−2024
2024
Jamaica
Japan
Jordan
Kazakhstan
Kosovo
Influenza all number positive reported vs. GAM
Shifted Week
Influenza all − Raw
Influenza all − Negative Binomial GAM

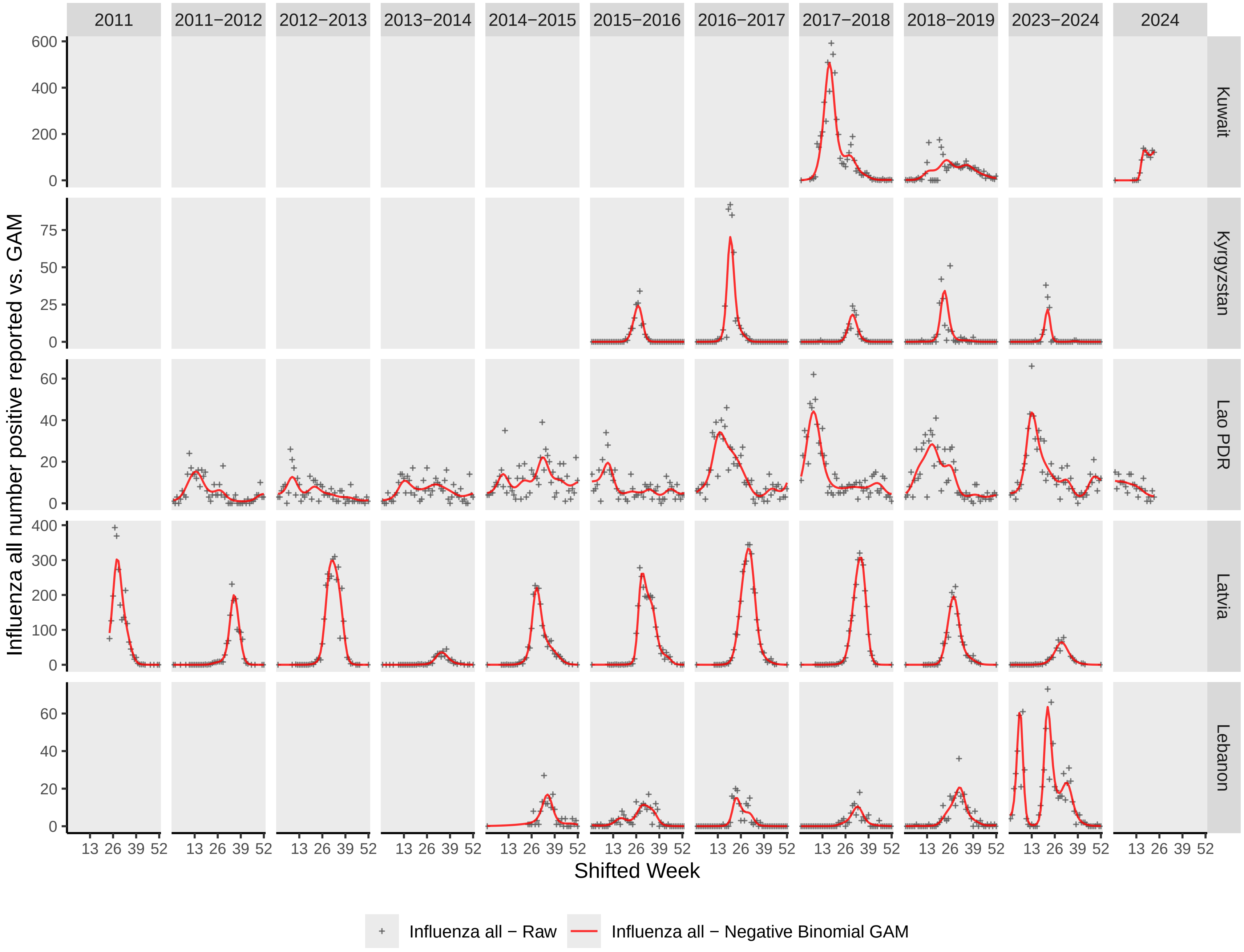

2011
2011−2012
2012−2013
2013−2014
2014−2015
2015−2016
2016−2017
2017−2018
2018−2019
2023−2024
2024
Kuwait
Kyrgyzstan
Lao PDR
Latvia
Lebanon
Influenza all number positive reported vs. GAM
Shifted Week
13 26 39 52
Influenza all − Raw
Influenza all − Negative Binomial GAM

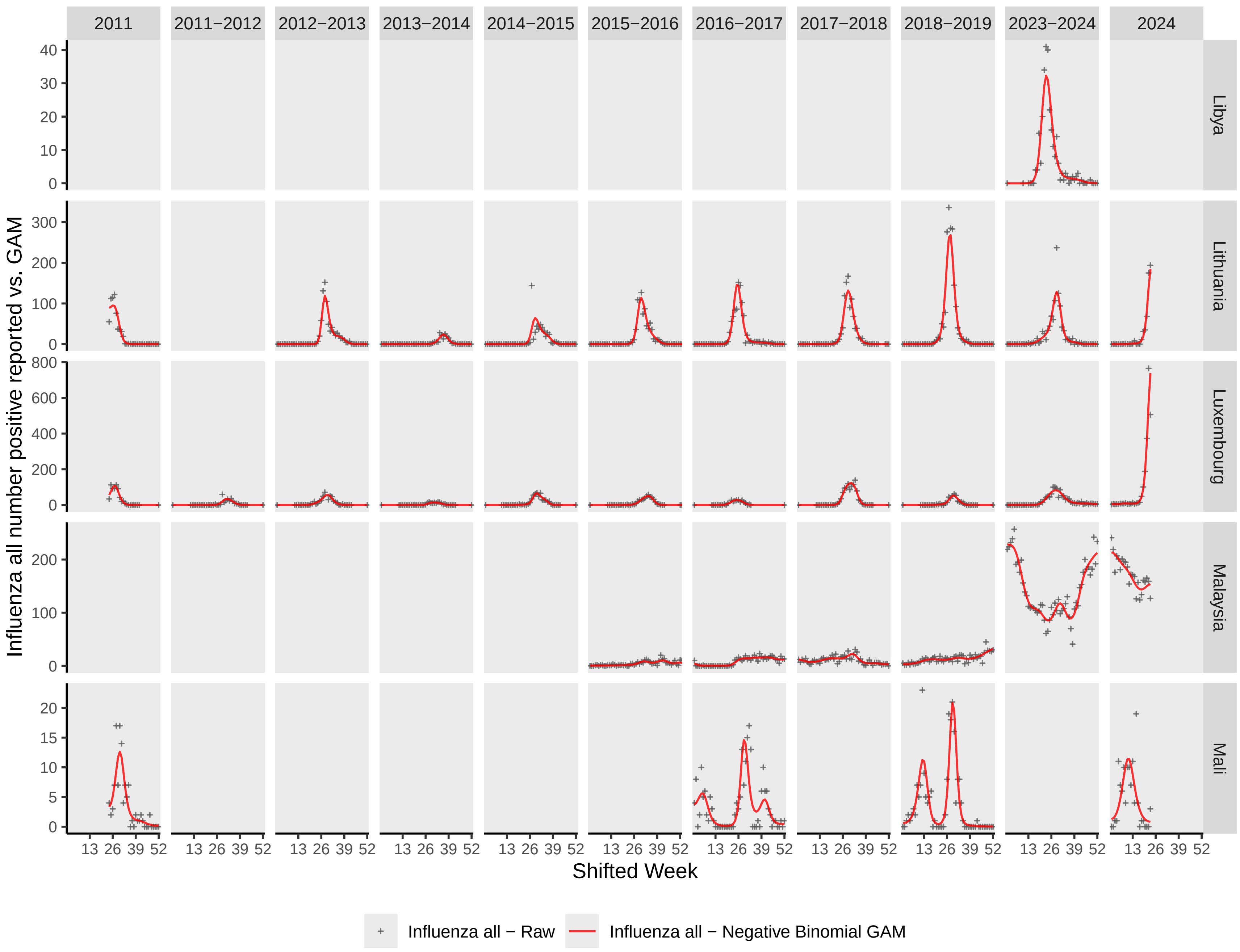

2011
2011−2012
2012−2013
2013−2014
2014−2015
2015−2016
2016−2017
2017−2018
2018−2019
2023−2024
2024
Libya
Lithuania
Luxembourg
Malaysia
Mali
Influenza all number positive reported vs. GAM
Shifted Week
13 26 39 52
Influenza all − Raw
Influenza all − Negative Binomial GAM

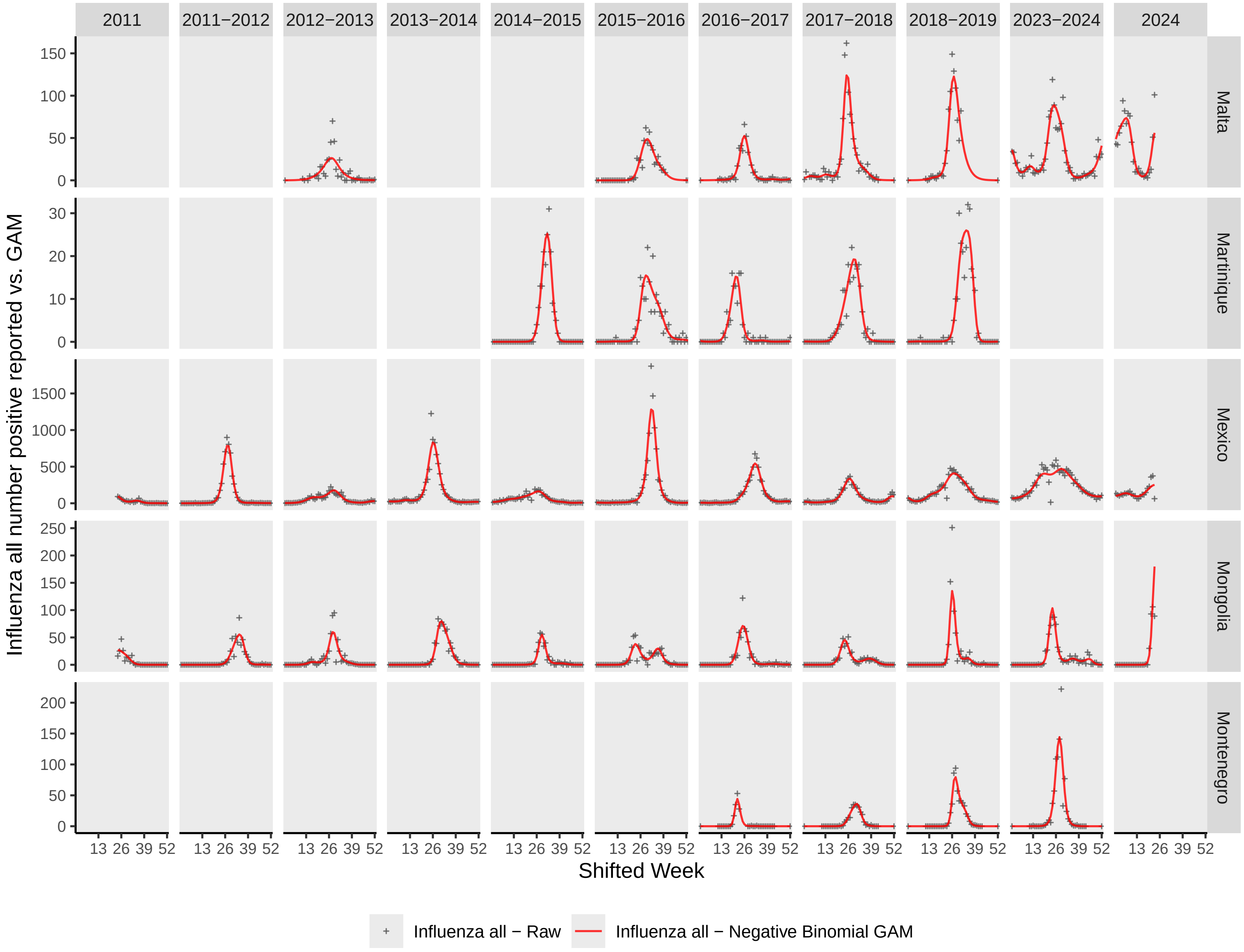

2011
2011−2012
2012−2013
2013−2014
2014−2015
2015−2016
2016−2017
2017−2018
2018−2019
2023−2024
2024
Malta
Martinique
Mexico
Mongolia
Montenegro
Influenza all number positive reported vs. GAM
Shifted Week
13 26 39 52
Influenza all − Raw
Influenza all − Negative Binomial GAM

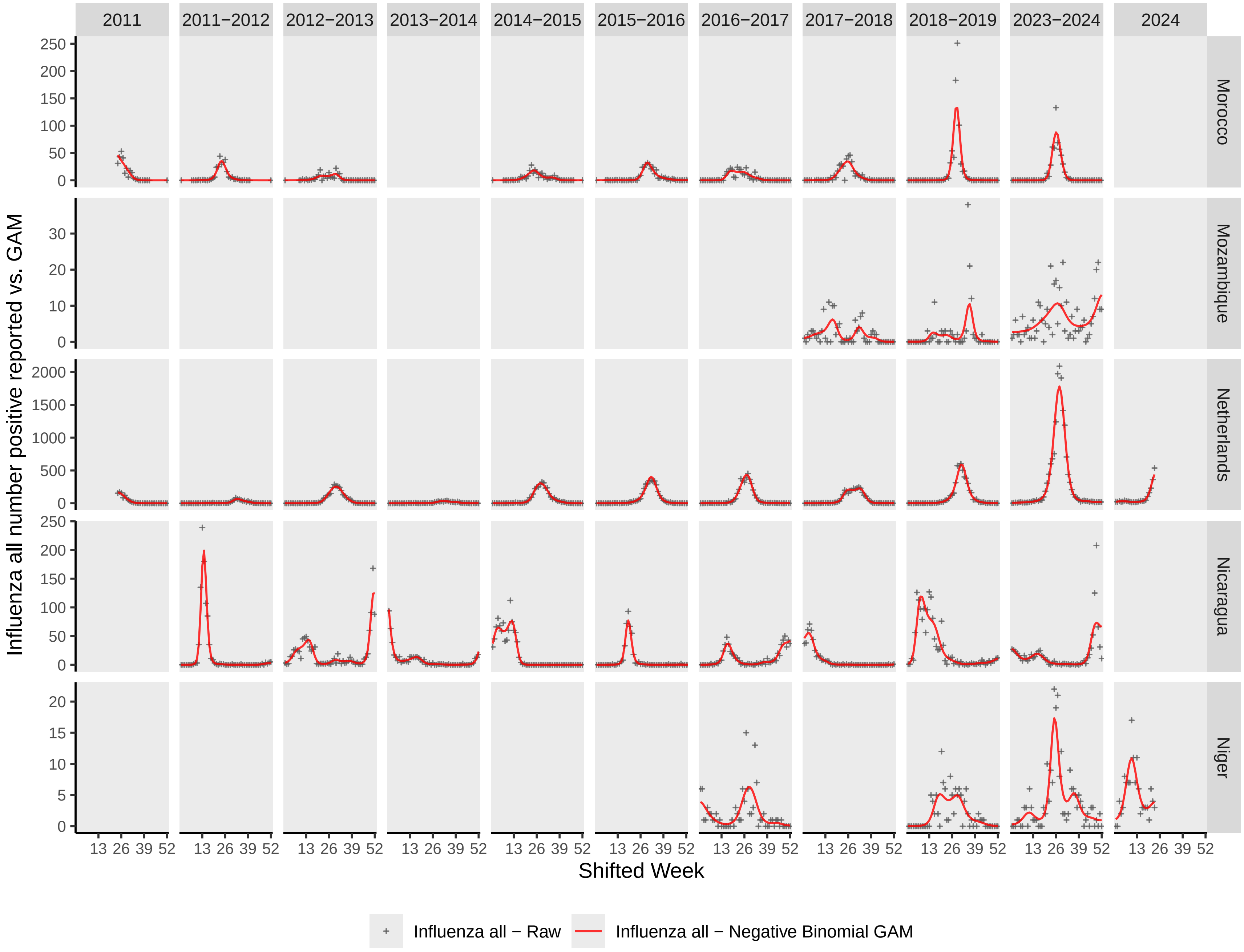
2011
2011−2012
2012−2013
2013−2014
2014−2015
2015−2016
2016−2017
2017−2018
2018−2019
2023−2024
2024
Morocco
Mozambique
Netherlands
Nicaragua
Niger
Influenza all number positive reported vs. GAM
Shifted Week
13 26 39 52
Influenza all − Raw
Influenza all − Negative Binomial GAM

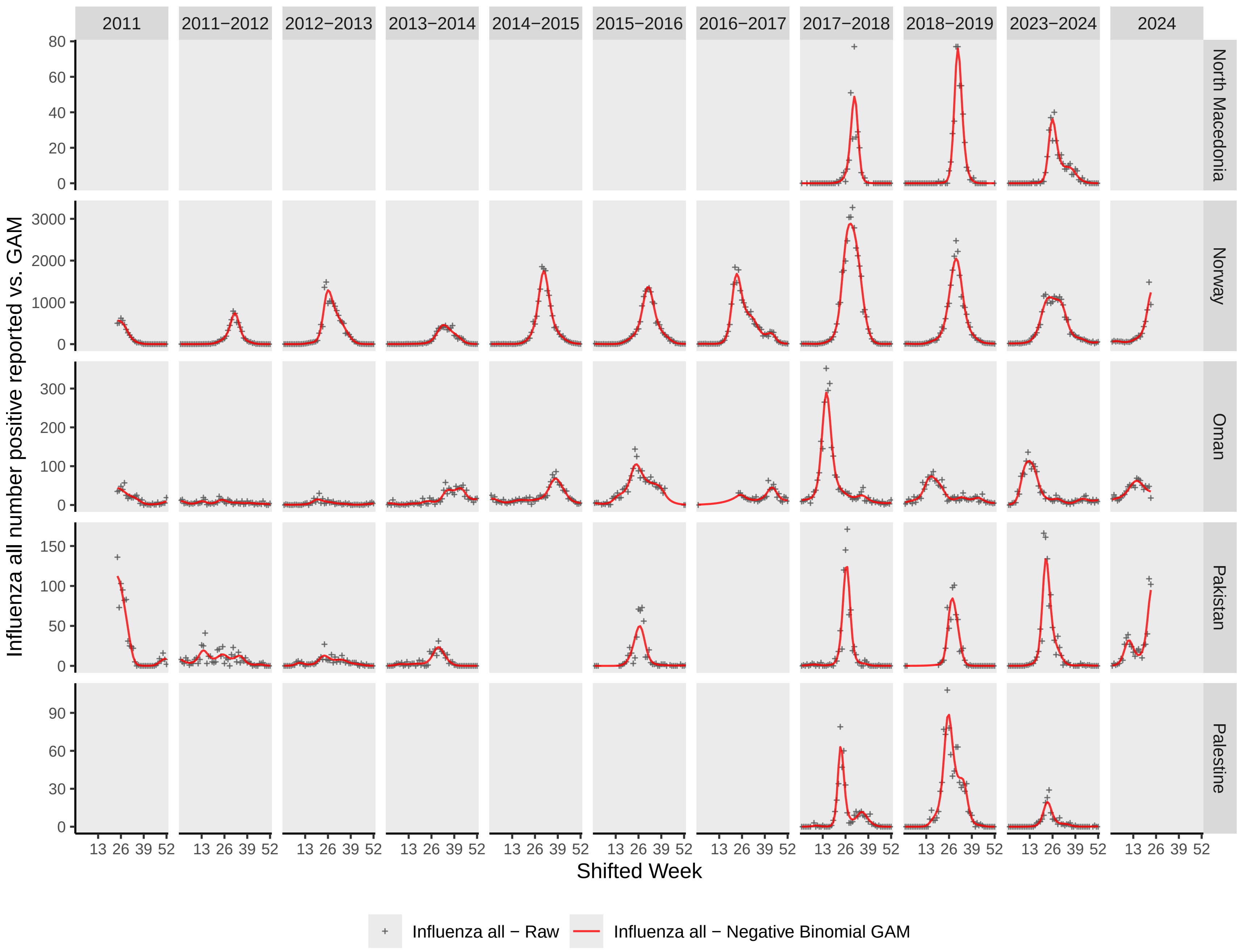

2011
2011−2012
2012−2013
2013−2014
2014−2015
2015−2016
2016−2017
2017−2018
2018−2019
2023−2024
2024
North Macedonia
Norway
Oman
Pakistan
Palestine
Influenza all number positive reported vs. GAM
Shifted Week
13 26 39 52
Influenza all − Raw
Influenza all − Negative Binomial GAM

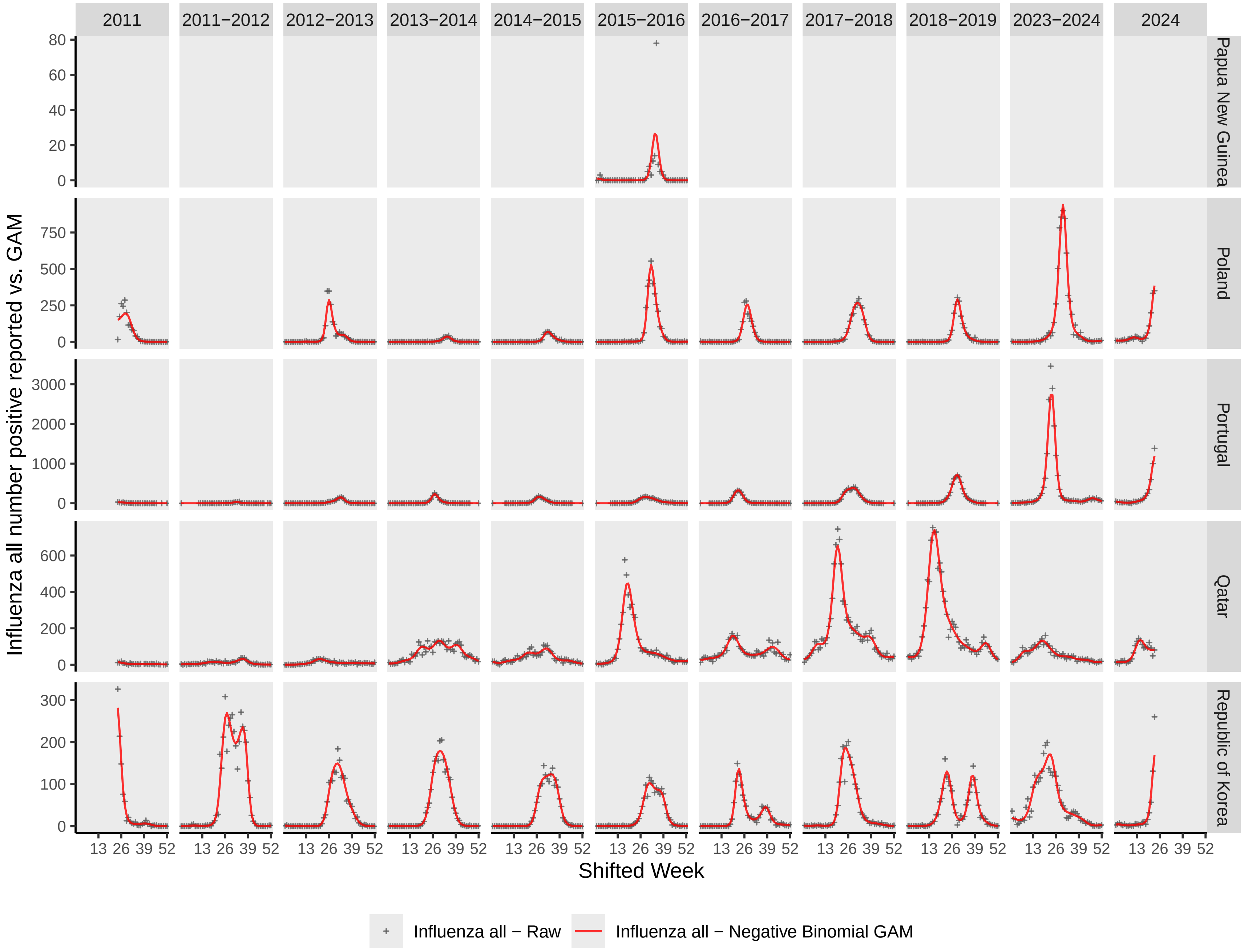
2011
2011–2012
2012–2013
2013–2014
2014–2015
2015–2016
2016–2017
2017–2018
2018–2019
2023–2024
2024
Papua New Guinea
Poland
Portugal
Qatar
Republic of Korea
Influenza all number positive reported vs. GAM
Shifted Week
13 26 39 52
Influenza all − Raw
Influenza all − Negative Binomial GAM

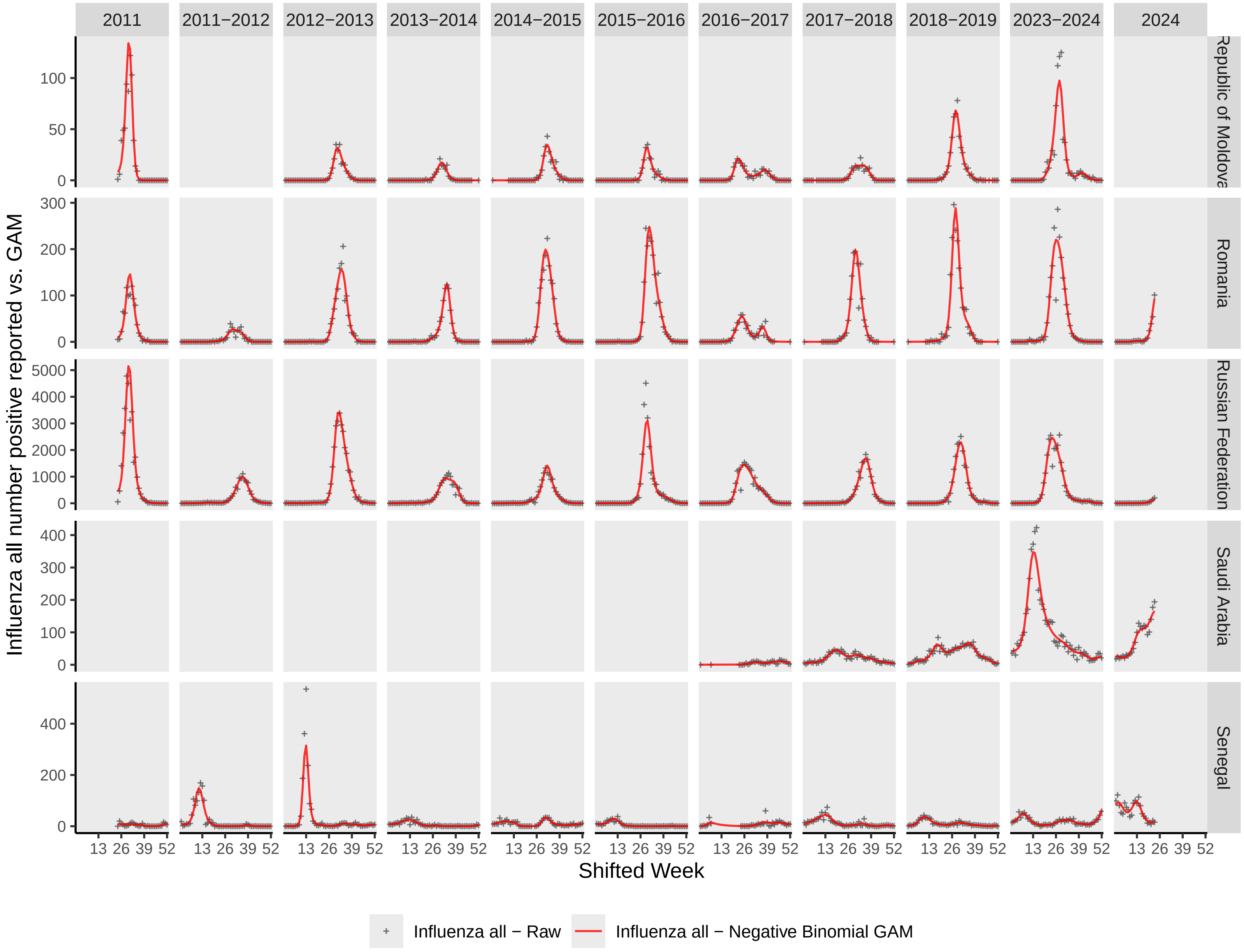

2011
2011−2012
2012−2013
2013−2014
2014−2015
2015−2016
2016−2017
2017−2018
2018−2019
2023−2024
2024
Republic of Moldova
Romania
Russian Federation
Saudi Arabia
Senegal
Influenza all number positive reported vs. GAM
Shifted Week
13 26 39 52
Influenza all − Raw
Influenza all − Negative Binomial GAM

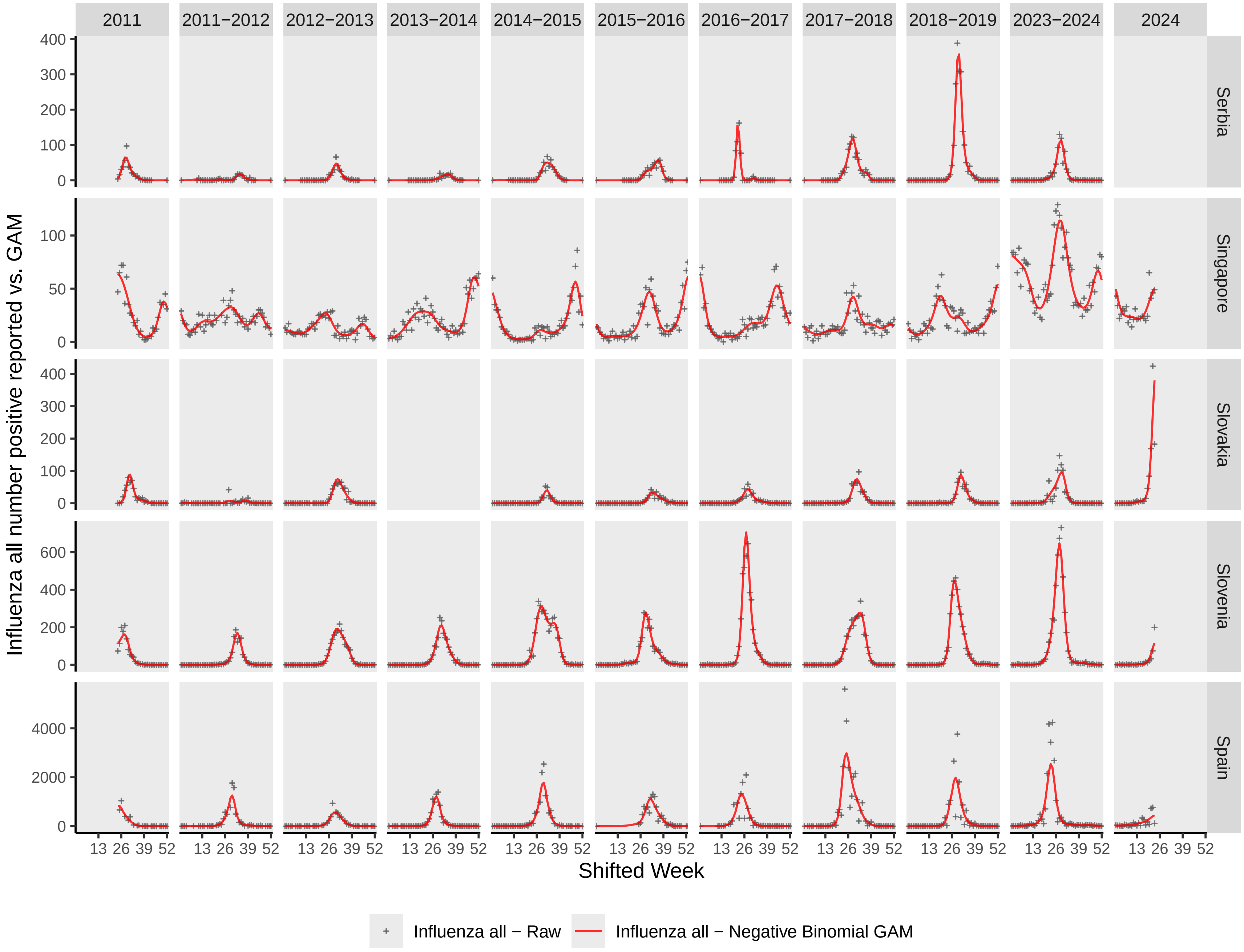
2011
2011−2012
2012−2013
2013−2014
2014−2015
2015−2016
2016−2017
2017−2018
2018−2019
2023−2024
2024
Serbia
Singapore
Slovakia
Slovenia
Spain
Influenza all number positive reported vs. GAM
Shifted Week
13 26 39 52
Influenza all − Raw
Influenza all − Negative Binomial GAM

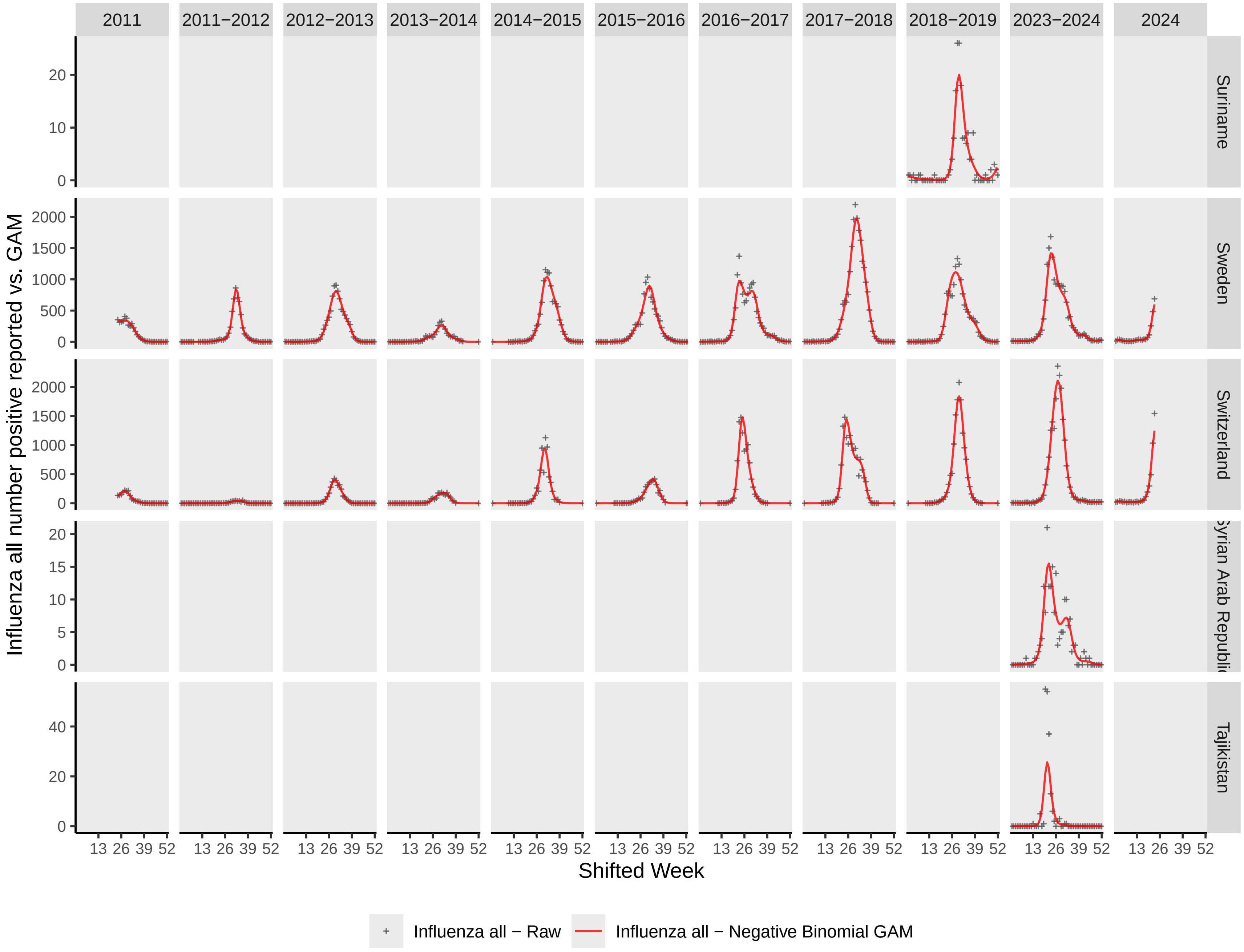
2011
2011−2012
2012−2013
2013−2014
2014−2015
2015−2016
2016−2017
2017−2018
2018−2019
2023−2024
2024
Suriname
Sweden
Switzerland
Syrian Arab Republic
Tajikistan
Influenza all number positive reported vs. GAM
Shifted Week
13 26 39 52
Influenza all − Raw
Influenza all − Negative Binomial GAM

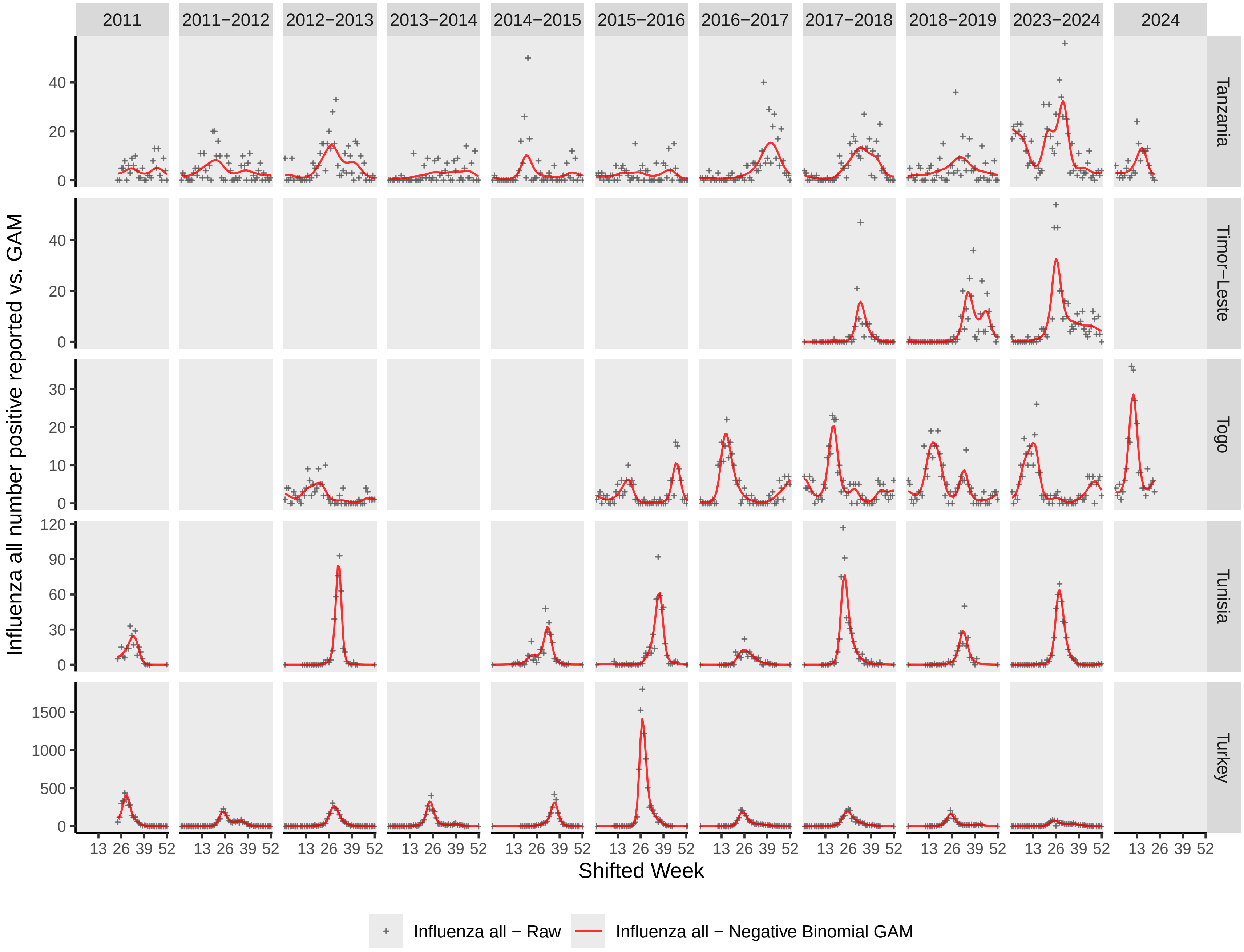
2011
2011−2012
2012−2013
2013−2014
2014−2015
2015−2016
2016−2017
2017−2018
2018−2019
2023−2024
2024
Tanzania
Timor−Leste
Togo
Tunisia
Turkey
Influenza all number positive reported vs. GAM
Shifted Week
13 26 39 52
0
20
40
10
30
60
90
120
500
1000
1500
Influenza all − Raw
Influenza all − Negative Binomial GAM

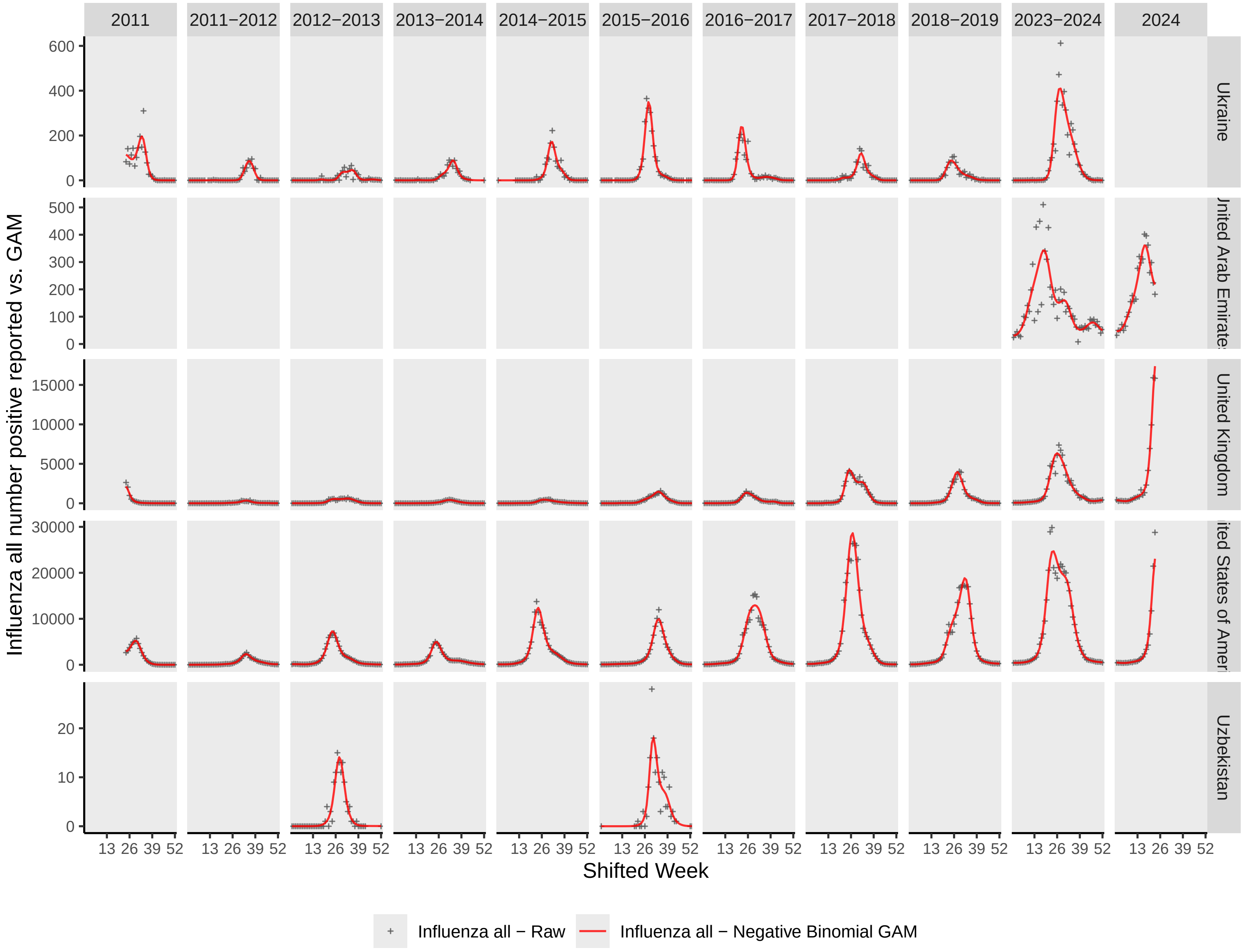

2011
2011−2012
2012−2013
2013−2014
2014−2015
2015−2016
2016−2017
2017−2018
2018−2019
2023−2024
2024
Ukraine
United Arab Emirates
United Kingdom
United States of America
Uzbekistan
Influenza all number positive reported vs. GAM
Shifted Week
13 26 39 52
Influenza all − Raw
Influenza all − Negative Binomial GAM

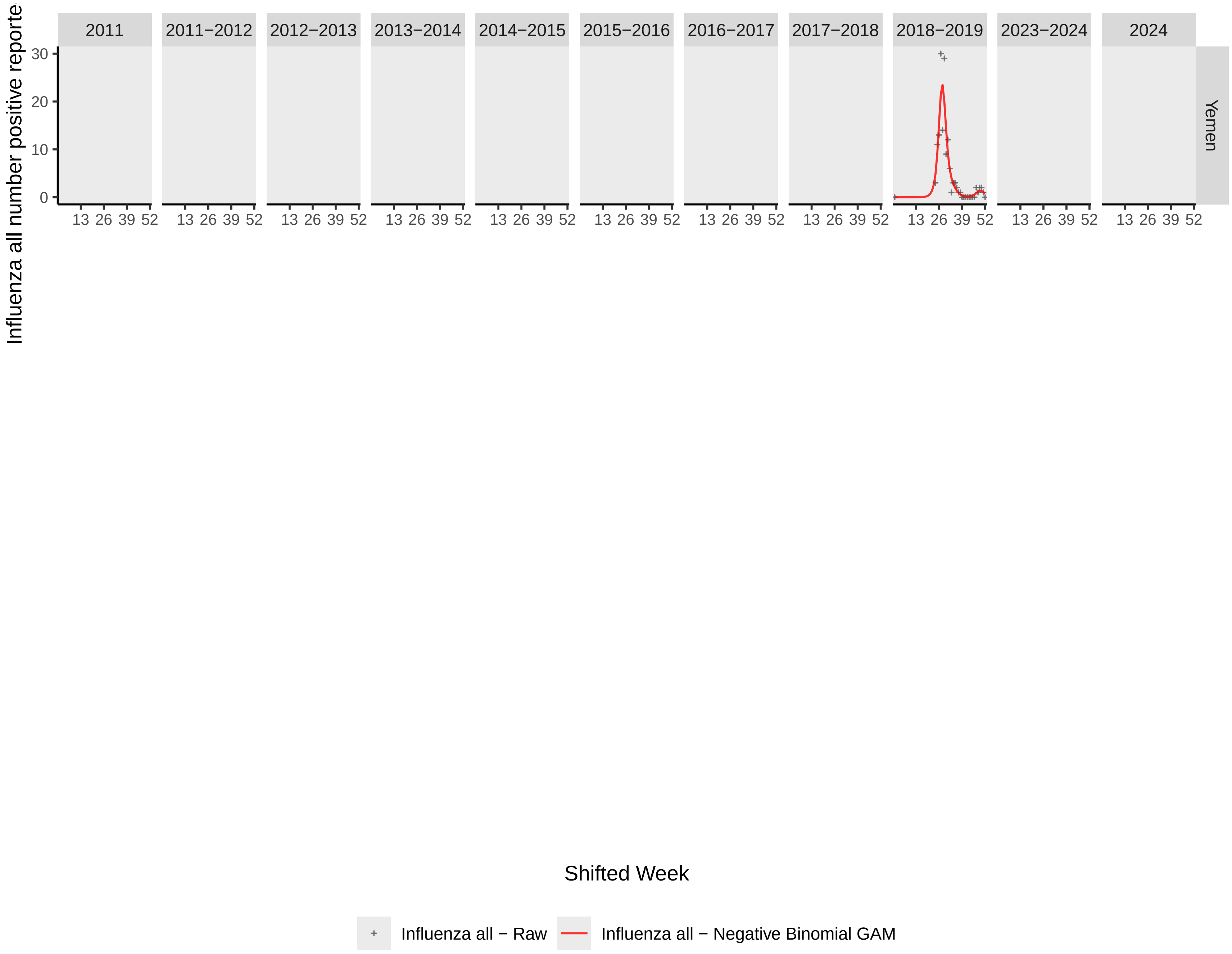

Influenza all number positive reporte
2011
2011−2012
2012−2013
2013−2014
2014−2015
2015−2016
2016−2017
2017−2018
2018−2019
2023−2024
2024
Yemen
30
20
10
0
13 26 39 52
Shifted Week
Influenza all − Raw
Influenza all − Negative Binomial GAM

**Supplementary Figure 2**: Determination of optimal clustering algorithm (k-means [kmeans], hierarchical k-means [hkmeans], hierarchical [hclust_ward], or partitioning around medoids [pam]) and cluster size (k) for calendar year (**A–D**) and shifted year (**E–H**) partitions. **A and E:** Composite scores (range: 0–1) calculated from Calinski-Harabasz indices, Dunn indices, Silhouette scores, and proportion of ambiguous clustering (PAC) scores for each algorithm and k. Values closer to 1 indicating better algorithm performance. **B and F:** Within-cluster sum of squares (WSS); **C and G:** mean Silhouette width; and **D and H:** Calinski-Harabasz index to determine optimal k. Optimal k is evaluated as the "elbow," maximum mean Silhouette width, and Calinski-Harabasz index peak for each time-window partition, identified with a vertical dotted red line.

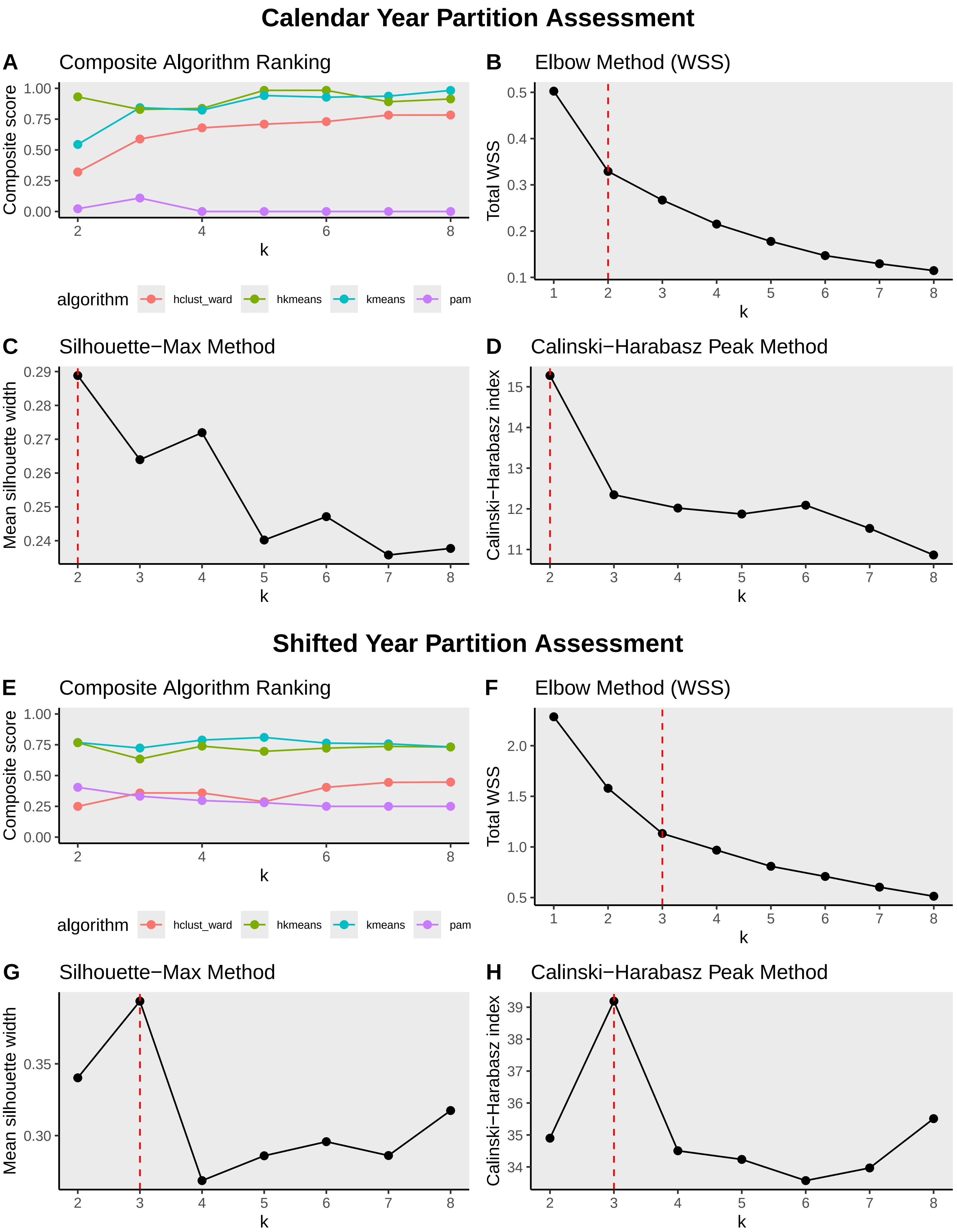
Calendar Year Partition Assessment
A
Composite Algorithm Ranking
Composite score
k
algorithm
hclust_ward
hkmeans
kmeans
pam
B
Elbow Method (WSS)
Total WSS
k
C
Silhouette−Max Method
Mean silhouette width
k
D
Calinski−Harabasz Peak Method
Calinski−Harabasz index
k
Shifted Year Partition Assessment
E
Composite Algorithm Ranking
Composite score
k
algorithm
hclust_ward
hkmeans
kmeans
pam
F
Elbow Method (WSS)
Total WSS
k
G
Silhouette−Max Method
Mean silhouette width
k
H
Calinski−Harabasz Peak Method
Calinski−Harabasz index
k

**Supplementary Figure 3**: Map of country clusters, or influenza epidemic zones, resulting from primary clustering analysis. Epidemic zone A–C corresponded to the three clusters identified from analyzing countries with a shifted-year time window, while epidemic zones D and E corresponded to the two clusters identified from analyzing countries with a calendar-year time window.

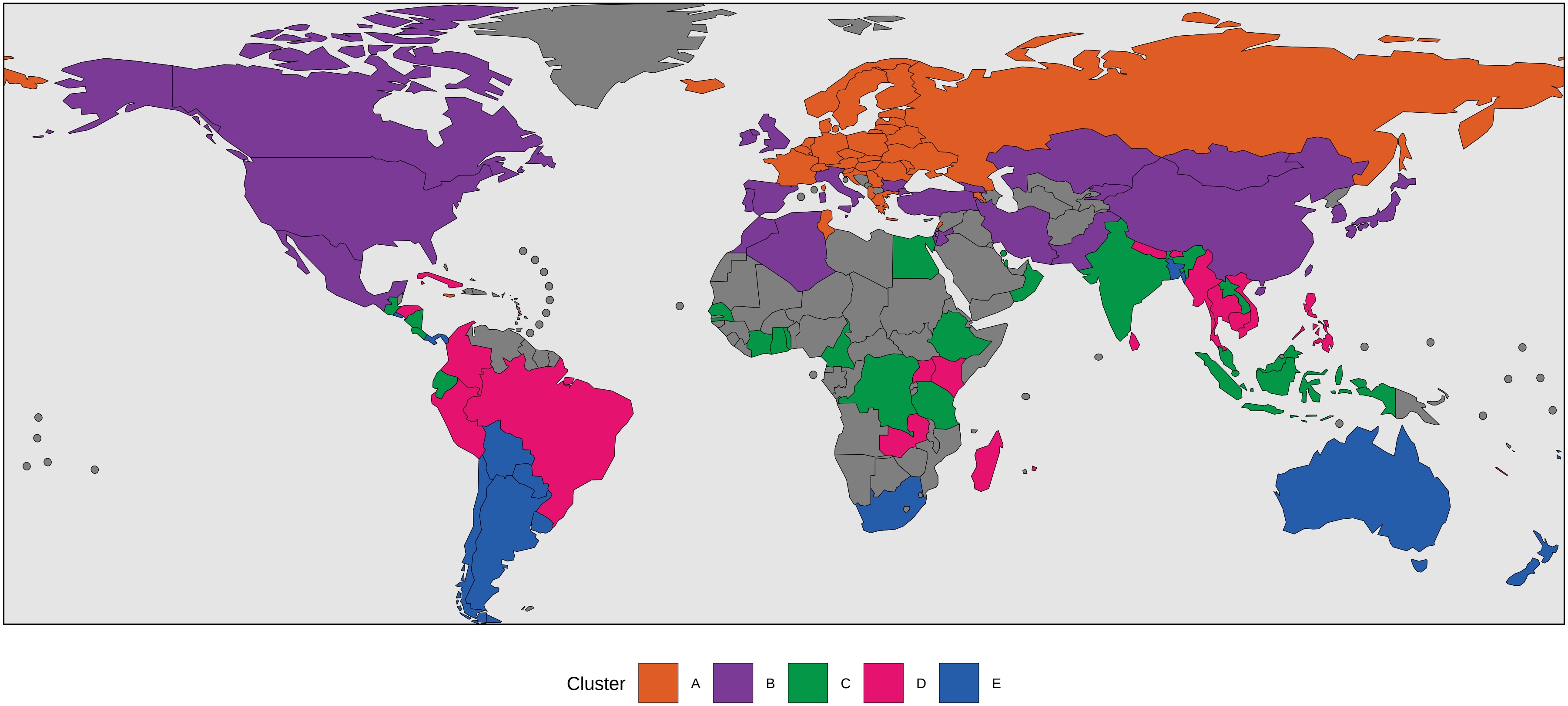
Cluster
A
B
C
D
E

**Supplementary Figure 4**: Summary seasonal curves of relative influenza burden for each country, calculated from generalized additive models (GAMs). GAMs of individual time-window years are depicted in the gray open circles, while the summary seasonal GAM is in the solid line, color-coded to match the country's assigned epidemic zone.

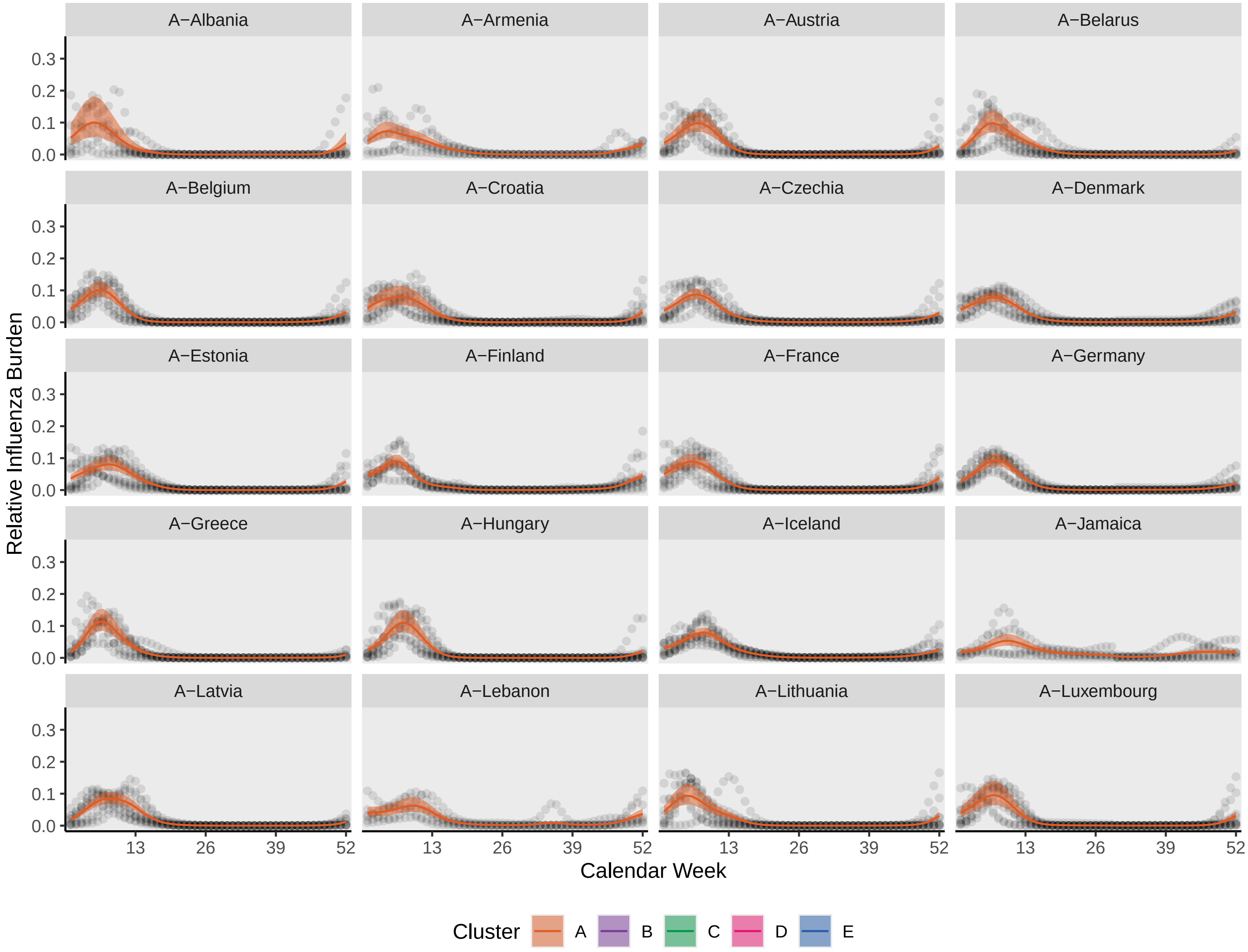

A−Albania
A−Armenia
A−Austria
A−Belarus
A−Belgium
A−Croatia
A−Czechia
A−Denmark
A−Estonia
A−Finland
A−France
A−Germany
A−Greece
A−Hungary
A−Iceland
A−Jamaica
A−Latvia
A−Lebanon
A−Lithuania
A−Luxembourg
Relative Influenza Burden
0.0
0.1
0.2
0.3
13
26
39
52
Calendar Week
Cluster
A
B
C
D
E

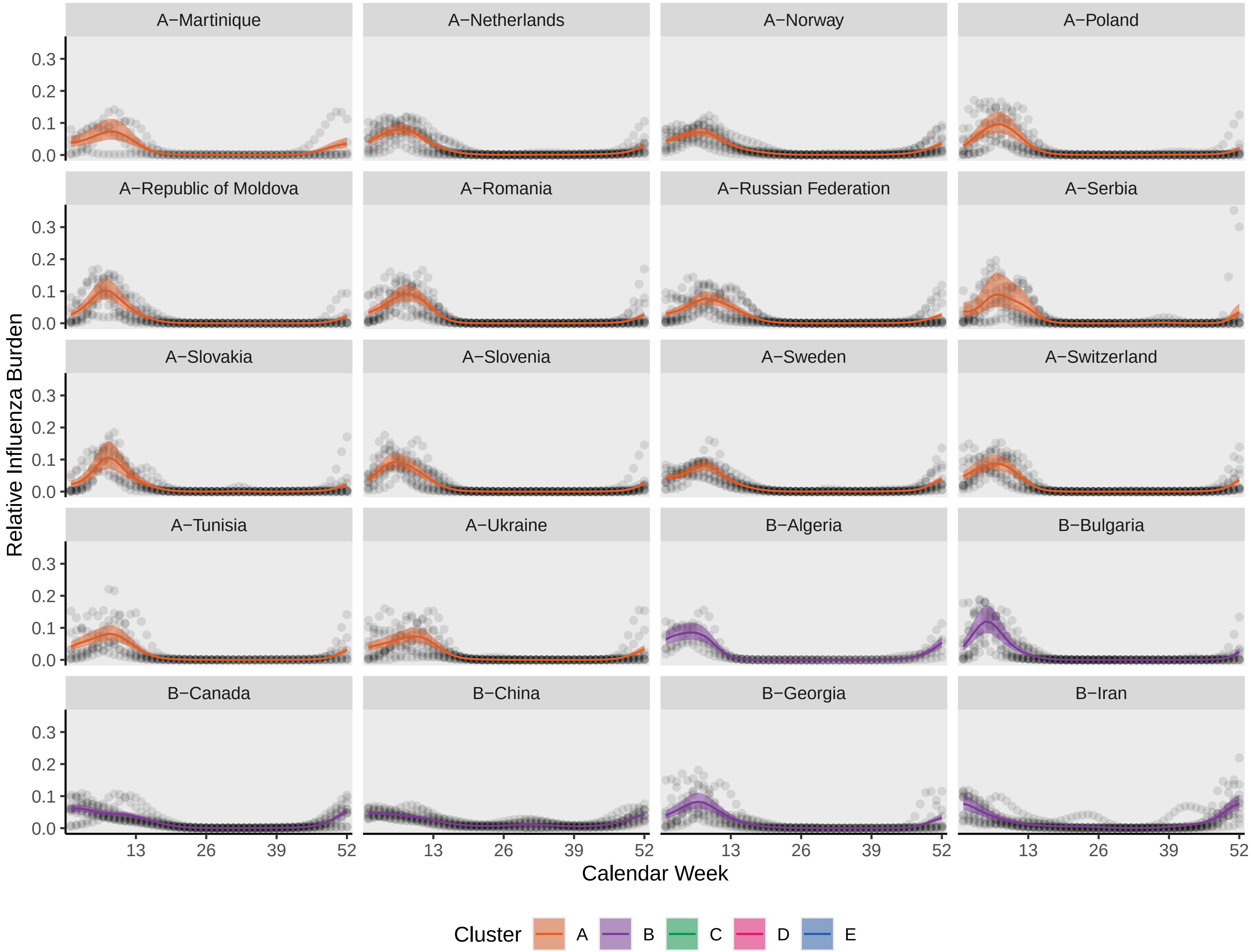
A−Martinique
A−Netherlands
A−Norway
A−Poland
A−Republic of Moldova
A−Romania
A−Russian Federation
A−Serbia
A−Slovakia
A−Slovenia
A−Sweden
A−Switzerland
A−Tunisia
A−Ukraine
B−Algeria
B−Bulgaria
B−Canada
B−China
B−Georgia
B−Iran
Relative Influenza Burden
0.0
0.1
0.2
0.3
13
26
39
52
Calendar Week
Cluster
A
B
C
D
E

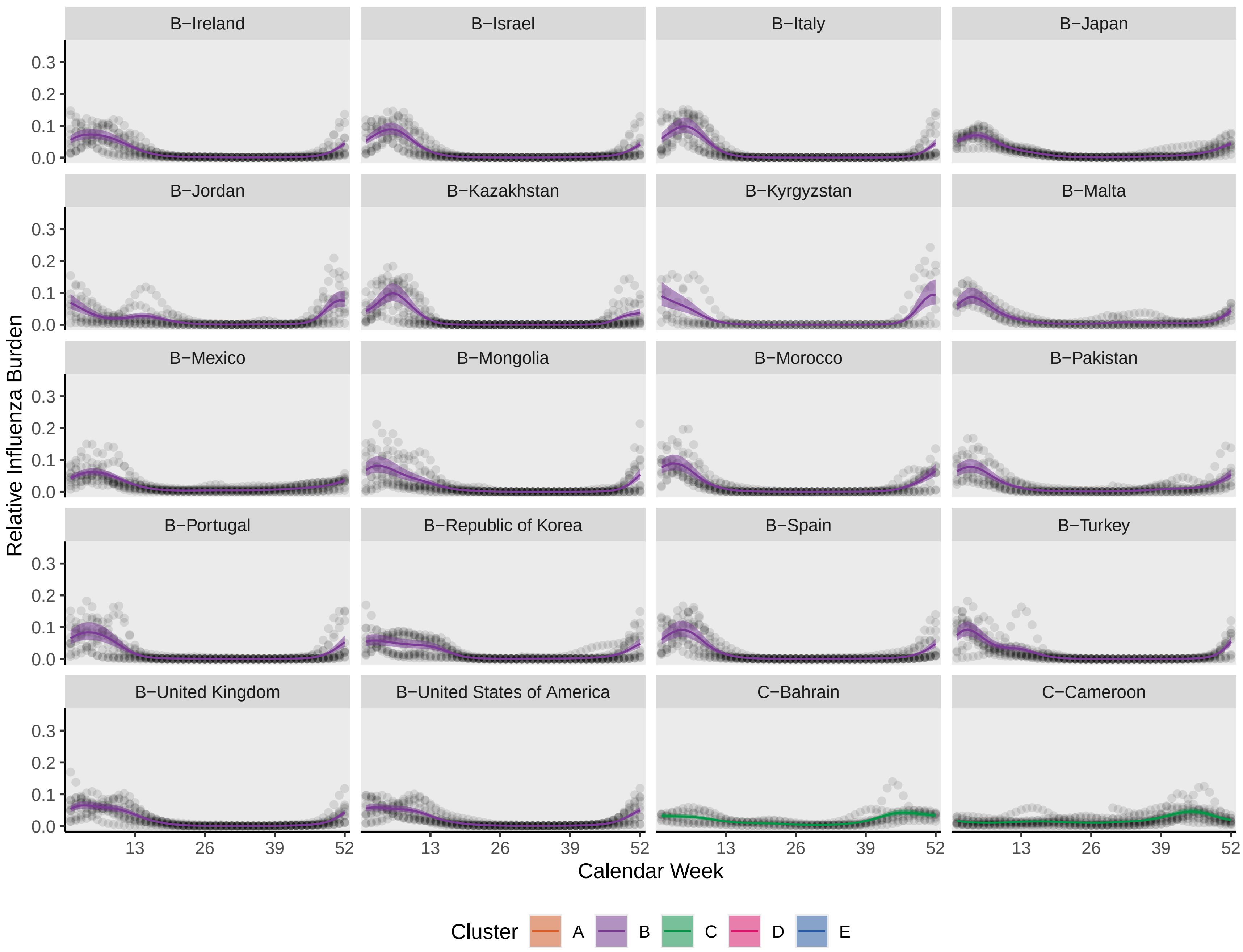
B−Ireland
B−Israel
B−Italy
B−Japan
B−Jordan
B−Kazakhstan
B−Kyrgyzstan
B−Malta
B−Mexico
B−Mongolia
B−Morocco
B−Pakistan
B−Portugal
B−Republic of Korea
B−Spain
B−Turkey
B−United Kingdom
B−United States of America
C−Bahrain
C−Cameroon
0.0
0.1
0.2
0.3
13
26
39
52
Relative Influenza Burden
Calendar Week
Cluster
A
B
C
D
E

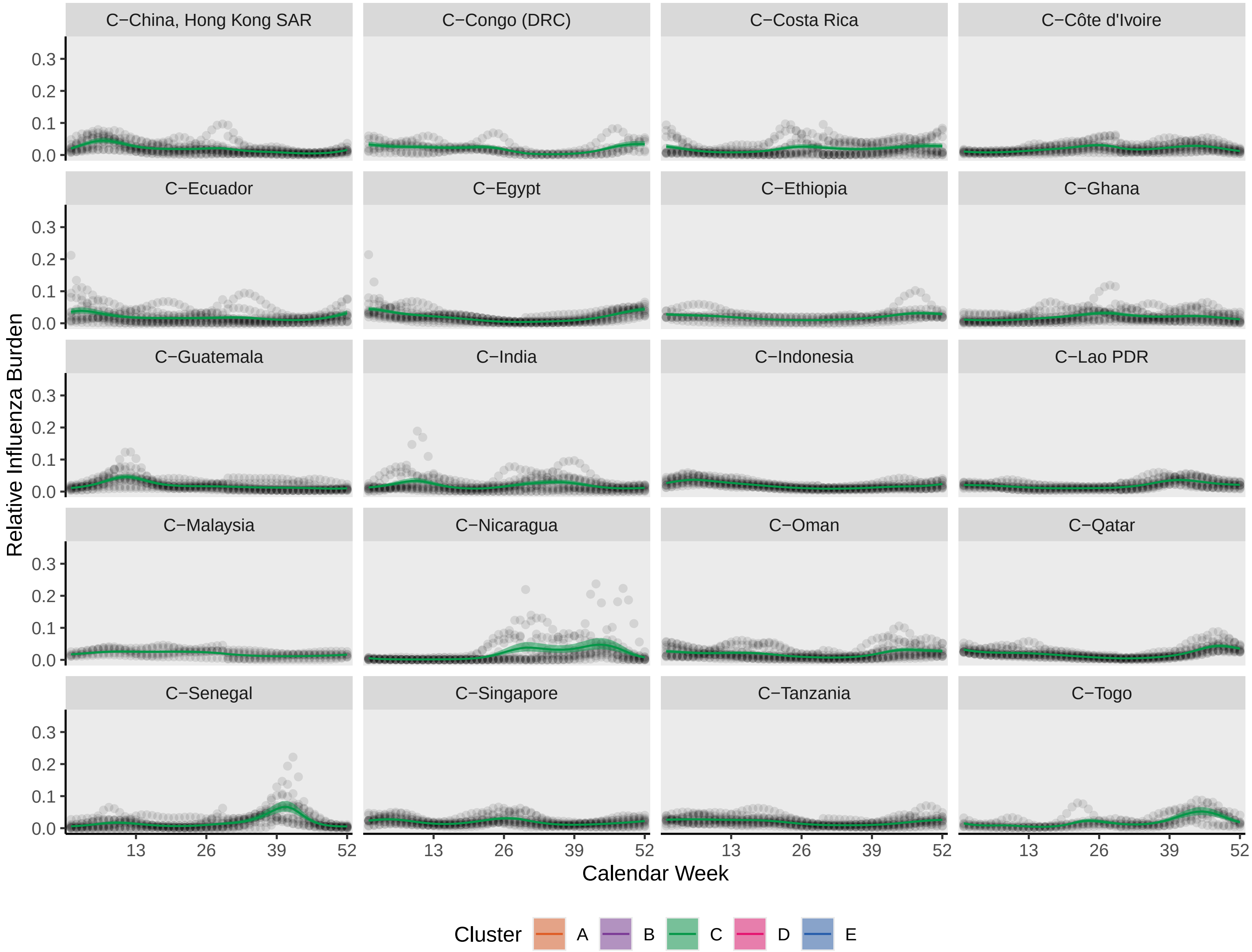
C−China, Hong Kong SAR
C−Congo (DRC)
C−Costa Rica
C−Côte d'Ivoire
C−Ecuador
C−Egypt
C−Ethiopia
C−Ghana
C−Guatemala
C−India
C−Indonesia
C−Lao PDR
C−Malaysia
C−Nicaragua
C−Oman
C−Qatar
C−Senegal
C−Singapore
C−Tanzania
C−Togo
Relative Influenza Burden
0.0
0.1
0.2
0.3
Calendar Week
13
26
39
52
Cluster
A
B
C
D
E

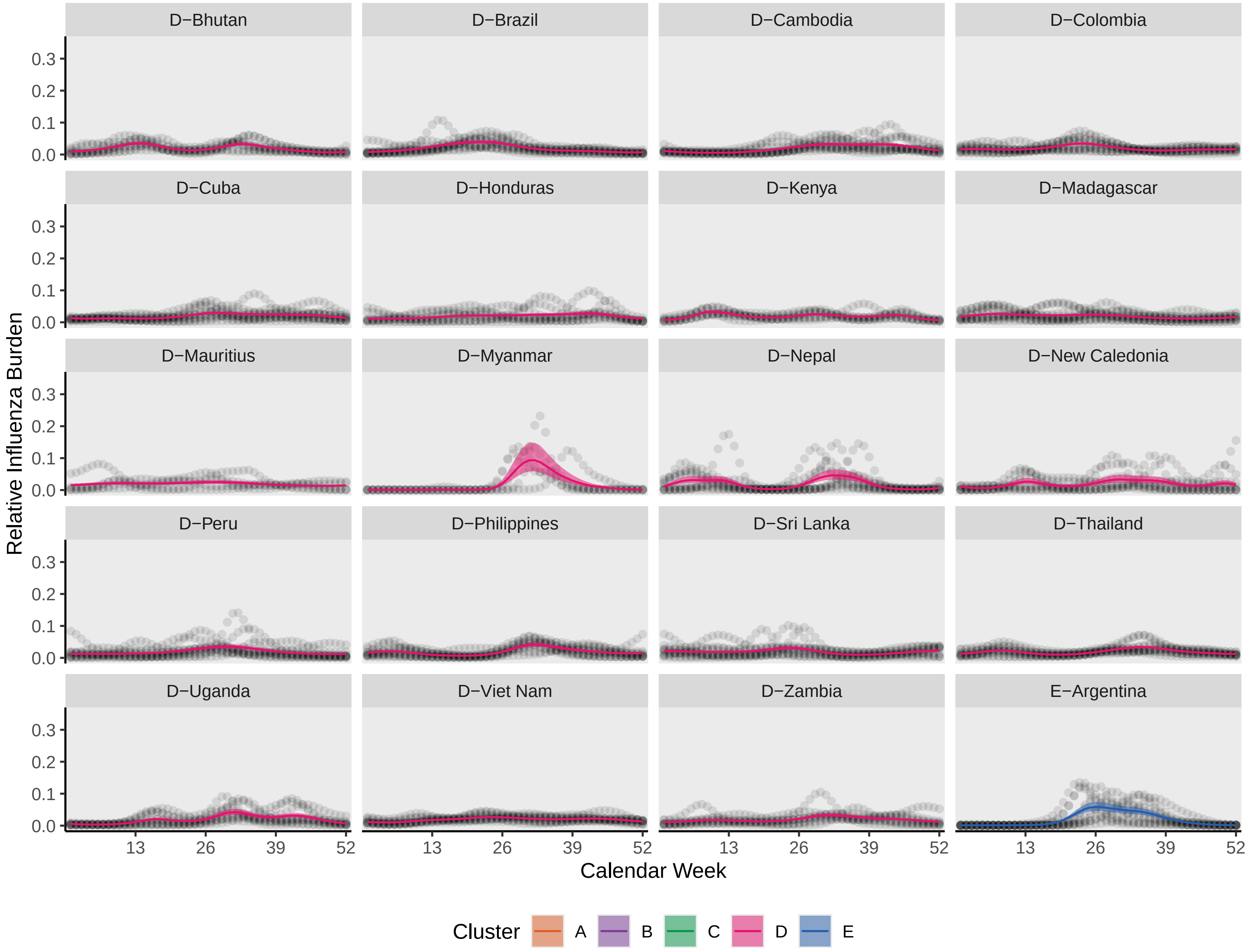
D−Bhutan
D−Brazil
D−Cambodia
D−Colombia
D−Cuba
D−Honduras
D−Kenya
D−Madagascar
D−Mauritius
D−Myanmar
D−Nepal
D−New Caledonia
D−Peru
D−Philippines
D−Sri Lanka
D−Thailand
D−Uganda
D−Viet Nam
D−Zambia
E−Argentina
Relative Influenza Burden
0.0
0.1
0.2
0.3
Calendar Week
13
26
39
52
Cluster
A
B
C
D
E

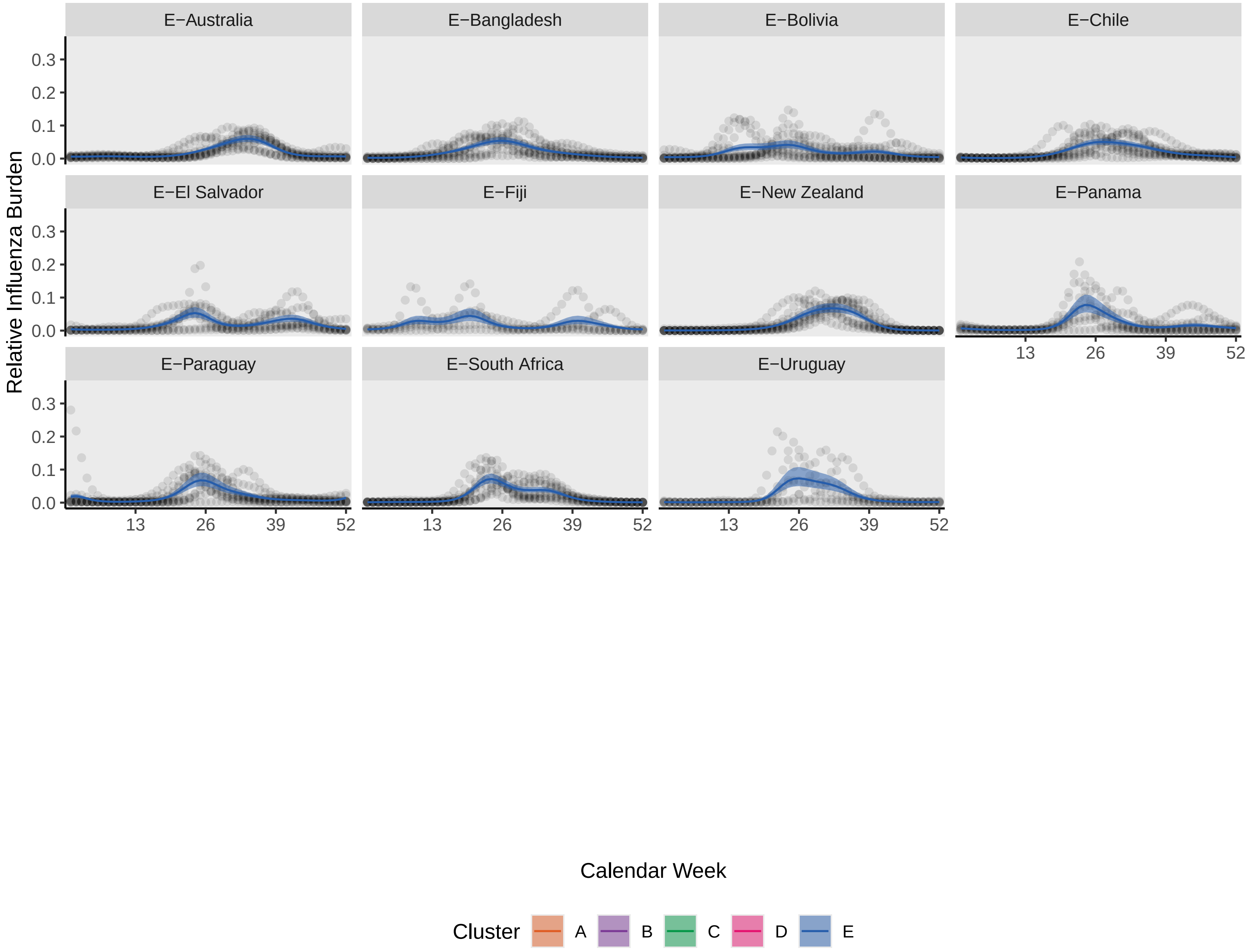
E−Australia
E−Bangladesh
E−Bolivia
E−Chile
E−El Salvador
E−Fiji
E−New Zealand
E−Panama
E−Paraguay
E−South Africa
E−Uruguay
Relative Influenza Burden
0.0
0.1
0.2
0.3
13
26
39
52
Calendar Week
Cluster
A
B
C
D
E

**Supplementary Figure 5**: Distribution of countries in the previously defined 18 influenza transmission zones across the five epidemic zones identified in this analysis.

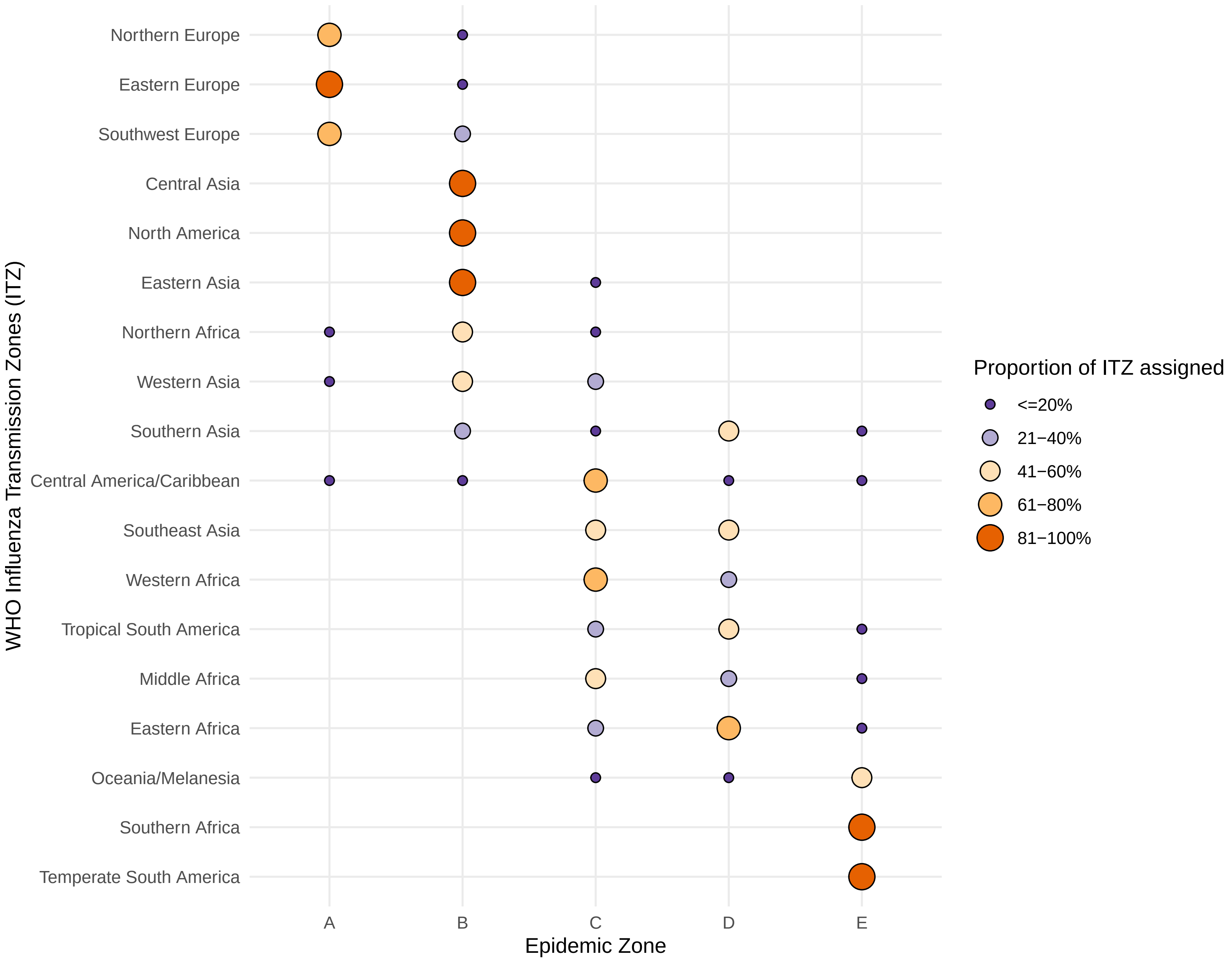

Northern Europe
Eastern Europe
Southwest Europe
Central Asia
North America
Eastern Asia
Northern Africa
Western Asia
Southern Asia
Central America/Caribbean
Southeast Asia
Western Africa
Tropical South America
Middle Africa
Eastern Africa
Oceania/Melanesia
Southern Africa
Temperate South America
WHO Influenza Transmission Zones (ITZ)
A
B
C
D
E
Epidemic Zone
Proportion of ITZ assigned
<=20%
21−40%
41−60%
61−80%
81−100%

**Supplementary Table 1**: Timing of influenza virus detection based on weekly surveillance data submitted to the World Health Organization Global Influenza Surveillance and Response System, 2011–2019 and 2023–2024.

| Country | Calendar quarter (Q) in which most influenza virus detections occurred | Time window used for analysis | Number of time windows with ≥100 detections | Number of time windows with data for ≥40% of weeks | Number of complete time windows available for analysis | Data available during anchor year* | Inclusion† into clustering analysis |
|---|---|---|---|---|---|---|---|
| **Afghanistan** | Q1 | Shifted year | 5 | 9 | 4 | Not applicable: too few time windows for analysis | Exclude: too few time windows |
| **Albania** | Q1 | Shifted year | 8 | 11 | 7 | Yes | Include |
| **Algeria** | Q1 | Shifted year | 6 | 9 | 5 | Yes | Include |
| **Angola** | Q2 | Calendar year | 0 | 2 | 0 | Not applicable: too few time windows for analysis | Exclude: too few time windows |
| **Antigua and Barbuda** | Q2 | Calendar year | 0 | 0 | 0 | Not applicable: too few time windows for analysis | Exclude: too few time windows |
| **Argentina** | Q3 | Calendar year | 10 | 10 | 10 | Yes | Include |
| **Armenia** | Q1 | Shifted year | 6 | 11 | 6 | Yes | Include |
| **Aruba** | Q1 | Shifted year | 1 | 3 | 1 | Not applicable: too few time windows for analysis | Exclude: too few time windows |
| **Australia** | Q3 | Calendar year | 10 | 10 | 10 | Yes | Include |
| **Austria** | Q1 | Shifted year | 11 | 11 | 9 | Yes | Include |
| **Azerbaijan** | Q1 | Shifted year | 3 | 11 | 3 | Not applicable: too few time windows for analysis | Exclude: too few time windows |
| **Bahamas** | Q1 | Shifted year | 0 | 0 | 0 | Not applicable: too few time windows for analysis | Exclude: too few time windows |
| **Bahrain** | Q4 | Shifted year | 7 | 10 | 6 | Yes | Include |

| Country | Calendar quarter (Q) in which most influenza virus detections occurred | Time window used for analysis | Number of time windows with ≥100 detections | Number of time windows with data for ≥40% of weeks | Number of complete time windows available for analysis | Data available during anchor year* | Inclusion† into clustering analysis |
|---|---|---|---|---|---|---|---|
| **Bangladesh** | Q3 | Calendar year | 10 | 10 | 10 | Yes | Include |
| **Barbados** | Q1 | Shifted year | 2 | 7 | 2 | Not applicable: too few time windows for analysis | Exclude: too few time windows |
| **Belarus** | Q1 | Shifted year | 9 | 11 | 8 | Yes | Include |
| **Belgium** | Q1 | Shifted year | 11 | 11 | 9 | Yes | Include |
| **Belize** | Q4 | Shifted year | 0 | 5 | 0 | Not applicable: too few time windows for analysis | Exclude: too few time windows |
| **Bermuda** | Q1 | Shifted year | 0 | 0 | 0 | Not applicable: too few time windows for analysis | Exclude: too few time windows |
| **Bhutan** | Q3 | Calendar year | 8 | 10 | 8 | Yes | Include |
| **Bolivia** | Q2 | Calendar year | 10 | 10 | 10 | Yes | Include |
| **Bosnia and Herzegovina** | Q1 | Shifted year | 2 | 7 | 1 | Not applicable: too few time windows for analysis | Exclude: too few time windows |
| **Brazil** | Q2 | Calendar year | 10 | 10 | 10 | Yes | Include |
| **Brunei Darussalam** | Q3 | Calendar year | 1 | 1 | 1 | Not applicable: too few time windows for analysis | Exclude: too few time windows |
| **Bulgaria** | Q1 | Shifted year | 10 | 11 | 9 | Yes | Include |
| **Burkina Faso** | Q1 | Shifted year | 5 | 9 | 4 | Not applicable: too few time windows for analysis | Exclude: too few time windows |
| **Cambodia** | Q3 | Calendar year | 10 | 10 | 10 | Yes | Include |

| Country | Calendar quarter (Q) in which most influenza virus detections occurred | Time window used for analysis | Number of time windows with ≥100 detections | Number of time windows with data for ≥40% of weeks | Number of complete time windows available for analysis | Data available during anchor year* | Inclusion† into clustering analysis |
|---|---|---|---|---|---|---|---|
| **Cameroon** | Q4 | Shifted year | 10 | 11 | 9 | Yes | Include |
| **Canada** | Q1 | Shifted year | 11 | 11 | 9 | Yes | Include |
| **Cayman Islands** | Q1 | Shifted year | 1 | 2 | 1 | Not applicable: too few time windows for analysis | Exclude: too few time windows |
| **Central African Republic** | Q3 | Calendar year | 4 | 8 | 4 | Not applicable: too few time windows for analysis | Exclude: too few time windows |
| **Chad** | Q1 | Shifted year | 0 | 4 | 0 | Not applicable: too few time windows for analysis | Exclude: too few time windows |
| **Chile** | Q3 | Calendar year | 10 | 10 | 10 | Yes | Include |
| **China** | Q1 | Shifted year | 11 | 11 | 9 | Yes | Include |
| **China, Hong Kong SAR** | Q1 | Shifted year | 11 | 11 | 9 | Yes | Include |
| **Colombia** | Q2 | Calendar year | 10 | 10 | 10 | Yes | Include |
| **Congo** | Q2 | Calendar year | 0 | 1 | 0 | Not applicable: too few time windows for analysis | Exclude: too few time windows |
| **Congo (DRC)** | Q4 | Shifted year | 6 | 11 | 5 | Yes | Include |
| **Costa Rica** | Q4 | Shifted year | 10 | 11 | 9 | Yes | Include |
| **Croatia** | Q1 | Shifted year | 11 | 11 | 9 | Yes | Include |
| **Cuba** | Q3 | Calendar year | 10 | 10 | 10 | Yes | Include |

| Country | Calendar quarter (Q) in which most influenza virus detections occurred | Time window used for analysis | Number of time windows with ≥100 detections | Number of time windows with data for ≥40% of weeks | Number of complete time windows available for analysis | Data available during anchor year* | Inclusion† into clustering analysis |
|---|---|---|---|---|---|---|---|
| **Cyprus** | Q1 | Shifted year | 4 | 5 | 3 | Not applicable: too few time windows for analysis | Exclude: too few time windows |
| **Czechia** | Q1 | Shifted year | 10 | 11 | 8 | Yes | Include |
| **Côte d'Ivoire** | Q4 | Shifted year | 10 | 11 | 9 | Yes | Include |
| **Denmark** | Q1 | Shifted year | 10 | 11 | 8 | Yes | Include |
| **Dominica** | Q1 | Shifted year | 0 | 3 | 0 | Not applicable: too few time windows for analysis | Exclude: too few time windows |
| **Dominican Republic** | Q2 | Calendar year | 2 | 10 | 2 | Not applicable: too few time windows for analysis | Exclude: too few time windows |
| **Ecuador** | Q1 | Shifted year | 11 | 11 | 9 | Yes | Include |
| **Egypt** | Q1 | Shifted year | 11 | 11 | 9 | Yes | Include |
| **El Salvador** | Q2 | Calendar year | 8 | 10 | 8 | Yes | Include |
| **Estonia** | Q1 | Shifted year | 10 | 11 | 9 | Yes | Include |
| **Ethiopia** | Q4 | Shifted year | 7 | 10 | 6 | Yes | Include |
| **Fiji** | Q2 | Calendar year | 5 | 9 | 5 | Yes | Include |
| **Finland** | Q1 | Shifted year | 10 | 10 | 8 | Yes | Include |
| **France** | Q1 | Shifted year | 11 | 11 | 9 | Yes | Include |

| Country | Calendar quarter (Q) in which most influenza virus detections occurred | Time window used for analysis | Number of time windows with ≥100 detections | Number of time windows with data for ≥40% of weeks | Number of complete time windows available for analysis | Data available during anchor year* | Inclusion[†] into clustering analysis |
|---|---|---|---|---|---|---|---|
| **French Guiana** | Q1 | Shifted year | 7 | 11 | 7 | No | Exclude: no data for comparison matrix year |
| **Gabon** | Q1 | Shifted year | 0 | 2 | 0 | Not applicable: too few time windows for analysis | Exclude: too few time windows |
| **Georgia** | Q1 | Shifted year | 9 | 11 | 8 | Yes | Include |
| **Germany** | Q1 | Shifted year | 11 | 11 | 9 | Yes | Include |
| **Ghana** | Q4 | Shifted year | 11 | 11 | 9 | Yes | Include |
| **Greece** | Q1 | Shifted year | 10 | 11 | 9 | Yes | Include |
| **Guadeloupe** | Q1 | Shifted year | 2 | 11 | 2 | Not applicable: too few time windows for analysis | Exclude: too few time windows |
| **Guatemala** | Q1 | Shifted year | 11 | 11 | 9 | Yes | Include |
| **Guinea** | Q4 | Shifted year | 2 | 4 | 2 | Not applicable: too few time windows for analysis | Exclude: too few time windows |
| **Guyana** | Q3 | Calendar year | 1 | 1 | 1 | Not applicable: too few time windows for analysis | Exclude: too few time windows |
| **Haiti** | Q4 | Shifted year | 3 | 5 | 3 | Not applicable: too few time windows for analysis | Exclude: too few time windows |
| **Honduras** | Q2 | Calendar year | 8 | 10 | 8 | Yes | Include |
| **Hungary** | Q1 | Shifted year | 11 | 11 | 9 | Yes | Include |
| **Iceland** | Q1 | Shifted year | 11 | 11 | 9 | Yes | Include |

| Country | Calendar quarter (Q) in which most influenza virus detections occurred | Time window used for analysis | Number of time windows with ≥100 detections | Number of time windows with data for ≥40% of weeks | Number of complete time windows available for analysis | Data available during anchor year* | Inclusion† into clustering analysis |
|---|---|---|---|---|---|---|---|
| **India** | Q1 | Shifted year | 11 | 11 | 9 | Yes | Include |
| **Indonesia** | Q1 | Shifted year | 11 | 11 | 9 | Yes | Include |
| **Iran** | Q1 | Shifted year | 11 | 11 | 9 | Yes | Include |
| **Iraq** | Q1 | Shifted year | 6 | 11 | 4 | Not applicable: too few time windows for analysis | Exclude: too few time windows |
| **Ireland** | Q1 | Shifted year | 11 | 11 | 9 | Yes | Include |
| **Israel** | Q1 | Shifted year | 11 | 11 | 9 | Yes | Include |
| **Italy** | Q1 | Shifted year | 11 | 11 | 9 | Yes | Include |
| **Jamaica** | Q1 | Shifted year | 6 | 11 | 5 | Yes | Include |
| **Japan** | Q1 | Shifted year | 11 | 11 | 9 | Yes | Include |
| **Jordan** | Q4 | Shifted year | 9 | 11 | 8 | Yes | Include |
| **Kazakhstan** | Q1 | Shifted year | 11 | 11 | 9 | Yes | Include |
| **Kenya** | Q3 | Calendar year | 7 | 8 | 7 | Yes | Include |
| **Kosovo** | Q1 | Shifted year | 4 | 5 | 3 | Not applicable: too few time windows for analysis | Exclude: too few time windows |
| **Kuwait** | Q4 | Shifted year | 3 | 3 | 2 | Not applicable: too few time windows for analysis | Exclude: too few time windows |

| Country | Calendar quarter (Q) in which most influenza virus detections occurred | Time window used for analysis | Number of time windows with ≥100 detections | Number of time windows with data for ≥40% of weeks | Number of complete time windows available for analysis | Data available during anchor year* | Inclusion† into clustering analysis |
|---|---|---|---|---|---|---|---|
| **Kyrgyzstan** | Q1 | Shifted year | 5 | 11 | 5 | Yes | Include |
| **Lao PDR** | Q4 | Shifted year | 10 | 11 | 9 | Yes | Include |
| **Latvia** | Q1 | Shifted year | 10 | 11 | 9 | Yes | Include |
| **Lebanon** | Q1 | Shifted year | 6 | 7 | 6 | Yes | Include |
| **Liberia** | Q3 | Calendar year | 0 | 1 | 0 | Not applicable: too few time windows for analysis | Exclude: too few time windows |
| **Libya** | Q1 | Shifted year | 1 | 2 | 1 | Not applicable: too few time windows for analysis | Exclude: too few time windows |
| **Liechtenstein** | Q1 | Shifted year | 0 | 2 | 0 | Not applicable: too few time windows for analysis | Exclude: too few time windows |
| **Lithuania** | Q1 | Shifted year | 10 | 11 | 8 | Yes | Include |
| **Luxembourg** | Q1 | Shifted year | 11 | 11 | 9 | Yes | Include |
| **Madagascar** | Q2 | Calendar year | 10 | 10 | 10 | Yes | Include |
| **Malaysia** | Q1 | Shifted year | 6 | 11 | 5 | Yes | Include |
| **Maldives** | Q2 | Calendar year | 4 | 6 | 4 | Not applicable: too few time windows for analysis | Exclude: too few time windows |
| **Mali** | Q1 | Shifted year | 4 | 10 | 2 | Not applicable: too few time windows for analysis | Exclude: too few time windows |
| **Malta** | Q1 | Shifted year | 7 | 10 | 6 | Yes | Include |

| Country | Calendar quarter (Q) in which most influenza virus detections occurred | Time window used for analysis | Number of time windows with ≥100 detections | Number of time windows with data for ≥40% of weeks | Number of complete time windows available for analysis | Data available during anchor year* | Inclusion† into clustering analysis |
|---|---|---|---|---|---|---|---|
| **Martinique** | Q1 | Shifted year | 5 | 11 | 5 | Yes | Include |
| **Mauritania** | Q4 | Shifted year | 0 | 6 | 0 | Not applicable: too few time windows for analysis | Exclude: too few time windows |
| **Mauritius** | Q3 | Calendar year | 5 | 8 | 5 | Yes | Include |
| **Mexico** | Q1 | Shifted year | 11 | 11 | 9 | Yes | Include |
| **Mongolia** | Q1 | Shifted year | 11 | 11 | 9 | Yes | Include |
| **Montenegro** | Q1 | Shifted year | 5 | 5 | 4 | Not applicable: too few time windows for analysis | Exclude: too few time windows |
| **Morocco** | Q1 | Shifted year | 9 | 11 | 8 | Yes | Include |
| **Mozambique** | Q1 | Shifted year | 3 | 9 | 3 | Not applicable: too few time windows for analysis | Exclude: too few time windows |
| **Myanmar** | Q3 | Calendar year | 5 | 5 | 5 | Yes | Include |
| **Namibia** | Q2 | Calendar year | 0 | 1 | 0 | Not applicable: too few time windows for analysis | Exclude: too few time windows |
| **Nepal** | Q3 | Calendar year | 9 | 9 | 9 | Yes | Include |
| **Netherlands** | Q1 | Shifted year | 11 | 11 | 9 | Yes | Include |
| **New Caledonia** | Q3 | Calendar year | 8 | 10 | 8 | Yes | Include |
| **New Zealand** | Q3 | Calendar year | 10 | 10 | 10 | Yes | Include |

| Country | Calendar quarter (Q) in which most influenza virus detections occurred | Time window used for analysis | Number of time windows with ≥100 detections | Number of time windows with data for ≥40% of weeks | Number of complete time windows available for analysis | Data available during anchor year* | Inclusion† into clustering analysis |
|---|---|---|---|---|---|---|---|
| **Nicaragua** | Q4 | Shifted year | 9 | 11 | 9 | Yes | Include |
| **Niger** | Q1 | Shifted year | 4 | 11 | 3 | Not applicable: too few time windows for analysis | Exclude: too few time windows |
| **Nigeria** | Q2 | Calendar year | 4 | 10 | 4 | Not applicable: too few time windows for analysis | Exclude: too few time windows |
| **North Korea** | Q1 | Shifted year | 2 | 1 | 0 | Not applicable: too few time windows for analysis | Exclude: too few time windows |
| **North Macedonia** | Q1 | Shifted year | 3 | 6 | 3 | Not applicable: too few time windows for analysis | Exclude: too few time windows |
| **Norway** | Q1 | Shifted year | 11 | 11 | 9 | Yes | Include |
| **Oman** | Q4 | Shifted year | 11 | 11 | 9 | Yes | Include |
| **Pakistan** | Q1 | Shifted year | 9 | 11 | 7 | Yes | Include |
| **Palestine** | Q1 | Shifted year | 3 | 4 | 3 | Not applicable: too few time windows for analysis | Exclude: too few time windows |
| **Panama** | Q3 | Calendar year | 8 | 10 | 8 | Yes | Include |
| **Papua New Guinea** | Q1 | Shifted year | 1 | 7 | 1 | Not applicable: too few time windows for analysis | Exclude: too few time windows |
| **Paraguay** | Q3 | Calendar year | 10 | 10 | 10 | Yes | Include |
| **Peru** | Q3 | Calendar year | 10 | 10 | 10 | Yes | Include |
| **Philippines** | Q3 | Calendar year | 10 | 10 | 10 | Yes | Include |

| Country | Calendar quarter (Q) in which most influenza virus detections occurred | Time window used for analysis | Number of time windows with ≥100 detections | Number of time windows with data for ≥40% of weeks | Number of complete time windows available for analysis | Data available during anchor year* | Inclusion† into clustering analysis |
|---|---|---|---|---|---|---|---|
| **Poland** | Q1 | Shifted year | 10 | 11 | 8 | Yes | Include |
| **Portugal** | Q1 | Shifted year | 11 | 11 | 9 | Yes | Include |
| **Qatar** | Q4 | Shifted year | 11 | 11 | 9 | Yes | Include |
| **Republic of Korea** | Q1 | Shifted year | 11 | 11 | 9 | Yes | Include |
| **Republic of Moldova** | Q1 | Shifted year | 9 | 11 | 8 | Yes | Include |
| **Romania** | Q1 | Shifted year | 11 | 11 | 9 | Yes | Include |
| **Russian Federation** | Q1 | Shifted year | 11 | 11 | 9 | Yes | Include |
| **Rwanda** | Q2 | Calendar year | 2 | 10 | 2 | Not applicable: too few time windows for analysis | Exclude: too few time windows |
| **Saint Kitts and Nevis** | Q1 | Shifted year | 0 | 0 | 0 | Not applicable: too few time windows for analysis | Exclude: too few time windows |
| **Saint Lucia** | Q1 | Shifted year | 0 | 4 | 0 | Not applicable: too few time windows for analysis | Exclude: too few time windows |
| **Saint Vincent and the Grenadines** | Q1 | Shifted year | 0 | 3 | 0 | Not applicable: too few time windows for analysis | Exclude: too few time windows |
| **Saudi Arabia** | Q4 | Shifted year | 5 | 5 | 4 | Not applicable: too few time windows for analysis | Exclude: too few time windows |
| **Senegal** | Q4 | Shifted year | 11 | 11 | 9 | Yes | Include |
| **Serbia** | Q1 | Shifted year | 10 | 11 | 9 | Yes | Include |

| Country | Calendar quarter (Q) in which most influenza virus detections occurred | Time window used for analysis | Number of time windows with ≥100 detections | Number of time windows with data for ≥40% of weeks | Number of complete time windows available for analysis | Data available during anchor year* | Inclusion† into clustering analysis |
|---|---|---|---|---|---|---|---|
| **Seychelles** | Q2 | Calendar year | 0 | 2 | 0 | Not applicable: too few time windows for analysis | Exclude: too few time windows |
| **Sierra Leone** | Q3 | Calendar year | 0 | 7 | 0 | Not applicable: too few time windows for analysis | Exclude: too few time windows |
| **Singapore** | Q1 | Shifted year | 11 | 11 | 9 | Yes | Include |
| **Slovakia** | Q1 | Shifted year | 10 | 11 | 8 | Yes | Include |
| **Slovenia** | Q1 | Shifted year | 11 | 11 | 9 | Yes | Include |
| **Somalia** | Q3 | Calendar year | 0 | 1 | 0 | Not applicable: too few time windows for analysis | Exclude: too few time windows |
| **South Africa** | Q3 | Calendar year | 10 | 10 | 10 | Yes | Include |
| **South Sudan** | Q3 | Calendar year | 1 | 2 | 1 | Not applicable: too few time windows for analysis | Exclude: too few time windows |
| **Spain** | Q1 | Shifted year | 11 | 11 | 9 | Yes | Include |
| **Sri Lanka** | Q2 | Calendar year | 10 | 10 | 10 | Yes | Include |
| **Suriname** | Q1 | Shifted year | 1 | 7 | 1 | Not applicable: too few time windows for analysis | Exclude: too few time windows |
| **Sweden** | Q1 | Shifted year | 11 | 11 | 9 | Yes | Include |
| **Switzerland** | Q1 | Shifted year | 11 | 11 | 9 | Yes | Include |
| **Syrian Arab Republic** | Q1 | Shifted year | 1 | 2 | 1 | Not applicable: too few time windows for analysis | Exclude: too few time windows |

| Country | Calendar quarter (Q) in which most influenza virus detections occurred | Time window used for analysis | Number of time windows with ≥100 detections | Number of time windows with data for ≥40% of weeks | Number of complete time windows available for analysis | Data available during anchor year* | Inclusion[†] into clustering analysis |
|---|---|---|---|---|---|---|---|
| **Tajikistan** | Q1 | Shifted year | 1 | 4 | 1 | Not applicable: too few time windows for analysis | Exclude: too few time windows |
| **Tanzania** | Q1 | Shifted year | 11 | 11 | 9 | Yes | Include |
| **Thailand** | Q3 | Calendar year | 10 | 10 | 10 | Yes | Include |
| **Timor-Leste** | Q1 | Shifted year | 3 | 4 | 3 | Not applicable: too few time windows for analysis | Exclude: too few time windows |
| **Togo** | Q4 | Shifted year | 7 | 11 | 6 | Yes | Include |
| **Trinidad and Tobago** | Q4 | Shifted year | 0 | 5 | 0 | Not applicable: too few time windows for analysis | Exclude: too few time windows |
| **Tunisia** | Q1 | Shifted year | 8 | 11 | 7 | Yes | Include |
| **Turkey** | Q1 | Shifted year | 10 | 11 | 9 | Yes | Include |
| **Turkmenistan** | Q1 | Shifted year | 0 | 2 | 0 | Not applicable: too few time windows for analysis | Exclude: too few time windows |
| **Turks and Caicos Islands** | Q1 | Shifted year | 0 | 0 | 0 | Not applicable: too few time windows for analysis | Exclude: too few time windows |
| **Uganda** | Q3 | Calendar year | 9 | 10 | 9 | Yes | Include |
| **Ukraine** | Q1 | Shifted year | 10 | 11 | 9 | Yes | Include |
| **United Arab Emirates** | Q4 | Shifted year | 2 | 2 | 1 | Not applicable: too few time windows for analysis | Exclude: too few time windows |
| **United Kingdom** | Q1 | Shifted year | 11 | 11 | 9 | Yes | Include |

| Country | Calendar quarter (Q) in which most influenza virus detections occurred | Time window used for analysis | Number of time windows with ≥100 detections | Number of time windows with data for ≥40% of weeks | Number of complete time windows available for analysis | Data available during anchor year* | Inclusion† into clustering analysis |
|---|---|---|---|---|---|---|---|
| **United States of America** | Q1 | Shifted year | 11 | 11 | 9 | Yes | Include |
| **Uruguay** | Q3 | Calendar year | 6 | 10 | 6 | Yes | Include |
| **Uzbekistan** | Q1 | Shifted year | 2 | 11 | 2 | Not applicable: too few time windows for analysis | Exclude: too few time windows |
| **Venezuela** | Q3 | Calendar year | 4 | 8 | 4 | Not applicable: too few time windows for analysis | Exclude: too few time windows |
| **Viet Nam** | Q3 | Calendar year | 10 | 10 | 10 | Yes | Include |
| **Yemen** | Q1 | Shifted year | 1 | 3 | 1 | Not applicable: too few time windows for analysis | Exclude: too few time windows |
| **Zambia** | Q3 | Calendar year | 8 | 10 | 8 | Yes | Include |
| **Zimbabwe** | Q2 | Calendar year | 0 | 1 | 0 | Not applicable: too few time windows for analysis | Exclude: too few time windows |

* Countries had to have ≥1 complete country-to-country time-window year available for comparison and for later determination of ideal cluster size, so only countries that had data reported during the time-window year with maximum country representation (anchor year: 2019 for calendar-year and 2015 for shifted-year groupings) were included.

† A country was included in the clustering analysis if there were ≥5 complete time windows of surveillance data where each time window included ≥100 influenza detections and ≥40% of the weeks had data reported, and if they had data available during the anchor year.

**Supplementary Table 2:** Cluster consensus scores assessing the stability of country assignments to clusters. The cluster consensus score is calculated from taking the mean of constituent country consensus scores. Country consensus scores averaged the pairwise consensus value between each country and its co-clustered countries to determine how consistently a given country was grouped with co-clustered countries across resamples. Higher scores demonstrated greater stability.

| Epidemic zone | Mean consensus score (%) |
|---|---|
| **A** | 61.8 |
| **B** | 46.9 |
| **C** | 83.3 |
| **D** | 78.5 |
| **E** | 60.4 |

**Supplementary Table 3:** Country consensus scores assessing the stability of country assignments to clusters. Scores averaged the pairwise consensus value between each country and its co-clustered countries to determine how consistently a given country was grouped with co-clustered countries across resamples. Higher scores demonstrated greater stability.

| Country | Epidemic zone | Number of clustering opportunities | Consensus score (%) |
|---|---|---|---|
| **Belarus** | A | 8118 | 71.2 |
| **Austria** | A | 9877 | 70.6 |
| **Germany** | A | 9929 | 69.2 |
| **Hungary** | A | 9973 | 69.1 |
| **Netherlands** | A | 9901 | 68.1 |
| **Romania** | A | 9917 | 67.9 |
| **Latvia** | A | 9042 | 67.5 |
| **Slovakia** | A | 9020 | 67.4 |
| **Poland** | A | 8966 | 67.2 |
| **Luxembourg** | A | 9957 | 67.1 |
| **Slovenia** | A | 9931 | 66.2 |
| **Czechia** | A | 9034 | 65.8 |
| **Lithuania** | A | 9015 | 65.3 |
| **Serbia** | A | 8986 | 65.2 |
| **Croatia** | A | 9924 | 64.6 |
| **Estonia** | A | 9000 | 64.2 |
| **Russian Federation** | A | 9965 | 64.0 |
| **Belgium** | A | 9895 | 63.3 |
| **Sweden** | A | 9915 | 63.0 |
| **France** | A | 9932 | 62.7 |
| **Republic of Moldova** | A | 8100 | 62.4 |
| **Lebanon** | A | 5403 | 62.4 |
| **Switzerland** | A | 9995 | 60.6 |
| **Albania** | A | 7232 | 60.5 |
| **Martinique** | A | 4537 | 59.9 |
| **Denmark** | A | 9010 | 59.9 |
| **Tunisia** | A | 7228 | 56.9 |
| **Norway** | A | 9846 | 55.5 |
| **Greece** | A | 8965 | 54.1 |
| **Iceland** | A | 9893 | 52.7 |
| **Ukraine** | A | 9011 | 52.2 |
| **Jamaica** | A | 5360 | 47.8 |
| **Finland** | A | 8976 | 45.6 |
| **Armenia** | A | 5395 | 42.6 |
| **Algeria** | B | 5429 | 59.8 |

| Country | Epidemic zone | Number of clustering opportunities | Consensus score (%) |
|---|---|---|---|
| **Malta** | B | 6293 | 58.5 |
| **Spain** | B | 9905 | 54.4 |
| **Ireland** | B | 9855 | 53.3 |
| **Italy** | B | 9894 | 53.3 |
| **United Kingdom** | B | 9842 | 52.3 |
| **Portugal** | B | 9944 | 51.2 |
| **Mongolia** | B | 9927 | 49.8 |
| **Israel** | B | 9884 | 49.0 |
| **Pakistan** | B | 8062 | 48.6 |
| **Canada** | B | 9925 | 48.6 |
| **Bulgaria** | B | 9024 | 48.5 |
| **Morocco** | B | 8105 | 47.4 |
| **Japan** | B | 9864 | 47.4 |
| **Turkey** | B | 8959 | 44.8 |
| **United States of America** | B | 9924 | 43.1 |
| **Republic of Korea** | B | 9874 | 43.1 |
| **Jordan** | B | 8083 | 43.0 |
| **Kazakhstan** | B | 9891 | 41.2 |
| **China** | B | 9901 | 40.9 |
| **Iran** | B | 9908 | 40.4 |
| **Georgia** | B | 8132 | 39.2 |
| **Mexico** | B | 9928 | 39.0 |
| **Kyrgyzstan** | B | 4468 | 29.9 |
| **Lao PDR** | C | 8948 | 90.8 |
| **Cameroon** | C | 9011 | 90.7 |
| **Côte d'Ivoire** | C | 9034 | 90.7 |
| **Nicaragua** | C | 8048 | 90.7 |
| **Ghana** | C | 9859 | 90.1 |
| **Singapore** | C | 9883 | 90.1 |
| **Senegal** | C | 9942 | 90.0 |
| **Qatar** | C | 9875 | 89.9 |
| **Togo** | C | 6335 | 89.5 |
| **Malaysia** | C | 5387 | 89.2 |
| **Oman** | C | 9914 | 88.4 |
| **Indonesia** | C | 9984 | 86.4 |
| **Congo (DRC)** | C | 5411 | 83.5 |
| **Costa Rica** | C | 9012 | 80.8 |
| **Tanzania** | C | 9900 | 80.2 |
| **Ethiopia** | C | 6298 | 77.0 |

| Country | Epidemic zone | Number of clustering opportunities | Consensus score (%) |
|---|---|---|---|
| **Ecuador** | C | 9904 | 74.9 |
| **Egypt** | C | 9907 | 74.5 |
| **Guatemala** | C | 9855 | 73.4 |
| **Bahrain** | C | 6364 | 73.2 |
| **India** | C | 9925 | 71.2 |
| **China, Hong Kong SAR** | C | 9935 | 67.1 |
| **Bhutan** | D | 7146 | 87.5 |
| **Zambia** | D | 7210 | 86.0 |
| **Uganda** | D | 8038 | 85.1 |
| **Madagascar** | D | 8984 | 84.5 |
| **Viet Nam** | D | 9017 | 83.3 |
| **Kenya** | D | 6299 | 82.6 |
| **Colombia** | D | 8996 | 81.4 |
| **Honduras** | D | 7131 | 79.4 |
| **Sri Lanka** | D | 9009 | 79.3 |
| **Mauritius** | D | 4479 | 79.1 |
| **Philippines** | D | 9042 | 77.7 |
| **Thailand** | D | 8944 | 77.3 |
| **Cambodia** | D | 8963 | 77.1 |
| **New Caledonia** | D | 7111 | 76.3 |
| **Myanmar** | D | 4482 | 75.7 |
| **Nepal** | D | 8020 | 72.0 |
| **Peru** | D | 9004 | 71.2 |
| **Cuba** | D | 9020 | 70.3 |
| **Brazil** | D | 8930 | 65.4 |
| **Uruguay** | E | 5363 | 70.3 |
| **Chile** | E | 9011 | 67.8 |
| **Argentina** | E | 9030 | 66.2 |
| **Paraguay** | E | 8931 | 65.0 |
| **Fiji** | E | 4532 | 64.3 |
| **Panama** | E | 7126 | 64.1 |
| **Bangladesh** | E | 8975 | 63.0 |
| **New Zealand** | E | 8984 | 55.0 |
| **Bolivia** | E | 8993 | 52.8 |
| **South Africa** | E | 8979 | 52.5 |
| **El Salvador** | E | 7238 | 52.2 |
| **Australia** | E | 9013 | 51.9 |

**Supplementary Table 4:** Robustness of leave-one-out approach where countries included in the original clustering analysis were synthetically left out of the clustering analysis and assigned to an epidemic zone based on their population-weighted mean latitude. The percent match was quantified per cluster as the number of countries that were assigned to the same cluster via the original clustering and proxying approaches over the total number of countries in the cluster.

| Epidemic zone | Number of countries assigned via ensemble consensus clustering | Number of countries assigned to original cluster via proxying by latitude | Percent match (%) |
|---|---|---|---|
| **A** | 34 | 27 | 79.4 |
| **B** | 24 | 17 | 70.8 |
| **C** | 22 | 19 | 86.4 |
| **D** | 19 | 13 | 68.4 |
| **E** | 12 | 9 | 75.0 |

**Supplementary Table 5**: Country, population-weighted mean latitude, whether that assignment is from the primary clustering analysis (original) or from population-weighted mean latitude (proxy), influenza epidemic zone assignment, and weeks of main influenza activity.

| Country | Population-weighted mean latitude (°) | Epidemic zone assignment based on main analysis or proxying by latitude | Epidemic zone | Calendar months of typical influenza epidemics* |
|---|---|---|---|---|
| **Greenland** | 65.516 | proxy | A | [November–May] |
| **Iceland** | 64.276 | original | A | November–May |
| **Faroe Islands** | 62.026 | proxy | A | [November–May] |
| **Finland** | 61.472 | original | A | November–May |
| **Norway** | 60.802 | original | A | November–May |
| **Estonia** | 59.106 | original | A | December–June |
| **Sweden** | 58.836 | original | A | November–May |
| **Latvia** | 56.852 | original | A | December–June |
| **Denmark** | 55.823 | original | A | November–May |
| **Lithuania** | 55.070 | original | A | November–May |
| **Russian Federation** | 54.271 | original | A | November–May |
| **Isle of Man** | 54.180 | proxy | A | [November–May] |
| **Belarus** | 53.564 | original | A | December–June |
| **Netherlands** | 52.076 | original | A | November–May |
| **Poland** | 51.753 | original | A | November–May |
| **Germany** | 51.028 | original | A | November–May |
| **Belgium** | 50.836 | original | A | November–May |
| **Czechia** | 49.851 | original | A | November–May |
| **Luxembourg** | 49.618 | original | A | November–May |
| **Guernsey** | 49.468 | proxy | A | [November–May] |
| **Jersey** | 49.194 | proxy | A | [November–May] |
| **Ukraine** | 48.646 | original | A | November–May |
| **Slovakia** | 48.580 | original | A | November–May |
| **Austria** | 47.820 | original | A | November–May |
| **Hungary** | 47.346 | original | A | November–May |
| **Liechtenstein** | 47.161 | proxy | A | [November–May] |
| **Republic of Moldova** | 47.104 | original | A | November–May |
| **Switzerland** | 47.078 | original | A | November–May |
| **France** | 47.072 | original | A | November–May |
| **Saint Pierre and Miquelon** | 46.880 | proxy | A | [November–May] |
| **Slovenia** | 46.162 | original | A | November–May |
| **Romania** | 45.605 | original | A | November–May |
| **Croatia** | 45.189 | original | A | November–May |
| **Serbia** | 44.611 | original | A | December–May |
| **Bosnia and Herzegovina** | 44.240 | proxy | A | [November–May] |
| **San Marino** | 43.945 | proxy | A | [November–May] |

| Country | Population-weighted mean latitude (°) | Epidemic zone assignment based on main analysis or proxying by latitude | Epidemic zone | Calendar months of typical influenza epidemics* |
|---|---|---|---|---|
| **Monaco** | 43.737 | proxy | A | [November–May] |
| **Albania** | 41.099 | original | A | November–May |
| **Armenia** | 40.248 | original | A | November–May |
| **Greece** | 38.685 | original | A | December–June |
| **Tunisia** | 35.847 | original | A | November–May |
| **Lebanon** | 33.858 | original | A | November–May |
| **Jamaica** | 18.041 | original | A | November–May |
| **Martinique** | 14.632 | original | A | November–May |
| **Åland Islands** | NA | proxy | A | [November–May] |
| **Svalbard and Jan Mayen Islands** | NA | proxy | A | [November–May] |
| **Gibraltar** | NA | proxy | A | [November–May] |
| **Ireland** | 53.148 | original | B | November–May |
| **United Kingdom** | 52.565 | original | B | November–May |
| **Mongolia** | 47.935 | original | B | November–May |
| **Kazakhstan** | 46.965 | original | B | November–May |
| **Canada** | 46.432 | original | B | November–May |
| **Italy** | 42.896 | original | B | November–May |
| **Bulgaria** | 42.742 | original | B | November–May |
| **Kosovo** | 42.561 | proxy | B | [November–May] |
| **Montenegro** | 42.529 | proxy | B | [November–May] |
| **Andorra** | 42.518 | proxy | B | [November–May] |
| **Georgia** | 41.912 | original | B | November–May |
| **Vatican** | 41.903 | proxy | B | [November–May] |
| **Kyrgyzstan** | 41.889 | original | B | October–April |
| **North Macedonia** | 41.773 | proxy | B | [November–May] |
| **Uzbekistan** | 40.661 | proxy | B | [November–May] |
| **Azerbaijan** | 40.304 | proxy | B | [November–May] |
| **Spain** | 39.737 | original | B | November–May |
| **North Korea** | 39.501 | proxy | B | [November–May] |
| **Portugal** | 39.491 | original | B | November–May |
| **Turkey** | 39.450 | original | B | November–May |
| **Tajikistan** | 39.061 | proxy | B | [November–May] |
| **Turkmenistan** | 38.774 | proxy | B | [November–May] |
| **Azores** | 38.323 | proxy | B | [November–May] |
| **United States of America** | 37.607 | original | B | November–May |
| **Republic of Korea** | 36.594 | original | B | November–May |
| **Malta** | 35.902 | original | B | November–April |
| **Japan** | 35.573 | original | B | November–May |
| **Algeria** | 35.570 | original | B | October–April |

| Country | Population-weighted mean latitude (°) | Epidemic zone assignment based on main analysis or proxying by latitude | Epidemic zone | Calendar months of typical influenza epidemics* |
|---|---|---|---|---|
| **Syria** | 35.140 | proxy | B | [November–May] |
| **Cyprus** | 34.999 | proxy | B | [November–May] |
| **Afghanistan** | 34.597 | proxy | B | [November–May] |
| **Iran** | 34.413 | original | B | October–April |
| **Iraq** | 33.599 | proxy | B | [November–May] |
| **Morocco** | 33.392 | original | B | October–April |
| **Madeira Islands** | 32.770 | proxy | B | [November–May] |
| **Bermuda** | 32.354 | proxy | B | [November–May] |
| **Israel** | 32.109 | original | B | November–May |
| **Jordan** | 32.030 | original | B | November–May |
| **China** | 31.892 | original | B | November–May |
| **Libya** | 31.889 | proxy | B | [November–May] |
| **Palestine** | 31.764 | proxy | B | [November–May] |
| **Pakistan** | 30.296 | original | B | October–April |
| **Kuwait** | 29.200 | proxy | B | [November–May] |
| **Canary Islands** | 28.356 | proxy | B | [November–May] |
| **Western Sahara** | 26.066 | proxy | B | [November–May] |
| **Bahamas** | 25.430 | proxy | B | [November–May] |
| **United Arab Emirates** | 24.993 | proxy | B | [November–May] |
| **Mexico** | 21.302 | original | B | October–April |
| **China, Macao SAR** | NA | proxy | B | [November–May] |
| **Egypt** | 29.890 | original | C | October–April |
| **Bahrain** | 26.173 | original | C | September–March |
| **Qatar** | 25.304 | original | C | October–April |
| **Taiwan** | 24.579 | proxy | C | [September–March] |
| **Saudi Arabia** | 23.761 | proxy | C | [September–March] |
| **Oman** | 23.274 | original | C | October–February |
| **China, Hong Kong SAR** | 22.334 | original | C | January–May |
| **Turks and Caicos Islands** | 21.747 | proxy | C | [September–March] |
| **India** | 21.157 | original | C | July–October |
| **Cayman Islands** | 19.312 | proxy | C | [September–March] |
| **Haiti** | 18.763 | proxy | C | [September–March] |
| **British Virgin Islands** | 18.427 | proxy | C | [September–March] |
| **Puerto Rico** | 18.314 | proxy | C | [September–March] |
| **Anguilla** | 18.209 | proxy | C | [September–March] |
| **Saint Martin** | 18.068 | proxy | C | [September–March] |
| **Sint Maarten** | 18.046 | proxy | C | [September–March] |
| **US Virgin Islands** | 17.913 | proxy | C | [September–March] |
| **Mauritania** | 17.909 | proxy | C | [September–March] |
| **Saint Barthélemy** | 17.896 | proxy | C | [September–March] |

| Country | Population-weighted mean latitude (°) | Epidemic zone assignment based on main analysis or proxying by latitude | Epidemic zone | Calendar months of typical influenza epidemics* |
|---|---|---|---|---|
| **Lao PDR** | 17.818 | original | C | August–February |
| **Bonaire, Sint Eustatius and Saba** | 17.502 | proxy | C | [September–March] |
| **Belize** | 17.471 | proxy | C | [September–March] |
| **Saint Kitts and Nevis** | 17.289 | proxy | C | [September–March] |
| **Nevis** | 17.149 | proxy | C | [September–March] |
| **Montserrat** | 16.770 | proxy | C | [September–March] |
| **Guadeloupe** | 16.209 | proxy | C | [September–March] |
| **Cape Verde** | 15.680 | proxy | C | [September–March] |
| **Dominica** | 15.383 | proxy | C | [September–March] |
| **Eritrea** | 15.221 | proxy | C | [September–March] |
| **Northern Mariana Islands** | 15.154 | proxy | C | [September–March] |
| **Guatemala** | 14.765 | original | C | January–July |
| **Sudan** | 14.671 | proxy | C | [September–March] |
| **Senegal** | 14.651 | original | C | July–December |
| **Yemen** | 14.497 | proxy | C | [September–March] |
| **Niger** | 14.081 | proxy | C | [September–March] |
| **Saint Lucia** | 13.945 | proxy | C | [September–March] |
| **Guam** | 13.476 | proxy | C | [September–March] |
| **Gambia** | 13.408 | proxy | C | [September–March] |
| **Saint Vincent and the Grenadines** | 13.152 | proxy | C | [September–March] |
| **Barbados** | 13.114 | proxy | C | [September–March] |
| **Mali** | 12.965 | proxy | C | [September–March] |
| **Aruba** | 12.521 | proxy | C | [September–March] |
| **Nicaragua** | 12.357 | original | C | July–December |
| **Burkina Faso** | 12.152 | proxy | C | [September–March] |
| **Curacao** | 12.135 | proxy | C | [September–March] |
| **Grenada** | 12.135 | proxy | C | [September–March] |
| **Guinea-Bissau** | 11.949 | proxy | C | [September–March] |
| **Chad** | 11.568 | proxy | C | [September–March] |
| **Djibouti** | 11.553 | proxy | C | [September–March] |
| **Trinidad and Tobago** | 10.478 | proxy | C | [September–March] |
| **Costa Rica** | 9.971 | original | C | October–January |
| **Guinea** | 9.718 | proxy | C | [September–March] |
| **Ethiopia** | 9.300 | original | C | October–April |
| **Marshall Islands** | 7.665 | proxy | C | [September–March] |
| **Benin** | 7.651 | proxy | C | [September–March] |
| **Togo** | 6.848 | original | C | September–January |
| **Côte d'Ivoire** | 6.511 | original | C | September–December |
| **Ghana** | 6.367 | original | C | July–November |

| Country | Population-weighted mean latitude (°) | Epidemic zone assignment based on main analysis or proxying by latitude | Epidemic zone | Calendar months of typical influenza epidemics* |
|---|---|---|---|---|
| **Suriname** | 5.798 | proxy | C | [September–March] |
| **Cameroon** | 5.424 | original | C | August–January |
| **French Guiana** | 5.019 | proxy | C | [September–March] |
| **Malaysia** | 3.483 | original | C | January–July |
| **Singapore** | 1.326 | original | C | December–March |
| **Gabon** | -0.151 | proxy | C | [September–March] |
| **Ecuador** | -1.404 | original | C | December–April |
| **Congo (DRC)** | -4.725 | original | C | November–April |
| **Indonesia** | -4.806 | original | C | November–May |
| **Tanzania** | -5.893 | original | C | December–June |
| **Papua New Guinea** | -7.347 | proxy | C | [September–March] |
| **Timor-Leste** | -8.688 | proxy | C | [September–March] |
| **Mozambique** | -19.942 | proxy | C | [September–March] |
| **Nepal** | 27.587 | original | D | January–April |
| **Bhutan** | 27.268 | original | D | February–May |
| **Cuba** | 22.222 | original | D | June–December |
| **Dominican Republic** | 18.769 | proxy | D | [June–November] |
| **Myanmar** | 18.541 | original | D | June–December |
| **Antigua and Barbuda** | 17.127 | proxy | D | [June–November] |
| **Honduras** | 14.890 | original | D | May–November |
| **Viet Nam** | 14.675 | original | D | May–September |
| **Thailand** | 13.771 | original | D | June–November |
| **Philippines** | 12.220 | original | D | June–November |
| **Cambodia** | 11.698 | original | D | June–December |
| **Venezuela** | 9.861 | proxy | D | [June–November] |
| **Sierra Leone** | 8.382 | proxy | D | [June–November] |
| **Nigeria** | 7.935 | proxy | D | [June–November] |
| **Palau** | 7.346 | proxy | D | [June–November] |
| **Sri Lanka** | 7.272 | original | D | April–July |
| **Micronesia** | 7.161 | proxy | D | [June–November] |
| **South Sudan** | 6.614 | proxy | D | [June–November] |
| **Guyana** | 6.612 | proxy | D | [June–November] |
| **Liberia** | 6.364 | proxy | D | [June–November] |
| **Colombia** | 5.946 | original | D | April–August |
| **Central African Republic** | 4.964 | proxy | D | [June–November] |
| **Somalia** | 4.808 | proxy | D | [June–November] |
| **Brunei** | 4.548 | proxy | D | [June–November] |
| **Maldives** | 3.572 | proxy | D | [June–November] |
| **Equatorial Guinea** | 2.471 | proxy | D | [June–November] |
| **Kiribati** | 1.444 | proxy | D | [June–November] |

| Country | Population-weighted mean latitude (°) | Epidemic zone assignment based on main analysis or proxying by latitude | Epidemic zone | Calendar months of typical influenza epidemics* |
|---|---|---|---|---|
| **Sao Tome and Principe** | 0.859 | proxy | D | [June–November] |
| **Uganda** | 0.590 | original | D | June–December |
| **Nauru** | -0.529 | proxy | D | [June–November] |
| **Kenya** | -0.877 | original | D | February–April |
| **Rwanda** | -2.034 | proxy | D | [June–November] |
| **Burundi** | -3.349 | proxy | D | [June–November] |
| **Republic of Congo** | -3.906 | proxy | D | [June–November] |
| **Seychelles** | -4.646 | proxy | D | [June–November] |
| **Peru** | -11.139 | original | D | April–October |
| **Zambia** | -13.858 | original | D | June–December |
| **Brazil** | -17.685 | original | D | March–September |
| **Madagascar** | -18.910 | original | D | January–July |
| **Mauritius** | -20.224 | original | D | February–September |
| **New Caledonia** | -21.853 | original | D | June–October |
| **Bangladesh** | 23.653 | original | E | April–October |
| **El Salvador** | 13.698 | original | E | May–August |
| **Panama** | 8.859 | original | E | May–September |
| **Chagos Archipelago** | -7.321 | proxy | E | [May–November] |
| **Tuvalu** | -8.289 | proxy | E | [May–November] |
| **Tokelau** | -8.899 | proxy | E | [May–November] |
| **Solomon Islands** | -9.263 | proxy | E | [May–November] |
| **Christmas Island** | -10.422 | proxy | E | [May–November] |
| **Angola** | -10.445 | proxy | E | [May–November] |
| **Comoros** | -11.980 | proxy | E | [May–November] |
| **Cocos Islands** | -12.157 | proxy | E | [May–November] |
| **Mayotte** | -12.794 | proxy | E | [May–November] |
| **Wallis and Futuna** | -13.663 | proxy | E | [May–November] |
| **Samoa** | -13.804 | proxy | E | [May–November] |
| **Malawi** | -14.258 | proxy | E | [May–November] |
| **American Samoa** | -14.309 | proxy | E | [May–November] |
| **Saint Helena** | -15.964 | proxy | E | [May–November] |
| **Vanuatu** | -16.989 | proxy | E | [May–November] |
| **Bolivia** | -17.385 | original | E | March–August |
| **French Polynesia** | -17.462 | proxy | E | [May–November] |
| **Fiji** | -17.823 | original | E | February–June |
| **Zimbabwe** | -18.716 | proxy | E | [May–November] |
| **Niue** | -19.053 | proxy | E | [May–November] |
| **Tonga** | -20.778 | proxy | E | [May–November] |
| **Reunion** | -21.092 | proxy | E | [May–November] |
| **Cook Islands** | -21.208 | proxy | E | [May–November] |

| Country | Population-weighted mean latitude (°) | Epidemic zone assignment based on main analysis or proxying by latitude | Epidemic zone | Calendar months of typical influenza epidemics* |
|---|---|---|---|---|
| **Namibia** | -21.820 | proxy | E | [May–November] |
| **Botswana** | -23.265 | proxy | E | [May–November] |
| **Pitcairn Islands** | -25.066 | proxy | E | [May–November] |
| **Paraguay** | -25.291 | original | E | May–September |
| **Swaziland** | -26.460 | proxy | E | [May–November] |
| **South Africa** | -28.396 | original | E | April–October |
| **Norfolk Island** | -29.055 | proxy | E | [May–November] |
| **Lesotho** | -29.418 | proxy | E | [May–November] |
| **Argentina** | -32.600 | original | E | May–November |
| **Australia** | -33.130 | original | E | May–November |
| **Chile** | -33.478 | original | E | May–November |
| **Uruguay** | -34.085 | original | E | May–November |
| **New Zealand** | -39.368 | original | E | May–November |
| **French Southern and Antarctic Lands** | -49.059 | proxy | E | [May–November] |
| **Falkland Islands** | -51.694 | proxy | E | [May–November] |
| **Heard Island and McDonald Islands** | -53.076 | proxy | E | [May–November] |
| **South Georgia** | -54.336 | proxy | E | [May–November] |
| **South Sandwich Islands** | -58.423 | proxy | E | [May–November] |
| **Antarctica** | -72.512 | proxy | E | [May–November] |

* The generalized additive model (GAM) fitted values for each country's relative influenza burden were used to determine the calendar months when influenza epidemics would most likely occur. Here, we identified the main influenza epidemic for each country, which we defined as the influenza epidemic with the longest duration. We defined the start of influenza epidemics as the first week in which the weekly relative influenza burden exceeded the median of the weekly relative influenza burden values for ≥3 weeks and the end of influenza epidemics as the first week in which the burden remained below the median for ≥3 weeks. Calendar weeks were then converted to months using the average number of weeks per month (4.345). If the epidemic zone was determined for a country by their population-weighted mean latitude, then the cluster-specific GAM fitted values were used to approximate the epidemic period (in brackets).

**Supplementary Table 6:** Summary seasonal start and end of each epidemic zone's main influenza epidemics. Summary seasonal curves were calculated for each zone using hierarchical GAMs for on country-year influenza burden. The main influenza epidemic is defined as the influenza epidemic with the longest duration. The start of influenza epidemics is the first week in which the weekly relative influenza burden exceeded the median of the weekly relative influenza burden values for ≥3 weeks and the end of influenza epidemics is the first week in which the burden remained below the median for ≥3 weeks. Calendar weeks were converted to months using the average number of weeks per month (4.345). Area under the curve calculations were used to estimate what proportion of influenza activity occurred during the main epidemic.

| | **Epidemic start** | | **Epidemic end** | | |
|---|---|---|---|---|---|
| **Epidemic zone** | **Week** | **Month** | **Week** | **Month** | **% of annual influenza burden** |
| **A** | 47 | November | 21 | May | 98.4 |
| **B** | 45 | November | 19 | May | 96.5 |
| **C** | 39 | September | 13 | March | 56.5 |
| **D** | 22 | June | 45 | November | 55.3 |
| **E** | 18 | May | 44 | November | 82.4 |
